# Whole genome sequencing identifies common and rare structural variants contributing to hematologic traits in the NHLBI TOPMed program

**DOI:** 10.1101/2021.12.16.21267871

**Authors:** Marsha M. Wheeler, Adrienne M. Stilp, Shuquan Rao, Bjarni V. Halldórsson, Doruk Beyter, Jia Wen, Anna V. Mikhaylova, Caitlin P. McHugh, John Lane, Min-Zhi Jiang, Laura M. Raffield, Goo Jun, Fritz J. Sedlazeck, Ginger Metcalf, Yao Yao, Joshua B. Bis, Nathalie Chami, Paul S. de Vries, Pinkal Desai, James S. Floyd, Yan Gao, Kai Kammers, Wonji Kim, Jee-Young Moon, Aakrosh Ratan, Lisa R. Yanek, Laura Almasy, Lewis C. Becker, John Blangero, Michael H. Cho, Joanne E. Curran, Myriam Fornage, Robert C. Kaplan, Joshua P. Lewis, Ruth J.F. Loos, Braxton D. Mitchell, Alanna C. Morrison, Michael Preuss, Bruce M. Psaty, Stephen S. Rich, Jerome I. Rotter, Hua Tang, Russell P. Tracy, Eric Boerwinkle, Goncalo Abecasis, Thomas W. Blackwell, Albert V. Smith, Andrew D. Johnson, Rasika A. Mathias, Deborah A. Nickerson, Matthew P. Conomos, Yun Li, NHLBI Trans-Omics for Precision Medicine (TOPMed) Consortium, Unnur Þorsteinsdóttir, Magnús K. Magnússon, Kari Stefansson, Nathan D. Pankratz, Daniel E. Bauer, Paul L. Auer, Alex P. Reiner

## Abstract

Genome-wide association studies (GWAS) have identified thousands of single nucleotide variants and small indels that contribute to the genetic architecture of hematologic traits. While structural variants (SVs) are known to cause rare blood or hematopoietic disorders, the genome-wide contribution of SVs to quantitative blood cell trait variation is unknown. Here we utilized SVs detected from whole genome sequencing (WGS) in ancestrally diverse participants of the NHLBI TOPMed program (N=50,675). Using single variant tests, we assessed the association of common and rare SVs with red cell-, white cell-, and platelet-related quantitative traits. The results show 33 independent SVs (23 common and 10 rare) reaching genome-wide significance. The majority of significant association signals (N=27) replicated in independent datasets from deCODE genetics and the UK BioBank. Moreover, most trait-associated SVs (N=24) are within 1Mb of previously-reported GWAS loci. SV analyses additionally discovered an association between a complex structural variant on 17p11.2 and white blood cell-related phenotypes. Based on functional annotation, the majority of significant SVs are located in non-coding regions (N=26) and predicted to impact regulatory elements and/or local chromatin domain boundaries in blood cells. We predict that several trait-associated SVs represent the causal variant. This is supported by genome-editing experiments which provide evidence that a deletion associated with lower monocyte counts leads to disruption of an *S1PR3* monocyte enhancer and decreased *S1PR3* expression.

## INTRODUCTION

Structural variants (SVs) are an important, yet under-studied type of human genetic variation. Numerous studies have implicated SVs (defined as >~50bp) with human diseases as well as normal phenotypic variation ^1–5^. Common SVs (MAF>1%) are enriched among loci identified in genome-wide association studies (GWAS) ^6^. In non-coding regions, SVs have a greater impact on gene expression compared to single nucleotide variants (SNVs) and small insertions and deletions (indels) ^7^. However, the discovery and genotyping of SVs is challenging and has lagged behind that of SNVs and indels. Many SVs are located within repetitive regions of the genome and often have complex internal structure ^6^. As a result, the contribution of SVs to the genetic architecture of complex traits remains poorly characterized.

The recent application of ensemble detection methods to whole genome sequencing (WGS) projects, particularly to large, multi-ancestry datasets, provides an opportunity to characterize the contribution of common and rare SVs to complex traits. Towards this end, we have utilized SVs detected from high-coverage WGS data from the NHLBI Trans-Omics for Precision Medicine (TOPMed) program and characterized their relationship to quantitative blood cell traits.

Red blood cell (RBC), white blood cell (WBC), and platelet laboratory parameters are routinely measured in clinical laboratories and used for monitoring general health status and diagnosis of acquired and inherited blood-related disorders. In the general population, hematologic quantitative traits are highly heritable and serve as a model system for studying the genetic architecture of complex traits ^8^. Thus far, hundreds of genomic loci and thousands of genetic variants have been associated with hematologic traits; however, these variants are almost exclusively SNVs and indels ^9,10^. For a few GWAS loci, there is evidence that common SVs are likely the causal variant responsible for the phenotypic effects in the population at large. For example, a common 3.7 kb alpha-globin gene deletion largely accounts for the strong association signal between RBC phenotypes and the 16p13.3 locus in African ancestry populations ^11–13^. While private SVs have been identified in individuals with rare Mendelian blood disorders (for example, a rare *PLAU* 78 kb tandem duplication responsible for autosomal dominant Quebec platelet disorder ^14^), the contribution of rare SVs (MAF<1%) to quantitative hematologic traits among unselected individuals has not been assessed.

In up to 50,675 ancestrally diverse TOPMed participants, we assessed the association of common and rare SVs (deletions, duplications and inversions) with variation in RBC, WBC and platelet-related quantitative traits. We characterized linkage disequilibrium patterns and conditional independence between blood cell trait-associated SVs and SNVs/indels previously associated with the same hematologic trait. Additionally, we used gene editing in monocytic and primary human HSPCs followed by xenotransplantation to demonstrate the mechanism by which a newly detected deletion disrupts an *S1PR3* monocyte enhancer and leads to decreased *S1PR3* expression and lower monocyte count.

## RESULTS

### Identification of common and rare SVs associated with blood cell traits

We performed single variant association tests, across 24 quantitative hematologic traits in up to 50,675 multi-ancestry TOPMed participants (**Table S1**). Single variant association tests were performed for SVs with a minor allele count (MAC) ≥ 5 (number of SVs=96,049). SVs in TOPMed were detected and genotyped from WGS using the Parliament2 pipeline^15^ and muCNV genotyper ^16^ (as described under Methods and in a separate manuscript). The QQ plots and genomic inflation factors (ranging from 0.981 to 1.056) were well-calibrated indicating good control of population stratification and relatedness (**Fig. S1 and Table S2**). Across the 24 hematologic traits, a total of 33 independent SVs (deletions=23, duplications=8, and inversions=2) were significant using the genome-wide significance threshold (P<5.7 × 10^−7^) (**Table 1 and Fig. S2**). Of these, 25 unique SVs (deletions=16, duplications=7, and inversions=2) reached a more stringent, experiment-wide significance threshold (P<5×10^−8^) (**Table 1 and Fig. S2**).

**Table 1.**
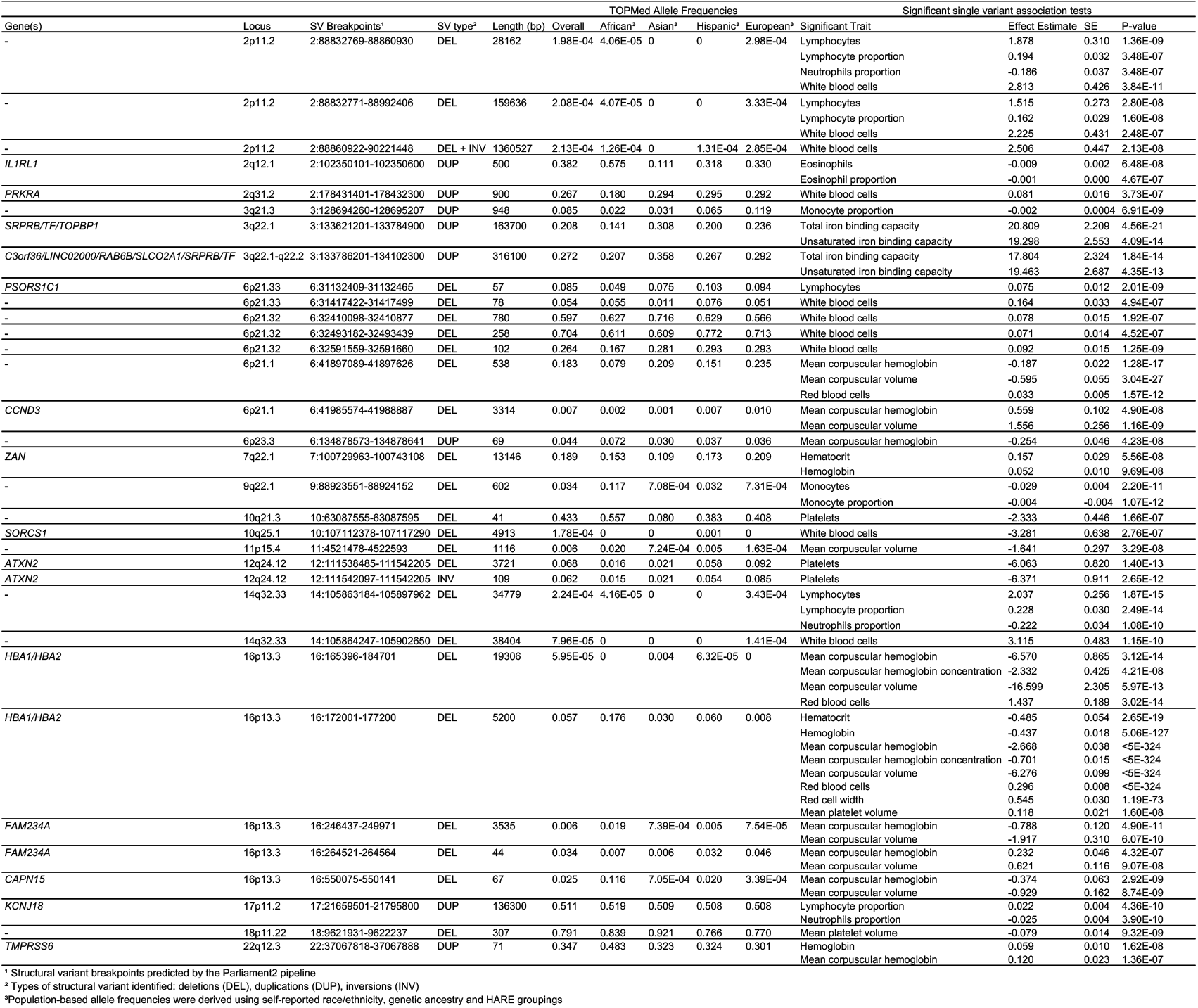
Summary of genome-wide structural variants associated with hematological traits.

The 33 trait-associated SVs ranged in size from ~40 bp to >160 kb (**Table 1**) and exhibited a range of allele frequencies: 23 are common (overall TOPMed MAF>1%) and 10 are rare (ranging from 0.006% to 0.7% MAF in TOPMed) with a few significant SVs exhibiting allele frequencies differences across populations (**Table 1**). For instance, the monocyte-associated deletion on chromosome 9q22.1 and a subset of the 16p13.3 red cell trait-associated SVs are more common in individuals of African ancestry than in those of non-African ancestry.

### Replication of significant SV-blood cell trait associations

We attempted replication for each of the 33 trait-associated SVs using a combination of short-read and long-read WGS data and genotype imputation. We utilized independent datasets composed of Icelandic (deCODE genetics)^17–19^ and multi-ancestry (UK Biobank, UKBB)^20^ participants. Note that the SV calling and genotyping algorithms used in replication datasets (described under Methods) are different from the Parliament2 pipeline used for SV discovery in TOPMed. To account for these methodological differences, we determined a set of “representative SVs” in deCODE genetics and UKBB datasets. We defined an SV in a replication dataset as “representative” if located within 5 kb of the trait-associated TOPMed SV and if the SV sizes overlapped by at least 25%.

Using these criteria, 4 of the 33 trait-associated SVs did not have an SV representative in deCODE or UKBB datasets, including SVs at 2q31.2 (2:178431401-178432300), 3q22.1 (3:133621201-133784900), 6p21.32 (6:32493182-32493439), and 17p11.2 (17:21659501-21795800) (**Table S3**). A total of 29 trait-associated SVs did have a representative and 27 of these were robustly replicated for the same blood cell trait (i.e. with a p-value < 0.05/number of its representative SVs in deCODE Icelandic, UKBB British, UKBB African, or UKBB South Asian cohorts and consistent direction of the effect) (**Table S3**). The two SVs that did not meet replication criteria (SVs at 2p11.2 [2:88832769-88860930] and 10q25.1 [10:107112378-107117290]) were rare and the 10q25.1 deletion had a p-value of 0.08 with consistent direction of effect.

### Trait-associated SVs in regions of LD with known GWAS loci

To determine if trait-associated SVs discovered in TOPMed are independent of previously reported GWAS SNVs/indels ^9,10,21–23^, we calculated pairwise linkage disequilibrium (LD) between TOPMed SVs and TOPMed SNVs/indels (**Table 2, Fig. S3)**. We also performed two sets of conditional analyses (see Methods). LD analysis shows 24 of 33 trait-associated SVs are in at least moderate LD (r2≥0.75) with one SNV/indel previously associated with the same hematologic trait (**Table 2, Fig. S3**). These include 9 SVs with at least one trait-associated SNV/indel in near perfect LD (r^2^≥0.99). Conditional regression analysis confirmed that these 24 SV association signals are markedly attenuated following adjustment for known SNV/indels at the same trait loci (**Table 3**), supporting the non-independence between SV and SNV/indel associations at these loci.

**Table 2.**
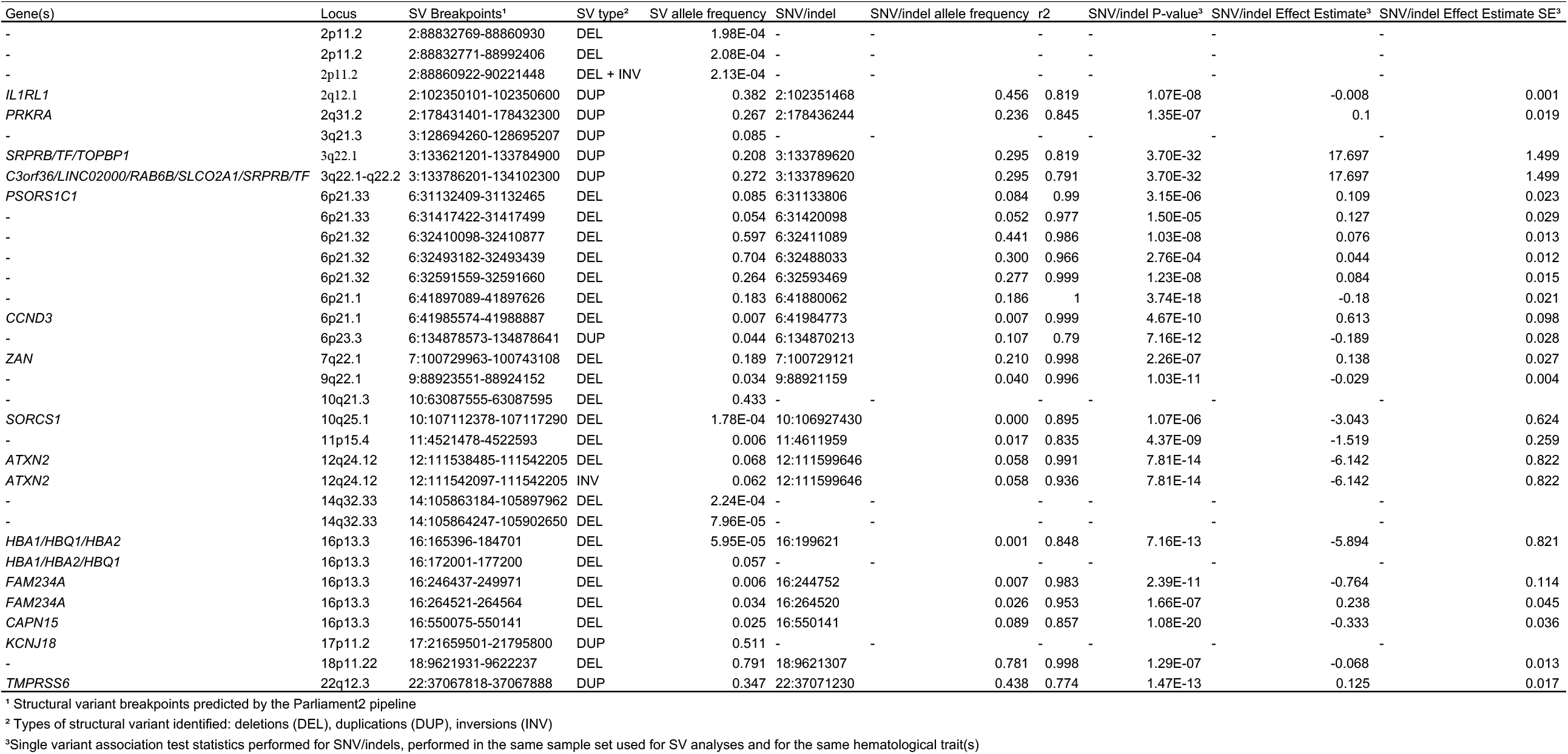
Single nucleotide variants (SNVs) and small insertions and deletions (indels) in linkage disequilibrium (r2≥0.75) with structural variants associated with hematological traits.

**Table 3.**
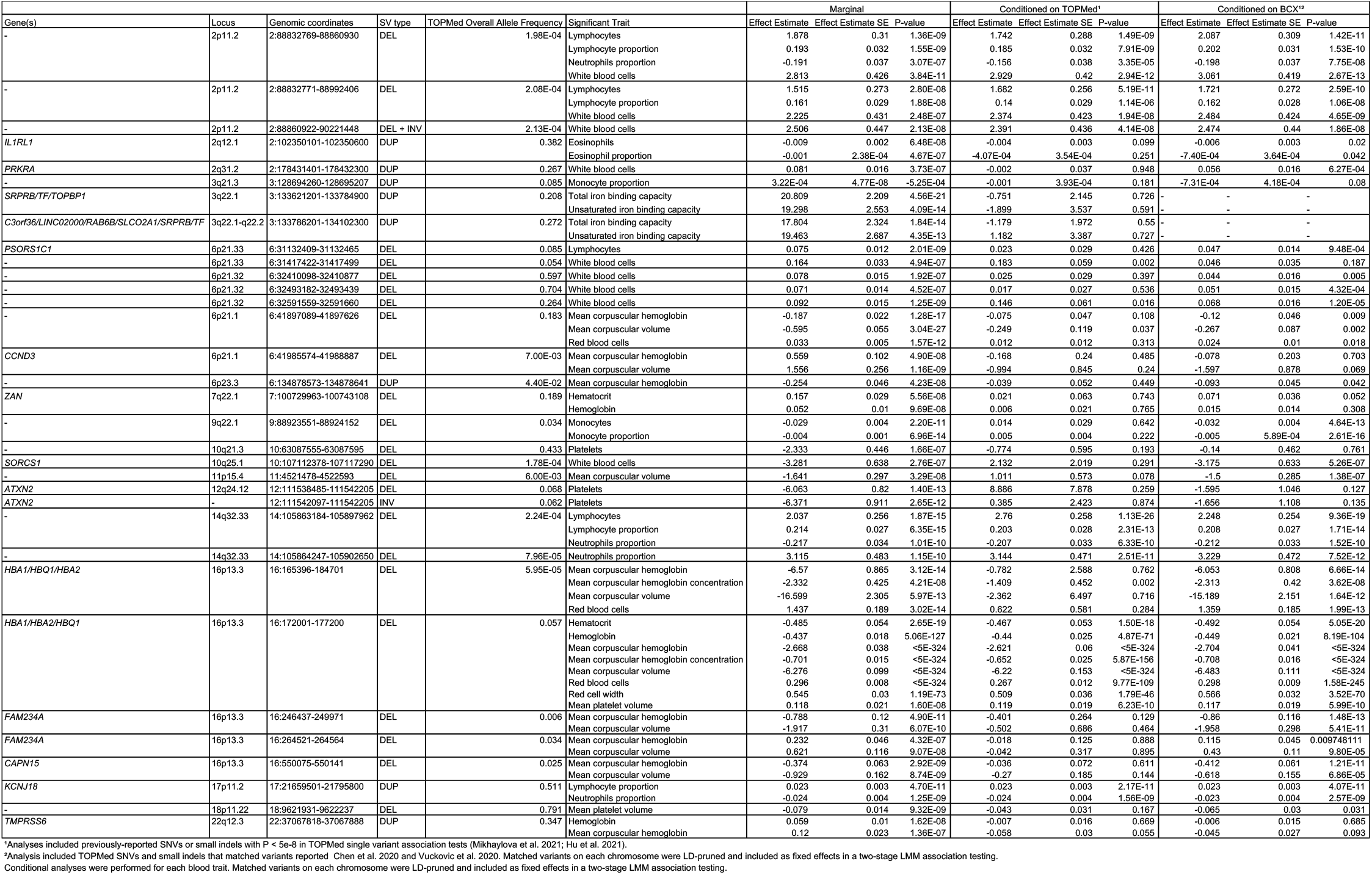
Structural variants conditioned on single nucleotide variants/small indels from previous genome-wide association studies.

### Trait-associated SVs conditionally independent of known GWAS SNVs/indels

A total of 7 trait-associated SVs remained genome-wide significant following conditional analyses (**Table 3**). This result suggests these association signals are independent from previously-reported GWAS SNVs/indels. These 7 SVs span 5 genomic loci. We discuss these 5 conditionally independent genomic loci in greater detail below.

#### 16p13.3 (alpha-globin) locus

The strongest association signal in our analyses was located at the 16p13.3 locus where a deletion spanning *HBA1/HBA2* (16:172001-177200) was associated with all 7 red cell traits (**Table 1, Fig. S4**). LD and conditional analyses indicate this deletion (16:172001-177200) is independent of other known red cell trait-associated SNVs/indels (**Table 2 and 3**). Although Parliament2 predicted this deletion as being 5.2 kb in size (**Table 1**), this deletion represents a previously characterized 3.7 kb alpha-globin deletion ^11^. This was confirmed by WGS read visualization in samples predicted to exhibit the *HBA1,HBA2* (16:172001-177200) deletion. Visualization shows SV breakpoints predicted by Parliament2 for this event are inaccurate and span the previously characterized 3.7 kb deletion (see example in **Fig. S4)**. The 3.7 kb alpha-globin deletion is known to be more common in African ancestry individuals ^21^. In our study, the overall allele frequency of the *HBA1/HBA2* deletion (16:172001-177200) was 5.7% and 17.6% in the African ancestry sub-population.

SV analyses also found the *HBA1/HBA2* deletion (16:172001-177200) as significantly associated with higher mean platelet volume (MPV) (**Table 1**). This was unexpected as none of the alpha-globin genes are known to regulate megakaryocyte or platelet production. While transcripts of genes located within the alpha-globin cluster on 16p13.3 are detectable in iPSC-induced megakaryocytes ^24^, we observed no evidence that this deletion is a *cis*-eQTL among African-ancestry individuals from the TOPMed GeneSTAR cohort (Bonferroni-corrected P-values >0.15 for all genes within a 1 Mb window). These observations are based on evidence from analysis of RNA from platelets (n=110) and iPSC-induced megakaryocytes (n=84) (data not shown). Relatedly, we found no association between the alpha-globin deletion (16:172001-177200) and circulating platelet counts in TOPMed (P=0.75).

In addition to the *HBA1/HBA2* deletion (16:172001-177200), analyses identified four other 16p13.3 deletions located within 500 kb of the alpha-globin gene cluster. LD and conditional analyses suggest these four deletions are not conditionally independent from trait-associated SNVs/indels in this region (**Table 2 and 3**). However, all four deletions showed a similar “thalassemia-like” pattern of red cell phenotypic association (lower MCH and MCV and higher RBC count) (**Table 1**) ^11^. These deletions range in size from ~40 bp to 19,000 bp. The ~19 kb deletion (allele frequency 0.006% in TOPMed overall and 0.3% in TOPMed Asian ancestry individuals) impacts both *HBA1* and *HBA2* and likely corresponds to the well-characterized alpha-thalassemia variant known as –(SEA) ^11^. The three other red cell trait-associated SVs on 16p13.3 are located ~75 to ~400 kb downstream of the alpha-globin genes and are not predicted to alter regions involved in alpha-globin gene regulation or show evidence by promoter Hi-C capture of physical interaction with globin gene promoters in blood cells (**Table S4**).

#### *17p11.2 (*KCNJ18*) locus*

A complex, multiallelic SV near the centromere of chromosome 17 (17p11.2) was significantly associated with higher lymphocyte proportions and lower neutrophil proportions (**Table 1, Fig. S4**). This SV is predicted to be a large duplication that includes the *KCNJ18* gene. Of note, the genomic region containing *KCNJ18* is not present in GRCh37; thus, this region was not interrogated in prior GRCh37 blood cell trait GWAS. In GRCh38, there is one copy of *KCNJ18*; however, based on Parliament2 SV calls, this region is likely duplicated (diploid copy number = 4) in most individuals (~87% of individuals in our TOPMed dataset). A subset of individuals (~2.7%) are estimated to have more than 4 diploid copies.

There is no LD between the *KCNJ18* SV and SNV/indels in the region (**Table 2**) and the SV-trait association is conditionally independent of known GWAS variants (**Table 3**). These results are consistent with a recent TOPMed WGS-based analysis, where no SNVs/indels in the 17p11.2 region were associated with WBC, neutrophil, or lymphocyte traits ^22^. However, given the phenotypic pattern (opposing effects on neutrophil and lymphocyte proportions) associated with the *KCNJ18* duplication, the complexity of the locus, the absence of a known role of *KCNJ18* in leukocyte biology, and the lack of detection in our replication cohorts (see above) additional work is needed to substantiate these results.

#### *2q31.2* (PRKRA) *locus*

A common duplication involving the 2q31.2 locus was significantly associated with higher WBC (**Table 1, Fig. S4**). This 2q31.2 duplication is predicted to span the coding region and 3’-UTR of the *PRKRA* gene. Investigation of the relationship between the 2q31.2 duplication and SNVs in the same genomic region showed the duplication is not in strong LD with SNVs/indels that passed QC metrics in TOPMed. However, this 2q31.2 duplication is in LD (r^2^=0.85) with a *PRKRA* SNV that failed QC metrics: chr2:178436244 (rs62176107) (**Table 2**).

Due to QC failure, rs62176107 was not previously tested for association with WBC traits in TOPMed ^22^. However, the WBC association signal for the 2q31.2 duplication was markedly attenuated (P=0.95) in conditional analyses with known TOPMed SNVs/indels (**Table 3**). Further analyses showed that signal attenuation in our conditional analysis is due to inter-chromosomal LD between the trait-associated 2q31.2 duplication and a known WBC trait-associated SNV^8^located in the HLA region (chr6:32623820, rs9271609, r^2^ = 0.945). Visualization of paired-end reads in samples exhibiting the 2q31.2 duplication show reads were aligned to the 2q31.2 region and two alternative HLA loci (chr6_GL000256v2_alt and chr6_GL000253v2_alt). Moreover, read-depth analysis of one of the alternative HLA loci shows alternative HLA loci copy number tracks very closely with chr2:178436244 (rs62176107) genotype (r^2^=0.85) (**Fig. S5**). Together these results strongly suggest the 2q31.2 SV is an insertion located in the HLA region, which may account for the association signal with WBC.

#### 2p11.2 and 14q32.33 immunoglobulin gene regions

Complex SVs at two loci, 2p11.2 and 14q32.33, were significantly associated with lymphocyte, neutrophil and WBC traits (**Table 1, Fig. S4**). These SV associations are conditionally independent of known WBC trait-associated SNV/indel (**Table 3**). SVs at both of these loci are rare and relatively large in size (**Table 1**). They are predicted to impact immunoglobulin kappa (2p11.2) and heavy chain (14q32.33) gene clusters. Based on their location and on visualization of WGS reads, these SVs likely represent somatic deletions and/or complex rearrangements due to V(D)J recombination events related to B cell maturation or immunoglobulin production^25^.

### Proportion of TOPMed SVs tagging known hematologic trait GWAS sentinel SNV/indels

To more broadly understand the extent to which SVs tag known, blood-cell trait SNVs/indels, we calculated LD for the genotypes of previously-reported SNVs/indels^10^ and the genotypes of SVs TOPMed participants. These analyses were performed in European ancestry samples (see Methods). Approximately 3% of previously-reported blood cell trait-associated SNVs/indels ^10^ (171 of the 6,652) were well-tagged (r^2^ > 0.8) by a TOPMed SV. For these 171 correlated pairs, we compared the trait-association p-values in TOPMed in an equivalent sample set of individuals with European ancestry. For most of the SNV/indel-SV pairs, the p-values were within an order of magnitude of each other (**Fig. S6**), indicating additional functional analyses are needed to identify the causal variant.

### Functional annotation of blood cell trait-associated SVs

Functional annotation can provide additional information to prioritize causal variants at trait-associated loci. Of the 33 trait-associated SVs, 7 SVs (4 duplications and 3 deletions) are predicted to overlap coding regions and thus potentially impact protein structure/function (**Table S4**). In addition to the SVs involving *KCNJ18* and *PRKRA* which are described above, two duplications spanning the transferrin gene (*TF*) coding and regulatory regions were associated with higher TIBC or transferrin levels (**Table 1**). In addition, three red cell phenotype-associated deletions are predicted to impact coding regions including: (1) the deletion encompassing the known 3.7 kb alpha-globin deletion which impacts the 3’ end of *HBA2* and 5’ end of *HBA1*; (2) the 19 kb deletion which comprises the SEA alpha-globin deletion and impacts both alpha-globin genes as well as *HBM* and *HBQ1*; and (3) a deletion which impacts the upstream and coding regions of the *ZAN* gene. This latter deletion also includes regulatory elements of the neighboring *EPO* gene, which encodes erythropoietin the major regulator of red cell production ^26^, and therefore likely represents the biologic mechanism by which this SV is associated with higher HGB/HCT levels.

Based on functional annotation, most trait-associated SVs (N=26) are predicted to only impact non-coding/regulatory genomic regions (intronic=10, intergenic=16) (**Table S4**). We cross-referenced SVs with candidate *cis* regulatory elements (cCREs) from ENCODE and several annotations relevant to 3D chromosome structure (frequently interacting regions or FIREs, topologically associating domains or TADs, super-interactive promoters or SIPs, and chromatin interactions ^27–32^). Annotation results show 14 trait-associated SVs overlapped cCREs, 18 overlapped with TAD boundaries, and 5 overlapped with FIREs in relevant tissues/cell-types (i.e., GM12878 and spleen) (**Table S4**). Two SVs overlapped SIPs in relevant cell-types. Chromatin interaction annotations from promoter capture Hi-C (pcHi-C) data ^29^ show that across 17 blood-cell-lineage cell types, 8 SVs overlap with the promoter regions of 19 genes. This includes deletions which overlap the promoter regions for the genes *EPO, HLA-DRB1* and a duplication which overlaps the promoter region of the *TF* gene. pcHi-C data also show 17 SVs overlap regions that interact with the promoters of 126 genes (**Table S4**). Similarly, monocyte Hi-C data^28^ show 27 SVs overlap potential regulatory regions interacting with promoters of 218 genes. Altogether, non-coding, functional annotations suggest most blood cell trait associated SVs may have an impact on transcriptional regulation.

### Fine-mapping and experimental validation of the 9q22.1 (*S1PR3*) monocyte locus

In cases where fine-mapping and functional evidence is similar between trait-associated SVs and correlated SNV/indels, further experimental follow-up may disentangle the causal variant. To illustrate this point, we performed experimental follow-up on a moderately sized deletion (602 bp) at the 9q22.1 locus. This deletion is near the *S1PR3* gene and was significantly associated with lower monocyte count and lower monocyte percentage (**Table 1**). This 9q22.1 deletion is also in near perfect LD with a recently reported monocyte-associated SNV (rs28450540) (**Table 2**) and several other SNVs, all of which are relatively specific to individuals of African ancestry (MAF=0.117).

To characterize the 9q22.1 locus, we compared the overlap between monocyte count-associated variants with deoxyribonuclease I (DNase I) sensitivity, an indicator of accessible chromatin. In several cell types, such as CD34^+^ common myeloid progenitor (CMP) cells and mesenchymal stem cells (MSCs), there was a relative absence of DNase I sensitivity adjacent to or overlying the 9q22.1 locus (**Fig. S7A**). However, in human primary CD14+ monocytes, several peaks of DNase I hypersensitivity overlap the monocyte-associated variants (**Fig. 1A**). Strikingly, the trait-associated 9q22.1 deletion (9:88923551-88924152) strictly overlapped a DNase I hypersensitivity peak, suggestive of regulatory potential (**Fig. 1A**). None of the SNVs with r^2^ > 0.8 with the 9q22.1 deletion directly overlapped DNase I peaks (**Fig. 1A**). In addition, sequences at the DNase I peak overlapping the 9q22.1 deletion showed histone modifications consistent with an enhancer signature in CD14^+^ monocytes, including the presence of H3K27ac and H3K4me1 and absence of H3K4me3 marks (**Fig. 1A**).

**Figure 1.**
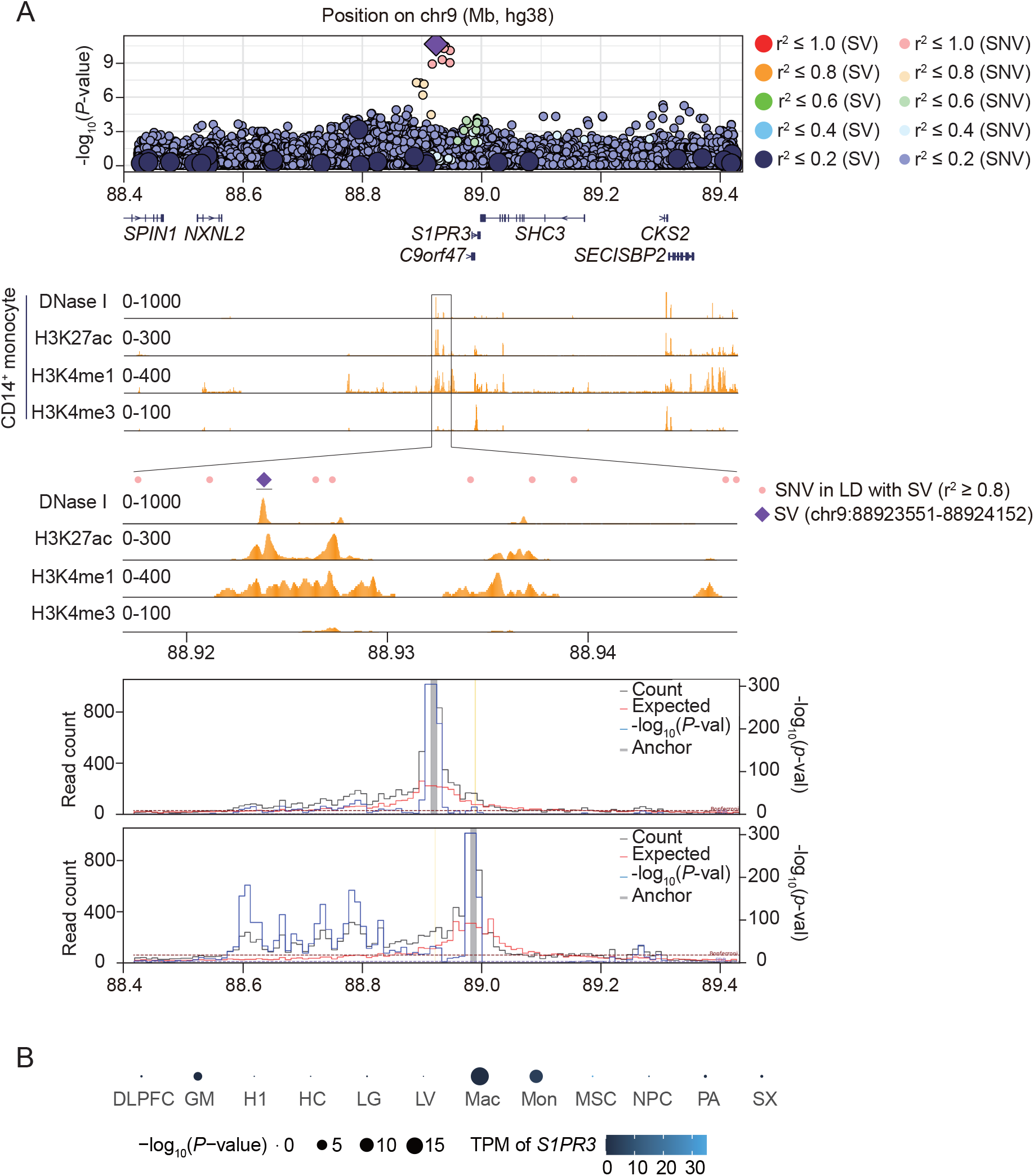
A structural variant at human 9q22.1 associated with decreased peripheral monocyte count. (A) Genome-wide association study summary statistics for 9q22.1 variants associated with peripheral monocyte counts. The purple diamond represents the trait-associated deletion (9:88923551-88924152); large circles represent other SVs; and small circles represent single nucleotide variants (SNVs) or indels. Color indicates the linkage disequilibrium (LD) calculated in the analysis sample set between the trait-associated deletion and individual SVs and SNVs. (B) Distribution of accessible chromatin (by DNase I sequencing) and histone modifications (H3K27ac, H3K4me1 and H3K4me3) in primary CD14+ monocytes across indicated genomic regions from ENCODE^27^. (C) Virtual 4C plot of long-range chromatin interactions anchored at the trait-associated, 9q22.1 deletion (9:88923551-88924152, upper panel) and the *S1PR3* promoter region (9:91605763-91606263, lower panel), shown as a grey bar, in macrophages. Yellow line highlights the *S1PR3* promoter region (upper panel) trait-associated, 9q22.1 deletion (lower panel). The observed and expected chromatin contact frequencies (or counts) are represented by the black and red lines, respectively. The left Y axis displays the range of chromatin contact frequency. The statistical significance (–log10(P-value)) of each long-range chromatin interaction is represented by the blue line, with its range listed in the right Y axis. The cell line or tissue specific FDR threshold (5%) is shown as a purple horizontal dashed line, and the more stringent Bonferroni threshold (P = 0.05) is shown as a maroon horizontal dashed line. (D) Long-range chromatin interaction between the trait-associated, 9q22.1 deletion and *S1PR3* promoter calculated in 12 different cell types. MSC (mesendoderm), NPC (neural progenitor cell), HC (hippocampus), H1 (human embryonic stem cells), LV (left ventricle), PA (pancreas), SX (spleen), DLPFC (dorsolateral prefrontal cortex), LG (lung) ^32^, GM (lymphoblast) ^75^, Mac (macrophages), Mon (monocytes) ^28^. Circle size represents the statistical significance, while the color indicates *S1PR3* mRNA level. TPM: transcripts per million.

A common feature of distal regulatory elements is long-range interaction with cognate promoters. We investigated these interactions from the viewpoint of the 9q22.1 SV using Hi-C data from monocytes and macrophages ^28^. We observed frequent interactions between the SV-deleted sequences and the *S1PR3* promoter, which is located in the same topologically associating domain (TAD) 67.3 kb downstream (**Fig. 1A**). Reciprocally, we investigated interactions from the viewpoint of the *S1PR3* promoter. The interactions between the *S1PR3* promoter and the 9q22.1 SV reached genome-wide significance in macrophage Hi-C data and were just below genome-wide significance in monocyte Hi-C data (**Fig. 1A** and **Fig. S7B**). In 10 other Hi-C datasets, including from cell types that express higher levels of *S1PR3* compared to monocytes or macrophages, such as MSCs, we did not observe significant interactions between the *S1PR3* promoter and the 9q22.1 deletion (**Fig. 1B**). These results suggest the trait-associated 9q22.1 SV overlaps a monocyte/macrophage-specific enhancer element that interacts with *S1PR3*.

Given this regulatory potential, we investigated whether the 9q22.1 SV was associated with expression changes of nearby genes. We performed expression quantitative trait loci (eQTL) analysis on the 9q22.1 SV in the TOPMed Multi-Ethnic Study of Atherosclerosis (MESA, using n = 169 AFHI participants including both African American and Hispanic/Latino individuals). *Cis*-eQTL analysis in CD14+ monocyte samples, revealed the strongest association to be between the 9q22.1 SV and *S1PR3* compared to all other genes in a 2 Mb window (**Fig. 2A**). Deletion of this region is significantly associated with decreased abundance of *S1PR3* (P = 5.20E-06) (**Fig. 2B**). Similar results were observed in peripheral blood mononuclear cells (PBMC) (**Fig. S8A**), but not in T cells, consistent with a cell type-specific cis-regulatory effect on *S1PR3* expression.

**Figure 2.**
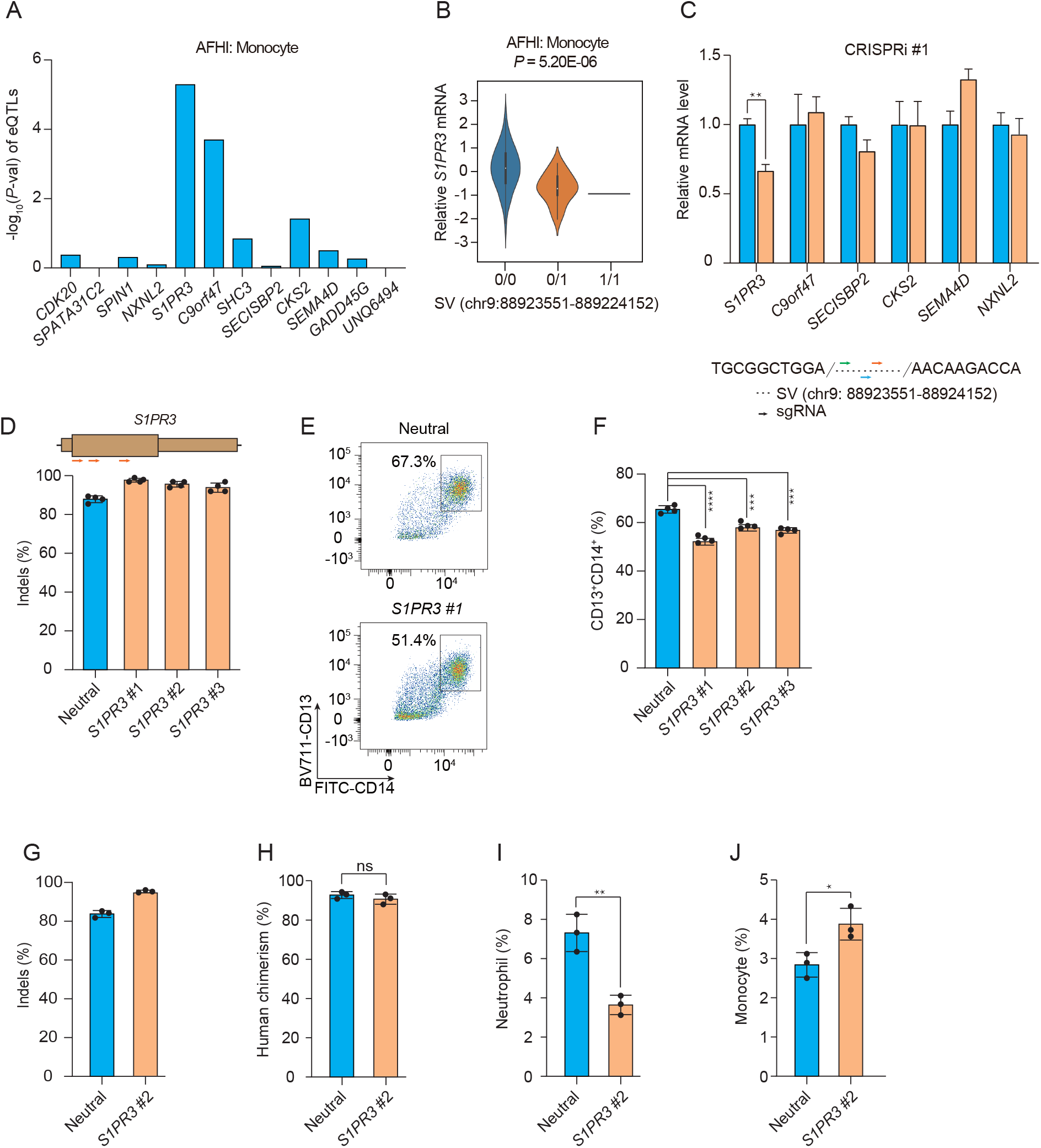
Genome and epigenome editing implicates *S1PR3* in the 9q22.1 monocyte association. (A) eQTL analysis between the 9q22.1 SV and genes within 2-Mb window in monocytes using data from the ancestry-stratified datasets from the Multi-Ethnic Study of Atherosclerosis (MESA, including n = 229 African American and n = 381 Hispanic/Latino participants). AFHI: African American and Hispanic/Latino. (B) eQTL analysis demonstrating the correlation between the 9q22.1 SV genotype and expression of S1PR3 in the ancestry-stratified datasets from the Multi-Ethnic Study of Atherosclerosis. (C) Expression of genes within a 2 Mb window in THP-1 cells after CRISPRi targeting the structural variant. Relative mRNA level of each gene represented by mean ± standard deviation (SD). N = 3 technical replicates. Location of three sgRNAs designed for CRISPRi are indicated below and **Figure S6C**. (D-F) *S1PR3* gene editing impaired monocyte differentiation in vitro. (D) Editing efficiency in HSPCs following 3xNLS-SpCas9:sgRNA electroporation with the indicated sgRNA. Gene edits were measured after 4 days of electroporation by Sanger sequencing (N = 4 biological replicates). Location of *S1PR3* coding sequence targeting sgRNAs are indicated above. (E) Representative flow cytometry indicating CD13+CD14+ cell populations from the neutral locus and *S1PR3* targeting group after 12-day differentiation. (F) CD13+CD14+ percentage in the *S1PR3* targeting group than the neutral locus targeting group. Mean ± standard deviation (SD), ***P < 0.001, ****P< 0.0001. N = 4 biological replicates. (G-J) Imbalanced myeloid differentiation due to *S1PR3* editing in bone marrow. Human CD34+ HSPCs from three healthy donors were edited by Cas9 RNP electroporation (EP) targeting a neutral locus and *S1PR3* coding sequence infused into NBSGW mice 24 h after electroporation. After 12 weeks, engrafted bone marrow was characterized by immunophenotyping. (G) Indels determined by Sanger sequencing before transplantation. (H-J) Quantification of different human cell types between the neutral locus and *S1PR3* targeting group. Human chimerism, hCD45+; Monocytes, hCD45+CD33+SSClowCD14+; neutrophil, hCD45+CD33+SSChighCD16+. Each dot represents one mouse. *P< 0.05, **P < 0.01.

To experimentally test the regulatory potential of the deleted sequences, we performed CRISPRi with dCas9-KRAB in monocytic THP-1 cells. Three sgRNAs were designed targeting different sequences within the 9q22.1 SV deleted segment (**Fig. S7C**). CRISPRi with each of these three sgRNAs significantly reduced the expression of *S1PR3* but not other nearby genes (**Fig. 2C** and **Fig. S7**). Taken together, the results provide strong evidence that the trait-associated SV deletes a monocyte-specific enhancer that controls the expression of *S1PR3* in monocytes.

To test the functional role of *S1PR3* in monocyte maturation and homeostasis, we edited human CD34+ hematopoietic stem and progenitor cells (HSPCs) with three sgRNAs targeting *S1PR3* or a sgRNA targeting a neutral locus and performed *in vitro* monocyte differentiation. Each of the *S1PR3*-targeting sgRNAs yielded highly efficient gene edits (95.7% ± 1.9% indels) (**Fig. 2D**). Compared with the neutral locus targeting control, each of the three *S1PR3-*edited cell populations showed a significant decrease in CD14+ monocyte differentiation efficiency *in vitro* (P<0.001) (**Fig. 2E-2F**), suggesting monocyte differentiation depends on *S1PR3* expression.

Lastly, to further validate the role of *S1PR3* in human hematopoiesis, we edited human hematopoietic stem and progenitor cells (HSPCs) with sgRNAs targeting a neutral locus or *S1PR3*, and infused the edited HSPCs into immunodeficient NBSGW mice. Human engraftment and multiple-lineage hematopoiesis were analyzed in the mouse bone marrow after 12 weeks. Gene edits were 95.4% in the input HSPC cell product for *S1PR3* and remained consistent (93.7%) in engrafting human cells (**Fig. 2G**). Overall human hematopoietic chimerism, and the fraction of lymphoid and erythroid lineage cells in the bone marrow was similar between the neutral locus and *S1PR3*-targeting group (**Fig. 2H** and **Fig. S9**). We observed a decrease of CD16+ neutrophil percentage (P = 0.004) and increase of CD14+ monocyte percentage (P = 0.02) in the bone marrow of *S1PR3*-edited groups (**Fig. 2I and 2J**). These results suggest that *S1PR3* loss of function leads to altered human myeloid homeostasis *in vivo*, consistent with a functional role of *S1PR3* in determining monocyte count.

## DISCUSSION

GWAS have identified thousands of SNVs and small indels that contribute to quantitative hematologic traits but the contribution of SVs to blood cell trait variation has mainly been limited to individuals with rare genetic blood disorders ^33–35^. Here we investigated the contribution of SVs to hematologic variation in ancestrally diverse TOPMed participants. Using single variant tests, we show 33 independent SVs were significantly associated with quantitative hematologic traits. These trait-associated SVs ranged in size (~40 bp to >160 kb) and allele frequency. Remarkably, most of these association signals were replicated in independent datasets, suggesting that despite the known challenges associated with SV discovery/genotyping in short-read data ^6^, WGS-based SV callsets can be successfully used to study complex trait variation.

Most trait-associated SVs are located in genomic regions previously associated with blood cell traits and most are not conditionally-independent of SNV/indels at the same loci previously identified through GWAS. One exception was a novel association between the 17p11.2 locus (*KCNJ18* SV) and white blood cell-related phenotypes. *KCNJ18* has no known role in leukocyte biology. It encodes a potassium channel and variants in this gene are associated with the Mendelian disorder, thyrotoxic hypokalemic periodic paralysis [MIM:613239]. Moreover, the trait-associated *KCNJ18* SV is located in a complex region of the genome which includes a segmental duplication near the centromere of chromosome 17. Likely due to this complexity, we did not identify a *KCNJ18* SV representative in replication datasets. Based on these results as well as the associated phenotypic pattern (opposing effects on neutrophil and lymphocyte proportions) additional analyses are required to ensure this finding reflects inherited genetic variation.

Functional annotation indicates most trait-associated SVs are located in non-coding regions of the genome. The majority of trait-associated SVs were predicted to impact regulatory elements or chromatin loop structure and chromatin domain boundaries. Together with LD and conditional analyses, this is consistent with the notion that SVs may provide mechanistic insights for a subset of known GWAS loci. For instance, in our analyses, a ~13 kb deletion on chromosome 7, which includes the *EPO* promoter, was associated HGB/HCT traits and near perfect LD with a previously-reported SNV chr7:100729121 (rs4729607)^9^. Recently, this chr7 deletion has been shown to alter iPSC expression levels of 5 nearby genes ^36^, including the genes, *TFR2* and *EPHB4* which are involved in iron metabolism and erythropoiesis ^37,38^ . In particular, this chr7 deletion is thought to modulate gene expression by impacting distal chromatin loop anchors ^36^.

In this study, we also identified a monocyte trait-associated SV (602 bp, 9q22.1 deletion) that directly overlaps a monocyte cell type-specific enhancer with accessible chromatin and enhancer signature histone modifications. The enhancer forms a physical interaction with the *S1PR3* gene in monocytes, and the 9q22.1 SV is an eQTL for *S1PR3* expression in monocytes. By using CRISPRi targeting the enhancer, we showed the enhancer positively regulates *S1PR3* in monocyte lineage cells. Prior GWAS have identified SNVs at this locus associated with blood cell traits including monocyte count, but without identification of the causal variant. This trait association represents an experimentally validated example where an ancestry-specific SV appears to underlie an SNV-tagged trait association through effects on cell-type specific gene regulation.

Gene editing of *S1PR3* significantly impacted both *in vitro* monocyte maturation and monocyte homeostasis in xenograft experiments. *S1PR3*, a receptor for the bioactive lipid sphingosine-1-phosphate (S1P), is a central regulator which drives myeloid differentiation^39^. Complementary to our results, previous studies have shown that *S1PR3* overexpression alone is sufficient to induce myeloid differentiation in human HSC^39^. The decreased efficiency of *in vitro* monocyte maturation, and relative increase in the fraction of bone marrow CD14^+^ monocytes with reciprocal decrease of bone marrow CD16^+^ neutrophils of engrafted mice, may suggest that *S1PR3* both plays cell autonomous roles in monocytes during maturation as well as impacts the trafficking of myeloid cells from bone marrow stores to circulating cells in the peripheral blood. Supporting this hypothesis, S1P receptors, which are chemotactically sensitive to S1P gradients, regulate multiple processes, including migration, matrix adhesion, and cell-cell contact. Therefore, the steep gradient of S1P concentration existing between bone marrow and blood, may drive cell types including monocytes, to navigate from the bone marrow to circulation^40^.

In summary, our results from a large ancestrally-diverse population-based data set add further evidence that complex trait association signals may be explained by the presence of structural variation. These findings complement recent WGS-based studies performed in European population isolates demonstrating the contribution of structural variation to complex trait variation (quantitative cardiometabolic and anthropometric traits)^5,17^. Several limitations of our study should be noted: 1) our analyses were restricted to deletions, inversions, and duplication and 2) were restricted to autosomal structural variation. Both of these limitations can be overcome with additional SV association studies that more broadly survey structural variation. In particular, the incorporation of long-read data into SV-based association analyses will greatly improve our understanding of how SVs contribute to hematological and complex trait variation.

## METHODS

### TOPMed study population

We included 50,675 participants from 12 TOPMed studies: Genetics of Cardiometabolic Health in the Amish (Amish, n=1,090)^41^, Atherosclerosis Risk in Communities Study (ARIC, n=3,717)^42^, Mount Sinai BioMe Biobank (BioMe, n=9,102)^43^, Coronary Artery Risk Development in Young Adults (CARDIA, n=2,966)^44^, Cardiovascular Health Study (CHS, n=3,478)^[45^ Genetic Epidemiology of COPD Study (COPDGene, n=5,595)^46^, Framingham Heart Study (FHS, n=2,760)^47^, Genetic Studies of Atherosclerosis Risk (GeneSTAR, n=1,494)^48^, Hispanic Community Health Study - Study of Latinos (HCHS_SOL, n=3,824)^49^, Jackson Heart Study (JHS, n=3,329)^50,51^, Multi-Ethnic Study of Atherosclerosis (MESA, n=2,516)^52^, and Women’s Health Initiative (WHI, n=10,804)^53^. The 50,675 TOPMed participants were categorized into discrete ancestry subgroups using the HARE (harmonized ancestry and race/ethnicity) machine learning algorithm, which uses genetically inferred ancestry to refine self-identified race/ethnicity and impute missing racial/ethnic values^54^ (see **Supplemental Methods)**. The ancestry composition in this study was 59% European, 24% African, 16% Hispanic/Latino, and 1% Asian. (**see Table S1**). Only samples with a missingness rate <10% in the structural variant dataset were included in analysis. Further descriptions of the design of the participating TOPMed cohorts and the sampling of individuals within each cohort for TOPMed WGS are provided in the section “Participating TOPMed studies” under **Supplemental Methods**. All studies were approved by the appropriate institutional review boards (IRBs) and informed consent was obtained from all participants.

### Blood cell trait measurements

Red blood cell, white blood cell and platelet quantitative traits were measured from freshly collected whole blood samples using automated hematology analyzers according to clinical laboratory standards. In studies where multiple blood cell measurements per participant were available, we selected a single measurement for each trait and each participant. Each trait was defined as follows: Hematocrit (HCT) is the percentage of volume of blood that is composed of red blood cells. Hemoglobin (HGB) is the mass per volume (grams per deciliter) of hemoglobin in the blood. Mean corpuscular hemoglobin (MCH) is the average mass in picograms of hemoglobin per red blood cell. Mean corpuscular hemoglobin concentration (MCHC) is the average mass concentration (grams per deciliter) of hemoglobin per red blood cell. Mean corpuscular volume (MCV) is the average volume of red blood cells, measured in femtoliters (fL). RBC count is the count of red blood cells in the blood, by number concentration in millions per microliter. Red cell distribution width (RDW) is the measurement of the ratio of variation in width to the mean width of the red blood cell volume distribution curve taken at +/- one CV. Total white blood cell count (WBC), neutrophil, monocyte, lymphocyte, eosinophil, basophil and platelet count are defined with respect to cell concentration in blood, measured in thousands/microliter. The proportion of neutrophils, monocytes, lymphocytes, or eosinophils were calculated by dividing the respective WBC sub-type count by the total measured WBC. Mean platelet volume (MPV) was measured in fL. For each trait, we identified extreme values that may represent measurement or recording errors or hematologic malignancies and removed them from the analysis.

In a subset of samples from the JHS and HCHS/SOL studies, we evaluated four iron-related phenotypes: serum iron, total iron binding capacity (TIBC), transferrin saturation, and ferritin. Serum iron (μg/dl) was measured by colorimetric assay using a ferrozine reagent (Roche Diagnostics, Indianapolis, IN). Unsaturated iron binding capacity (UIBC) was assayed by colorimetric assay on the same sample, TIBC (μg/dl) was calculated by the formula: TIBC = serum iron + UIBC. Serum ferritin (ng/ml) was measured with Roche reagents using a particle enhanced immunoturbidimetric assay. Transferrin saturation (%) was calculated by the formula: SAT = serum iron/TIBC x 100.

### WGS data and quality control in TOPMed

WGS was performed through the NHLBI TOPMed program on genomic DNA isolated from peripheral blood. WGS was generated to an average depth of 38X by six sequencing centers (Broad Genomics, Northwest Genomics Center, Illumina, New York Genome Center, Baylor, and McDonnell Genome Institute)^55^. Most WGS was performed using PCR-free library preparation, Illumina HiSeq X Ten or NovaSeq instruments and 150bp paired end reads. Sequencing reads were aligned to the human reference genome (GRCh38) by the TOPMed Informatics Research Center (IRC) using the read mapping pipeline described in Regier, A. et al. 2018^56^.

### Single nucleotide variant and small indel discovery and genotyping in TOPMed

We utilized the TOPMed freeze 8 genotype call set produced by the IRC as previously described ^56^. Briefly, SNVs and indels were discovered on a per sample basis, then merged and genotyped across samples. SNV and indel quality control (QC), was performed by calculating Mendelian consistency scores and by applying a support vector machine (SVM) classifier trained on known variant sites and Mendelian inconsistencies. SNV- and indel-based, sample QC measures included: concordance between annotated and inferred genetic sex, concordance between prior array genotype data and TOPMed WGS data, and pedigree checks. Further details regarding data processing, and quality control are described on the TOPMed website (https://www.nhlbiwgs.org/topmed-whole-genome-sequencing-methods-freeze-8) and in a common document accompanying each TOPMed study’s dbGaP accession.

### Structural variant discovery and genotyping in TOPMed

We utilized the TOPMed SV freeze 1.0 call set, which contains 138,134 TOPMed samples. The details of SV discovery and genotyping in TOPMed are reported in a separate manuscript. Briefly, SV calls were assessed from each sample separately using Parliament2 pipeline ^15^. The Parliament2 pipeline provides the union of SV calls from six different programs: BreakDancer, BreakSeq, CNVnator, Delly, Lumpy and Manta. SV calls were merged across samples using survivor ^57^ and filtered using SVTyper^58^. Sample genotypes for each variant were assessed using muCNV ^16^. After final filtering, the TOPMed SV freeze 1.0 call set consists of a total of 466,455 autosomal SV sites: 231,817 deletions, 197,412 duplications and 37,226 inversions. Of these, 96,049 had MAC >= 5 in at least one trait and were included in association analyses. For association analysis, the genotypes of each SV were represented in a bi-allelic genotype format (GT = 0/0, 0/1, 1/1), similar to SNVs and small indels generated from the same WGS data.

### Single variant association analysis of SVs and blood cell traits using linear mixed models

Single variant SV association tests for all variants with a minor allele count (MAC) ≥ 5 were performed for each blood cell trait using a two-stage linear mixed model (LMM) approach implemented in the GENESIS software ^59,60^. In the first stage, a null model assuming no association between the outcome and any SV was fit, adjusting for the fixed effect covariates of: age at trait measurement; sex; a variable indicating TOPMed study and study phase (study_phase); indicators for stroke, COPD, and VTE; the first 11 PC-AiR^61^ principal components (PCs) of genetic ancestry as estimated from the WGS SNV/indel genotypes. We additionally included as fixed effect covariates, the first 10 principal components estimated from read depth (“batch PCs”). To calculate batch PCs, we first computed the average sequencing depth for every 1 kb genomic region (“bin”) across the 22 autosomes ^62^. We removed bins containing repetitive sequences with poor mappability (<1.0 using 50bp k-mers in GEMTools v1.759) or sequences overlapping known CNVs in the Database of Genomic Variants. Following normalization of the approximately 150,000 remaining bins, we performed Randomized Singular Value Decomposition (rSVD)^63^, to generate batch PCs, which were used to correct for batch and technical artifacts arising from the sequencing process.

In the first-stage null model, a 4th degree sparse empirical kinship matrix (KM) computed with PC-Relate^64^ was included to account for genetic relatedness among participants. To control genomic inflation, we additionally allowed for heterogeneous residual variances by study and ancestry group. Details on how ancestry groups were estimated for this adjustment are in the supplemental methods. Following fitting of the first-stage null model, we performed a rank-based inverse-normal transformation of the marginal residuals, and subsequently rescaled the residuals by their variance prior to transformation. This rescaling allows for clearer interpretation of estimated SV genotypic effect sizes from the subsequent association tests. In the second stage, we fit another LMM using the rank-normalized and rescaled residuals as the outcome, with the same fixed effect covariates, sparse KM, and heterogeneous residual variance model as in Stage 1. The output of the Stage 2 null model was then used to perform genome-wide score tests of genetic association for all SVs that passed the TOPMed SV quality filters and with a minor allele count (MAC) ≥ 5. Missing SV genotype calls were imputed to the mean before performing the association tests. The total number of unique SVs tested across all traits was 96,049.

Basophils was tested as a binary outcome (basophil count > 0), so the null model was fit as a logistic mixed model using the GMMAT method as implemented in GENESIS, rather than a two-stage LMM. The same fixed effect covariates and sparse KM were used in the null model, and score tests were used for association.

### Effective number of independent association tests and significance threshold

For single variant SV analyses, we used the simpleM method ^65^ to estimate the effective number of independent tests (Meff) as 87,547, which corresponds to a genome-wide significance threshold of 5.7 × 10^−7^ after Bonferroni correction. To further correct for testing multiple phenotypes ^66^, we used an experiment-wide significance threshold of 5 × 10^−8^.

### Visualization of SVs associated with blood cell traits

For SVs significantly associated with blood cell traits, we performed additional quality control by visualizing aligned WGS reads in variant samples. Visualization was performed using samplot on the NHLBI Biodata Catalyst cloud computing platform (https://doi.org/10.5281/zenodo.3822858). For each SV event, we visualized aligned reads for multiple samples and excluded any SV events that were not clearly supported by the aligned WGS data. Additionally, we selected SV events in instances where Parliament2 identified multiple overlapping SVs by different SV calling algorithms. This was concluded following data visualization and we selected SVs based on the resolution of predicted Parliament2 breakpoints.

### Replication of trait-associated SVs using deCODE genetics and UK Biobank datasets

We performed replication analyses for each TOPMed SV-blood cell trait association signal using deCODE genetics and UKBB datasets. Briefly, SVs were called in Icelanders (deCODE genetics) using 49,962 short-read ^18,19^ and 3,622 long-read sequenced ^17^ individuals. These data were phased and all genotyped variants were imputed into 166,281 individuals using a previously described methodology ^67,68^. SVs were called from 150,119 short-read sequenced individuals in UKBB. Three cohorts were used in UKBB with 132,169, 2,963, and 3,047 sequenced and 431,805, 9,633, and 9,252 imputed individuals, in British/Irish (XBI), African (XAF) and South Asian (XSA) populations, respectively ^20^. For replication analyses, we defined an SV in replication datasets to represent a TOPMed SV, if located within 5 kb of the TOPMed SV and if the two SV sizes overlapped by at least 25%. We tested for association for all representative SVs and their corresponding phenotypes based on the linear mixed model implemented in BOLT-LMM ^69^ and described in ^17,20^. We considered an association to be replicated if at least one of the p-values from the deCODE, UKBB British, UKBB African, or UKBB South Asian cohorts was < 0.05/(number of its representative SVs in given dataset).

### Functional annotation of SVs

We annotated genome sequence information for SVs significantly associated with blood cell traits using AnnoSV^70^. From AnnoSV, we ascertained gene annotations (based on RefSeq, ENSEMBL), the presence of similar SVs in genomic databases (e.g., DGV) and breakpoint information including overlap with repetitive elements. To understand the potential impact of trait-associated SVs on non-coding/regulatory regions, we cross-referenced SVs with five different genomic annotations including, frequently interacting regions (FIREs) from Hi-C data ^31,32^, topologically associating domain (TAD) boundaries, cell-type specific regulatory networks from super interactive promoters (SIPs) ^30^, candidate *cis* regulatory elements (cCREs) from ENCODE, and chromatin interaction information from chromatin conformation data including Hi-C ^28^ and promoter capture Hi-C (pcHi-C) ^29^.

### Linkage disequilibrium and conditional analyses for trait-associated SVs

For each blood cell trait, we performed conditional association analyses to determine which genome-wide significant SVs were (1) independent of previously reported GWAS variants and (2) independent of SNV and small indels previously detected in TOPMed ^21–23^. To address the first question, we used variants detected in multi-ancestry and European populations reported in Chen et al. 2020 and Vuckovic et al. 2020 ^9,10^. The genome-wide significant variants from Chen et al. 2020 and Vuckovic et al. 2020 were matched to TOPMed SNVs and small indels that passed the IRC quality filters based on chromosome, position, and alleles. For each trait, the set of matched variants on each chromosome was then LD-pruned at r^2^ = 0.8 in the sample set for that trait, preferentially keeping variants with more significant p-values in the TOPMed analysis sample set. This pruned set of variants were combined across chromosomes and included as fixed effect covariates in the null model using the same fully-adjusted two stage LMM association testing procedure described above ^59,60^.To identify SVs independent of GWAS variants detected in the TOPMed data, we used a similar procedure, starting with any SNV or small indels with P < 5e-8 in single variant association tests using the same sample set for the trait. This set of variants was then LD-pruned at r^2 = 0.8, again preferentially keeping variants with more significant p-values, and included as fixed effects in a two-stage LMM association testing ^59,60^.

To investigate the proportion of SVs in LD with known, blood-cell trait SNVs/indels, we additionally calculated LD (r^2^) for genotypes of 6,652 previously-reported SNVs/indels from European ancestry samples^10^ and SVs with MAC ≥ 5 on the same chromosome. Only TOPMed participants with inferred European ancestry (N = 29,244) were used for the LD calculation. Each SNV/indel was then matched to the SV with the highest r^2^ value.

### Hematopoietic cell lines

THP-1 cells (Cat# TIB-202) were obtained from the American Type Culture Collection and cultured in RPMI 1640 (Thermo Fisher Scientific, USA). To make the complete growth medium, the following components were added: 2-mercaptoethanol (Cat# 21985-023, Thermo Fisher Scientific) to a final concentration of 0.05 mM; fetal bovine serum (Cytiva) to a final concentration of 10%.

### Primary hematopoietic cells and monocyte-macrophage differentiation

Human CD34^+^ HSPCs from mobilized peripheral blood of deidentified healthy donors were purchased from Fred Hutchinson Cancer Research Center, Seattle, Washington. CD34^+^ HSPCs were cultured in StemSpan SFEM medium (Cat# 09650, STEMCELL Technologies) supplemented with 1x StemSpan CD34^+^ expansion supplement (Cat# 02691, STEMCELL Technology). To induce monocyte-macrophage differentiation from CD34^+^ HSPCs, the cytokine cocktail of M-CSF 30 ng/mL, FLT3-Ligand 100 ng/mL, SCF 50 ng/ml, IL-3 5 ng/mL, IL-6 3 ng/ul and L-Glutamine 2 mM was supplemented to the culture media for 11 days before analysis. GM-CSF 5 ng/ml was supplemented in the culture media for the first 4 days. All cytokines of human origin and from PeproTech.

### CRISPR/Cas9 guide design, cloning, lentiviral vector production and transduction and 3xNLS-SpCas9 preparation

Streptococcus pyogenes Cas9 (SpCas9) guide RNAs that either target *S1PR3* coding sequence or bind near the structural deletion (9:88923551-8892452) were identified using computational algorithms with prioritization for on-target efficiency and reduced off-target effects (CRISPOR: http://crispor.tefor.net/). For RNP experiments, the chemically modified sgRNAs were synthesized by Integrated DNA Technologies. SpCas9 proteins were expressed and purified as previously described^71^.

For CRISPRi experiments, oligos (from GENEWIZ company) were annealed and ligated into LentiGuide-Puro (Addgene, Cat#52963). Following lentiviral production and transduction into THP-1 cell lines with stable dCas9-KRAB expression (Addgene, Cat#89567), 10 μg/ml blasticidin and 1 μg/ml puromycin were added to select for sgRNA expression in cells with stable dCas9-KRAB expression.

The sequence of sgRNAs are summarized in **Table S5**.

### CRISPR-Cas9 genome editing in CD34^+^ HSPCs and THP-1 cells

CD34^+^ HSPCs and THP-1 cells were maintained in their favorable medium (see before) 24 h before electroporation. Approximately 100,000 cells per condition were electroporated using the Lonza 4D nucleofector with 100 pmol 3xNLS-SpCas9 protein and 300 pmol modified sgRNA targeting the locus of interest. In addition to mock treated cells, “safe-targeting” RNPs were used as experimental controls as indicated in each figure legend^72^. After electroporation, cells were induced for monocyte-macrophage differentiation as described previously. Genomic DNA was isolated from an aliquot of cells, the sgRNA targeted locus was amplified by PCR which was subject to Sanger sequencing and then TIDE analysis to quantify indel spectrum at day 4 after electroporation.

### Determination of target gene expression

Total RNA was extracted from cell cultures 4 days after electroporation using the RNeasy Plus Mini Kit (QIAGEN) and reverse transcribed using the iScript cDNA synthesis kit (Biorad) according to the manufacturer’s instructions. Expression of target genes was quantified using real-time RT-qPCR with GAPDH as an internal control. All gene expression data represent the mean of at least three biological replicates. Primers for PCR are summarized in **Table S6**.

### Flow cytometry analysis

For analysis of surface markers, cells were stained in PBS containing 2% (w/v) BSA, with anti-human CD34 (581), anti-human CD33 (P67.6), anti-human CD117 (104D2), anti-human CD16 (3G8), anti-human CD14 (63D3). Monocyte and macrophage were indicated by anti-human CD14^+^. Flow cytometry data were acquired on a LSRII or LSR Fortessa (BD Biosciences) and analyzed using FlowJo software (Tree Star).

### Immunophenotyping of human CD34^+^ HSPCs xenograft from NBSGW mice

NOD.Cg-KitW-41J Tyr Þ Prkdcscid Il2rgtm1Wjl (NBSGW) mice were obtained from Jackson Laboratory (Stock 026622). CD34^+^ HSPCs were maintained and edited as described above. Cells were allowed to recover for 24 h after electroporation, and then infused by retro-orbital injection into non-irradiated NBSGW female mice. Bone marrow was isolated for human xenograft analysis 16 weeks after human CD34^+^ HSPCs engraftment. For flow cytometry analysis, bone marrow cells were first incubated with Human TruStainFcX (422302, BioLegend) and TruStainfcX (anti-mouse CD16/32, 101320, BioLegend) blocking antibodies for 10 min and then stained with marker panels designed for multi-lineage analysis, including anti-mouse CD45 (30-F11), anti-human CD45 (HI30), anti-human CD34 (581), anti-human CD33 (P67.6), anti-human CD117 (104D2), anti-human CD16 (3G8), anti-human CD14 (63D3), anti-human CD235a (HI264), anti-human CD3 (UCHT1), anti-human CD19 (HIB19), and Fixable Viability Dye eFluor 780 for live/dead staining (65-0865-14, Thermo Fisher). Percentage human engraftment was calculated as hCD45^+^ cells/(hCD45^+^ + mCD45^+^ cells).

### DNase I-sequencing, chromatin and Hi-C datasets

DNase I-sequencing and histone modification datasets, including H3K27ac, H3K4me1 and H3K4me3, were downloaded from the ENCODE project ^27^, and were then analyzed using WashU Epigenome Browser online software ^73^. In situ Hi-C maps of DNA interactions in human monocytes and macrophages were previously described ^28^, and visualized by HUGIn2 (Hi-C Unifying Genomic Interrogator, http://hugin2.genetics.unc.edu/Project/hugin/)^74^.

## Data Availability

All data produced in the present study are available upon reasonable request to the authors.
TOPMed genomic data and pre-existing Parent study phenotypic data are made available to the scientific community in study-specific accessions in the database of Genotypes and Phenotypes (dbGaP) and in the NHLBI BioData Catalyst cloud platform.

https://www.ncbi.nlm.nih.gov/gap/advanced_search/?TERM=topmed

## ACKNOWLEDGEMENTS

Molecular data for the Trans-Omics in Precision Medicine (TOPMed) program was supported by the National Heart, Lung and Blood Institute (NHLBI). See the TOPMed Omics Support Table (**Table S7**) for study specific omics support information. Core support including centralized genomic read mapping and genotype calling, along with variant quality metrics and filtering were provided by the TOPMed Informatics Research Center (3R01HL-117626-02S1; contract HHSN268201800002I). Core support including phenotype harmonization, data management, sample-identity QC, and general program coordination were provided by the TOPMed Data Coordinating Center (R01HL-120393; U01HL-120393; contract HHSN268201800001I). We gratefully acknowledge the studies and participants who provided biological samples and data for TOPMed.

Genetics of Cardiometabolic Health in the Amish (Amish)

The TOPMed component of the Amish Research Program was supported by NIH grants R01 HL121007, U01 HL072515, and R01 AG18728.

Atherosclerosis Risk in Communities Study (ARIC) study

Whole genome sequencing (WGS) for the Trans-Omics in Precision Medicine (TOPMed) program was supported by the National Heart, Lung and Blood Institute (NHLBI). WGS for “NHLBI TOPMed: Atherosclerosis Risk in Communities (ARIC)” (phs001211) was performed at the Baylor College of Medicine Human Genome Sequencing Center (HHSN268201500015C and 3U54HG003273-12S2) and the Broad Institute for MIT and Harvard (3R01HL092577-06S1). Centralized read mapping and genotype calling, along with variant quality metrics and filtering were provided by the TOPMed Informatics Research Center (3R01HL-117626-02S1). Phenotype harmonization, data management, sample-identity QC, and general study coordination, were provided by the TOPMed Data Coordinating Center (3R01HL-120393-02S1). We gratefully acknowledge the studies and participants who provided biological samples and data for TOPMed.

The Genome Sequencing Program (GSP) was funded by the National Human Genome Research Institute (NHGRI), the National Heart, Lung, and Blood Institute (NHLBI), and the National Eye Institute (NEI). The GSP Coordinating Center (U24 HG008956) contributed to cross-program scientific initiatives and provided logistical and general study coordination. The Centers for Common Disease Genomics (CCDG) program was supported by NHGRI and NHLBI, and whole genome sequencing was performed at the Baylor College of Medicine Human Genome Sequencing Center (UM1 HG008898 and R01HL059367).

The Atherosclerosis Risk in Communities study has been funded in whole or in part with Federal funds from the National Heart, Lung, and Blood Institute, National Institutes of Health, Department of Health and Human Services (contract numbers HHSN268201700001I, HHSN268201700002I, HHSN268201700003I, HHSN268201700004I and HHSN268201700005I). The authors thank the staff and participants of the ARIC study for their important contributions.

Mount Sinai BioMe Biobank (BioMe)

The Mount Sinai BioMe Biobank has been supported by The Andrea and Charles Bronfman Philanthropies and in part by Federal funds from the NHLBI and NHGRI (U01HG00638001; U01HG007417; X01HL134588). We thank all participants in the Mount Sinai Biobank. We also thank all our recruiters who have assisted and continue to assist in data collection and management and are grateful for the computational resources and staff expertise provided by Scientific Computing at the Icahn School of Medicine at Mount Sinai.

Coronary Artery Risk Development in Young Adults (CARDIA)

The Coronary Artery Risk Development in Young Adults Study (CARDIA) is conducted and supported by the National Heart, Lung, and Blood Institute (NHLBI) in collaboration with the University of Alabama at Birmingham (HHSN268201800005I & HHSN268201800007I), Northwestern University (HHSN268201800003I), University of Minnesota (HHSN268201800006I), and Kaiser Foundation Research Institute (HHSN268201800004I). CARDIA was also partially supported by the Intramural Research Program of the National Institute on Aging (NIA) and an intra-agency agreement between NIA and NHLBI (AG0005).

Cardiovascular Health Study (CHS)

Cardiovascular Health Study: This research was supported by contracts HHSN268201200036C, HHSN268200800007C, HHSN268201800001C, N01HC55222, N01HC85079, N01HC85080, N01HC85081, N01HC85082, N01HC85083, N01HC85086, 75N92021D00006, and grants U01HL080295 and U01HL130114 from the National Heart, Lung, and Blood Institute (NHLBI), with additional contribution from the National Institute of Neurological Disorders and Stroke (NINDS). Additional support was provided by R01AG023629 from the National Institute on Aging (NIA). A full list of principal CHS investigators and institutions can be found at CHS-NHLBI.org. The content is solely the responsibility of the authors and does not necessarily represent the official views of the National Institutes of Health.

Genetic Epidemiology of COPD Study (COPDGene)

The COPDGene project described was supported by Award Number U01 HL089897 and Award Number U01 HL089856 from the National Heart, Lung, and Blood Institute. The content is solely the responsibility of the authors and does not necessarily represent the official views of the National Heart, Lung, and Blood Institute or the National Institutes of Health. The COPDGene project is also supported by the COPD Foundation through contributions made to an Industry Advisory Board comprised of AstraZeneca, Boehringer Ingelheim, GlaxoSmithKline, Novartis, Pfizer, Siemens and Sunovion. A full listing of COPDGene investigators can be found at: http://www.copdgene.org/directory

Framingham Heart Study (FHS)

The Framingham Heart Study (FHS) acknowledges the support of contracts NO1-HC-25195, HHSN268201500001I and 75N92019D00031 from the National Heart, Lung and Blood Institute and grant supplement R01 HL092577-06S1 for this research. We also acknowledge the dedication of the FHS study participants without whom this research would not be possible. Dr. Vasan is supported in part by the Evans Medical Foundation and the Jay and Louis Coffman Endowment from the Department of Medicine, Boston University School of Medicine.

Genetic Studies of Atherosclerosis Risk (GeneSTAR)

WGS for “NHLBI TOPMed: GeneSTAR (Genetic Study of Atherosclerosis Risk)” (phs001218) was performed at the Broad Institute of MIT and Harvard (HHSN268201500014C), at PsomaGen (formerly Macrogen, HHSN268201500014C), and at Illumina (HL112064).

GeneSTAR was supported by the National Institutes of Health/National Heart, Lung, and Blood Institute (U01 HL72518, HL087698, HL112064) and by a grant from the National Institutes of Health/National Center for Research Resources (M01-RR000052) to the Johns Hopkins General Clinical Research Center.

Hispanic Community Health Study - Study of Latinos (HCHS_SOL)

The Hispanic Community Health Study/Study of Latinos is a collaborative study supported by contracts from the National Heart, Lung, and Blood Institute (NHLBI) to the University of North Carolina (HHSN268201300001I / N01-HC-65233), University of Miami (HHSN268201300004I / N01-HC-65234), Albert Einstein College of Medicine (HHSN268201300002I / N01-HC-65235), University of Illinois at Chicago – HHSN268201300003I / N01-HC-65236 Northwestern Univ), and San Diego State University (HHSN268201300005I / N01-HC-65237). The following Institutes/Centers/Offices have contributed to the HCHS/SOL through a transfer of funds to the NHLBI: National Institute on Minority Health and Health Disparities, National Institute on Deafness and Other Communication Disorders, National Institute of Dental and Craniofacial Research, National Institute of Diabetes and Digestive and Kidney Diseases, National Institute of Neurological Disorders and Stroke, NIH Institution-Office of Dietary Supplements.

Jackson Heart Study (JHS)

The Jackson Heart Study (JHS) is supported and conducted in collaboration with Jackson State University (HHSN268201300049C and HHSN268201300050C), Tougaloo College (HHSN268201300048C), and the University of Mississippi Medical Center (HHSN268201300046C and HHSN268201300047C) contracts from the National Heart, Lung, and Blood Institute (NHLBI) and the National Institute for Minority Health and Health Disparities (NIMHD). The authors also wish to thank the staff and participants of the JHS.

Multi-Ethnic Study of Atherosclerosis (MESA)

Whole genome sequencing (WGS) for the Trans-Omics in Precision Medicine (TOPMed) program was supported by the National Heart, Lung and Blood Institute (NHLBI). WGS for “NHLBI TOPMed: Multi-Ethnic Study of Atherosclerosis (MESA)” (phs001416.v1.p1) was performed at the Broad Institute of MIT and Harvard (3U54HG003067-13S1). Centralized read mapping and genotype calling, along with variant quality metrics and filtering were provided by the TOPMed Informatics Research Center (3R01HL-117626-02S1, contract HHSN268201800002I). Phenotype harmonization, data management, sample-identity QC, and general study coordination, were provided by the TOPMed Data Coordinating Center (3R01HL-120393; U01HL-120393; contract HHSN268180001I). The MESA project is conducted and supported by the National Heart, Lung, and Blood Institute (NHLBI) in collaboration with MESA investigators. Support for MESA is provided by contracts 75N92020D00001, HHSN268201500003I, N01-HC-95159, 75N92020D00005, N01-HC-95160, 75N92020D00002, N01-HC-95161, 75N92020D00003, N01-HC-95162, 75N92020D00006, N01-HC-95163, 75N92020D00004, N01-HC-95164, 75N92020D00007, N01-HC-95165, N01-HC-95166, N01-HC-95167, N01-HC-95168, N01-HC-95169, UL1-TR-000040, UL1-TR-001079, UL1-TR-001420. Also supported in part by the National Center for Advancing Translational Sciences, CTSI grant UL1TR001881, and the National Institute of Diabetes and Digestive and Kidney Disease Diabetes Research Center (DRC) grant DK063491 to the Southern California Diabetes Endocrinology Research Center. Infrastructure for the CHARGE Consortium is supported in part by the National Heart, Lung, and Blood Institute (NHLBI) grant R01HL105756.

Women’s Health Initiative (WHI)

The WHI program is funded by the National Heart, Lung, and Blood Institute, National Institutes of Health, U.S. Department of Health and Human Services through contracts 75N92021D00001, 75N92021D00002, 75N92021D00003, 75N92021D00004, 75N92021D00005.

M.M.W. was supported by a fellowship from the NHLBI BioData Catalyst program (award 1OT3HL142479-01, 1OT3HL142478-01, 1OT3HL142481-01, 1OT3HL142480-01, 1OT3HL147154). D.E.B. was supported by NHLBI DP2HL137300, R01HL130733, R01HL150553. Y.L. and J.W. were partially supported by U01HG011720 and R01HL129132. N.P. and J.L. were partially supported by R01HL154385. A.P.R was partially supported by R01 HL146500 and R01 HL130733. We thank S.A. Wolfe for sharing 3xNLS-SpCas9 protein; R. Mathieu and the HSCI-BCH Flow Cytometry Facility, supported by the Harvard Stem Cell Institute and the NIH (U54DK110805) for assistance with flow cytometry; Fred Hutchinson Cancer Research Center, Seattle, Washington for CD34+ HSPCs (supported by Cooperative Centers of Excellence in Hematology NIDDK Grant U54DK106829).

The views expressed in this manuscript are those of the authors and do not necessarily represent the views of the National Heart, Lung, and Blood Institute; the National Institutes of Health; or the U.S. Department of Health and Human Services.

