## Supplementary_materials for "Whole genome sequencing identifies common and rare structural variants contributing to hematologic traits in the NHLBI TOPMed program": Supplemental_figures_S1-S6.pdf

### FIGURE LEGENDS

**Figure S1. QQ plots of the marginal structural variant analyses in TOPMed.** (A) HCT; (B) HGB; (C) MCH; (D) MCHC; (E) MCV; (F) RBC; (G) RDW; (H) BASO_binary; (I) EOSIN; (J) LYMPHS; (K) MONOS; (L) NEUTRO; (M) WBC; (N) EOSIN%; (O) LYMPHS%; (P) MONOS%; (Q) NEUTRO%; (R) MPV; (S) PLT; (T) FERRITIN; (U) IRON; (V) SAT; (W) UIBC; (X) TIBC.

**Figure S2. Manhattan plots of the marginal structural variant analyses in TOPMed.** (A) HCT; (B) HGB; (C) MCH; (D) MCHC; (E) MCV; (F) RBC; (G) RDW; (H) BASO_binary; (I) EOSIN; (J) LYMPHS; (K) MONOS; (L) NEUTRO; (M) WBC; (N) EOSIN%; (O) LYMPHS%; (P) MONOS%; (Q) NEUTRO%; (R) MPV; (S) PLT; (T) FERRITIN; (U) IRON; (V) SAT; (W) UIBC; (X) TIBC.

**Figure S3. Locus zoom plots for structural variants significantly associated with hematological and hemostasis traits in TOPMed.** A) EOSIN, B) LYMPH, C) LYMPH PROP, D) MONOS, E) MONOS PROP, F) NEUTRO PROP, G) WBC, H) HCT, I) HGB, J) MCH, K) MCHC, L) MCV, M) RBC, N) RDW, O), MPV, P) PLT, Q) TIBC, R) UIBC

**Figure S4. Visualization of WGS reads for conditionally-independant trait associatiaed SVs.** WGS reads were visualized in TOPMed samples predicted by Parliament2 to exhibit the SV event. WGS reads were visualized using SAMPLOT, which displays coverage and paired-end read evidence supporting SV events.

**Figure S5. Comparison of mean read-depth for an alternative HLA locus (chr6_GL000256v2_alt) and chr2:178436244 (rs62176107) genotypes.** Genotypes for chr2:178436244 (rs62176107) are missing (./.), homozygous reference (0/0), heterozygous (0/1) and homozygous alternate (1/1) and are plotted against the mean read-depth estimates for chr6_GL000256v2_alt.

**Figure S6. Comparison of trait-association p-values for previously-reported SNVs from a European ancestry sample set[10] and the TOPMed SV tagging each SNV for pairs with r^2 > 0.8**. Points with p < 1e-4 in either analysis are labeled with the trait.

**Figure S7. Chromatin status and chromatin conformation across the 9q22.1 locus (Related to Figure 1).**

(A) Distribution of accessible chromatin (by DNase I sequencing) and histone modifications (H3K27ac, H3K4me1 and H3K4me3) in CD34+ common myeloid progenitors (CMP) and human mesenchymal stem cells (MSCs) across indicated genomic region. Datasets are obtained from the ENCODE project[27].

(B) Virtual 4C plot of long-range chromatin interactions anchored at the deletion (chr9:88923551-88924152, upper panel) and the *S1PR3* promoter (chr9:91605763-91606263, lower panel), shown as a grey bar, in monocytes[28]. Yellow line highlights the *S1PR3* promoter (chr9:91605763-91606263, upper panel) and the deletion (chr9:88923551-88924152, lower panel). The observed and expected chromatin contact frequencies (or counts) are represented by the black and red lines, respectively. The left Y axis displays the range of chromatin contact frequency. The statistical significance (–log10(P-value)) of each long-range chromatin interaction is represented by the blue line, with its range listed in the right Y axis. The cell line or tissue specific FDR threshold (5%) is shown as a cyan horizontal dashed line, and the more stringent Bonferroni threshold (P = 0.05) is shown as a blue horizontal dashed line.

**Figure S8. Epigenome editing implicates *S1PR3* in the 9q22.1 monocyte count association (Related to Figure 2).**

(A-B) eQTL analysis between the 9q22.1 SV and genes within 2 Mb window in peripheral blood mononuclear cells (PBMCs; A) and T-cell (B) using data from the ancestry-stratified datasets from the Multi-Ethnic Study of Atherosclerosis (MESA, including n = 229 African American and n = 381 Hispanic/Latino participants). AFHI: African American and Hispanic/Latino.

(C) Schematic of sgRNA locations targeting the structural variant (chr9:88923551-88924152, upper panel). PAM sequences highlighted in green.

(D) Expression of genes within a 2 Mb window in THP-1 cells after CRISPRi targeting the structural variant sequences. Relative mRNA level of each gene was represented by mean ± standard deviation (SD). N = 3 technical replicates.

(E) Expression of each gene within a 2 Mb window around the 9q22.1 locus in THP-1 cells relative to *GAPDH*. Genes of undetectable expression level (*CDK20*, *SPATA31C2*, *SPIN1*, *SHC3*, *GADD45G* and *UNQ6494*) are not shown. N = 3 technical replicates.

**Figure S9. Human hematopoietic reconstitution of immunodeficient mice after *S1PR3* editing (Related to Figure 2).**

(A-D) Healthy donor CD34+ HSPCs were edited by indicated RNP and infused to NBSGW mice. After 12 weeks, bone marrow cells were analyzed by flow cytometry. Each symbol represents one mouse. HSPCs, hCD45+CD19-CD33-CD34+; B cells, hCD45+CD19+; Erythroid, hCD45-CD45-CD235a+; Myeloid, hCD45+CD33+.

**Figure S1**

(A)

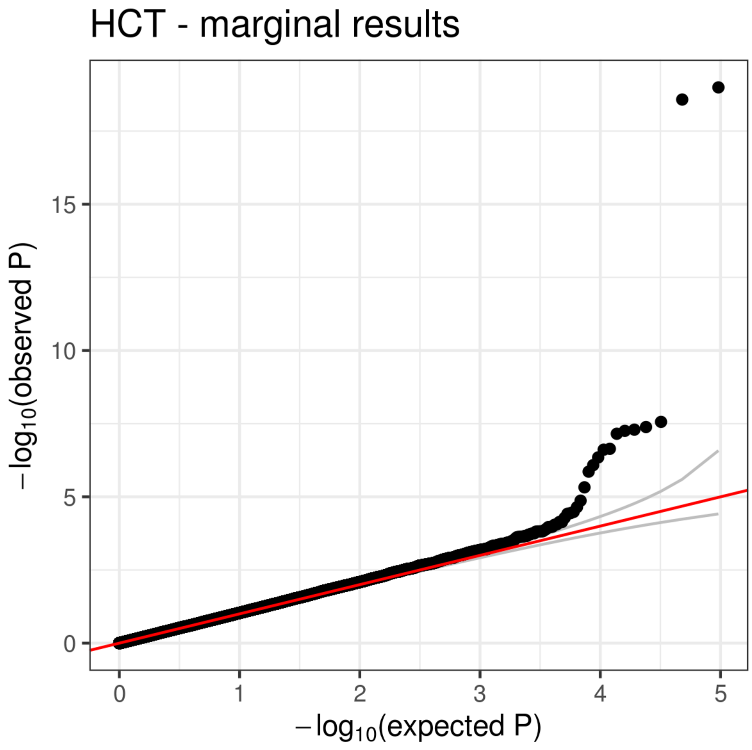
HCT - marginal results
15
10
5
0
$-\log_{10}$(observed P)
0
1
2
3
4
5
$-\log_{10}$(expected P)

(B)

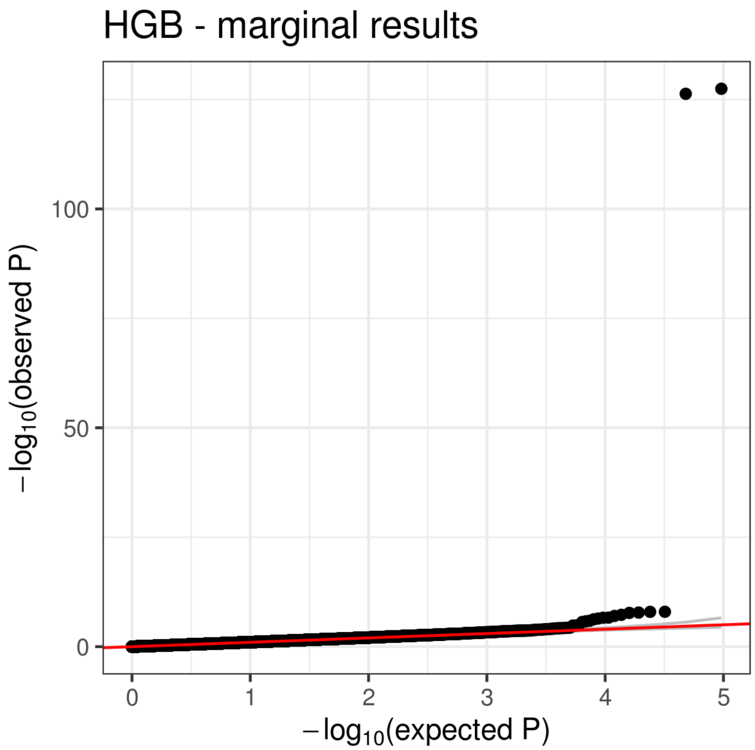
HGB - marginal results
100
50
0
−log10(observed P)
0
1
2
3
4
5
−log10(expected P)

(C)

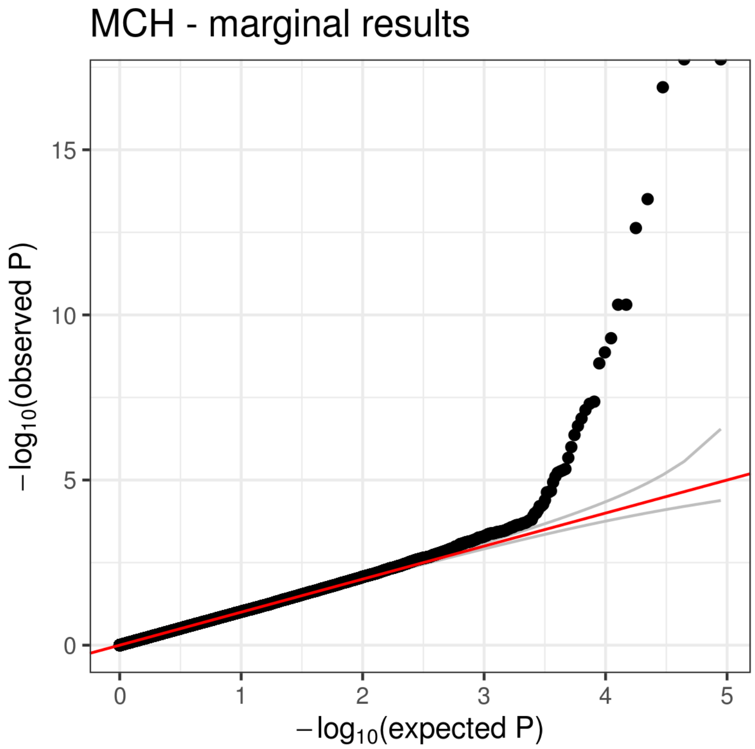
MCH - marginal results
15
10
5
0
−log10(observed P)
0
1
2
3
4
5
−log10(expected P)

(D)

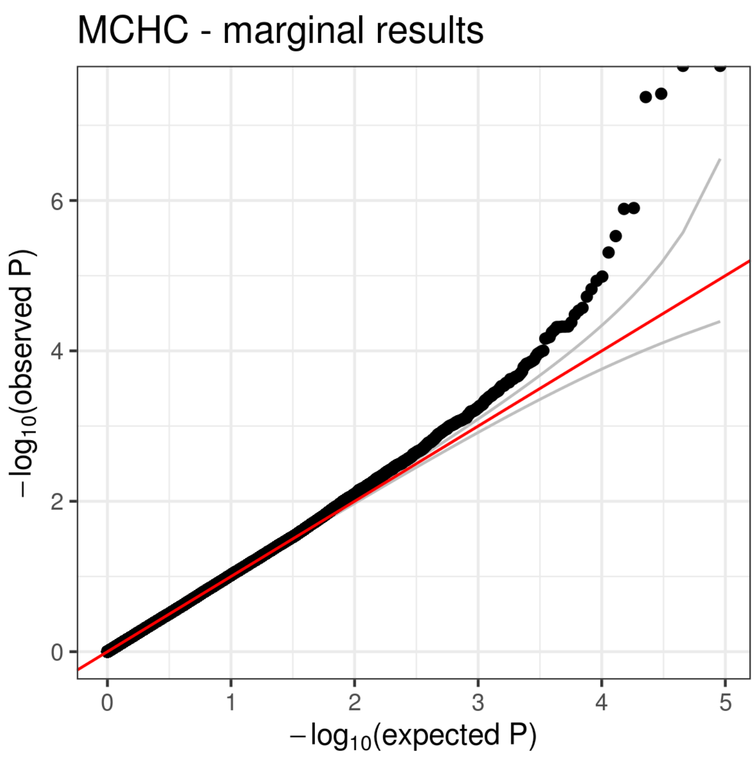

MCHC - marginal results
6
4
2
0
0
1
2
3
4
5
−log10(observed P)
−log10(expected P)

(E)

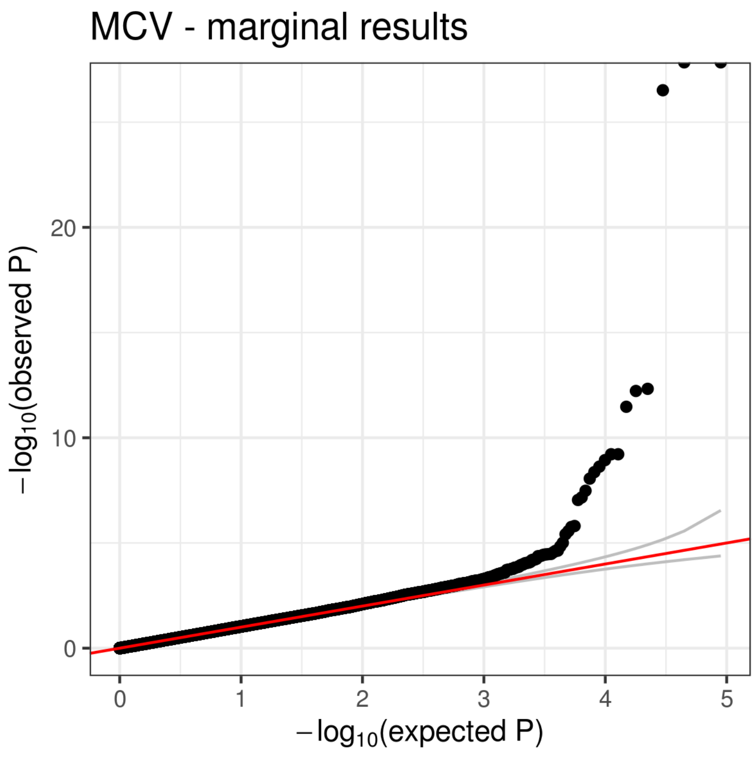

MCV - marginal results
20
10
0
0
1
2
3
4
5
−log10(observed P)
−log10(expected P)

(F)

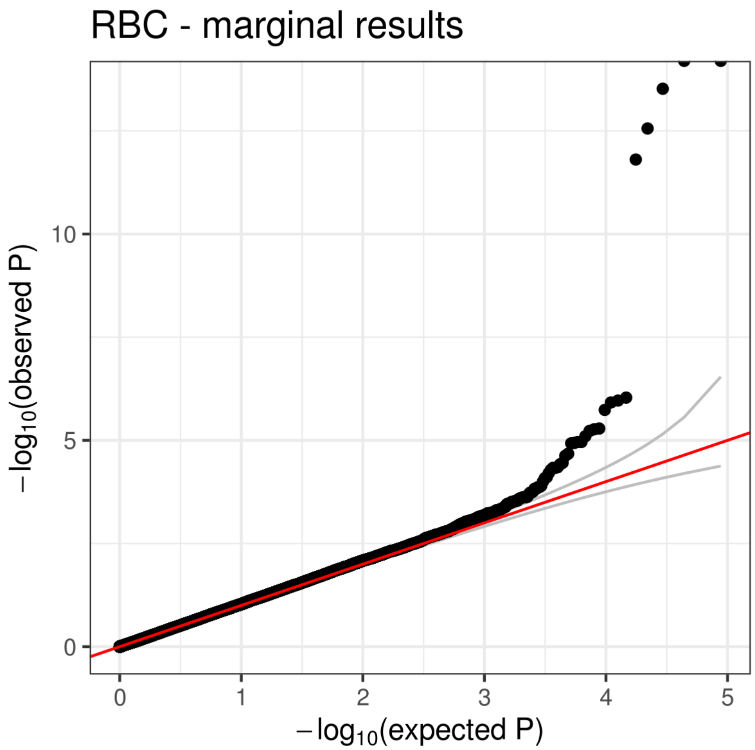
RBC - marginal results
$-\log_{10}$(observed P)
$-\log_{10}$(expected P)
0
5
10
0
1
2
3
4
5

(G)

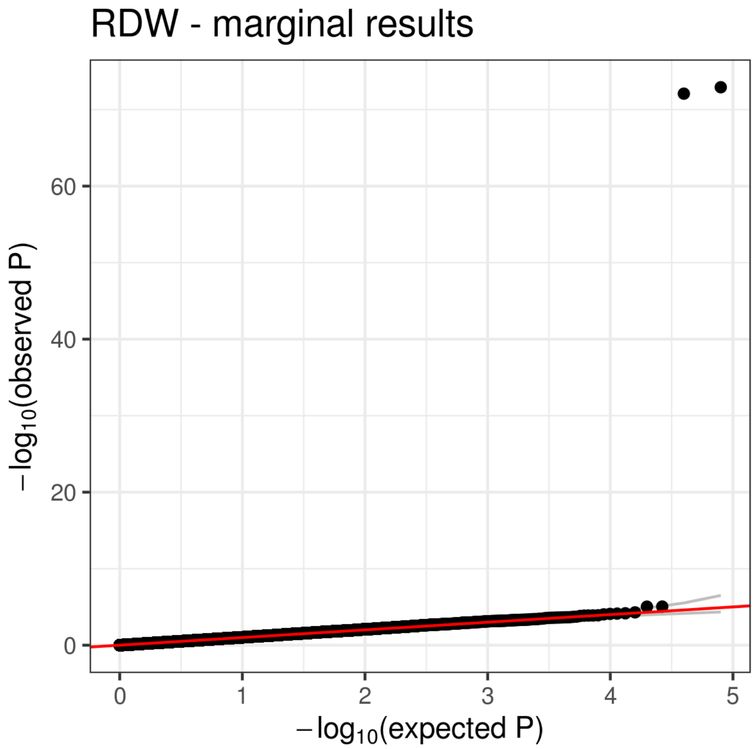
RDW - marginal results
$-\log_{10}$(observed P)
$-\log_{10}$(expected P)
0
20
40
60
0
1
2
3
4
5

(H)

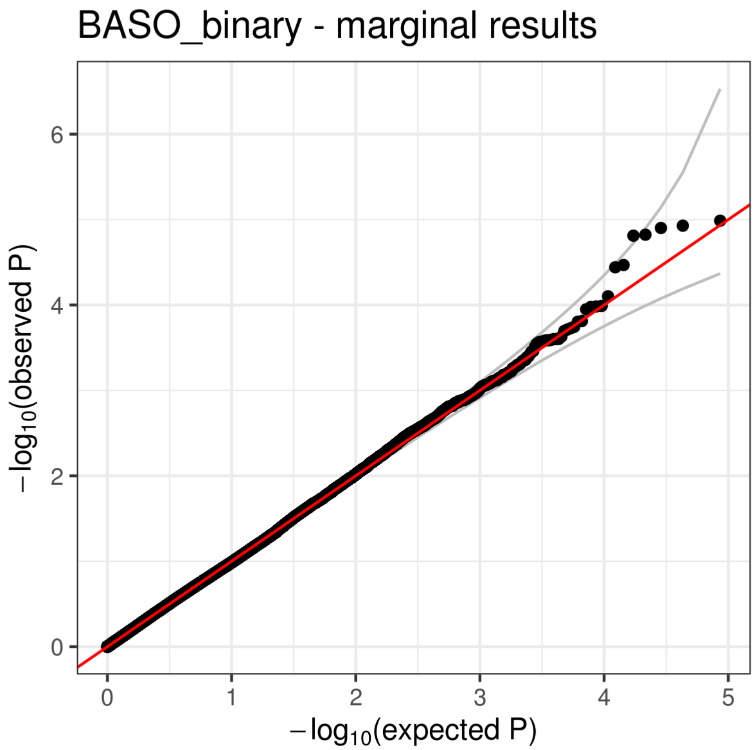
BASO_binary - marginal results
−log10(observed P)
−log10(expected P)
0
2
4
6
0
1
2
3
4
5

(I)

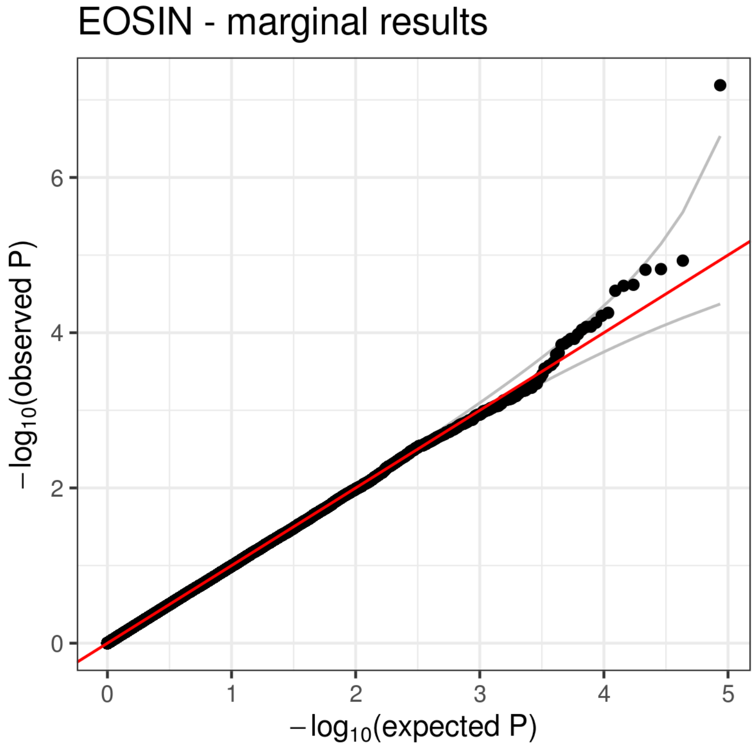
EOSIN - marginal results
−log10(observed P)
−log10(expected P)
0
2
4
6
0
1
2
3
4
5

(J)

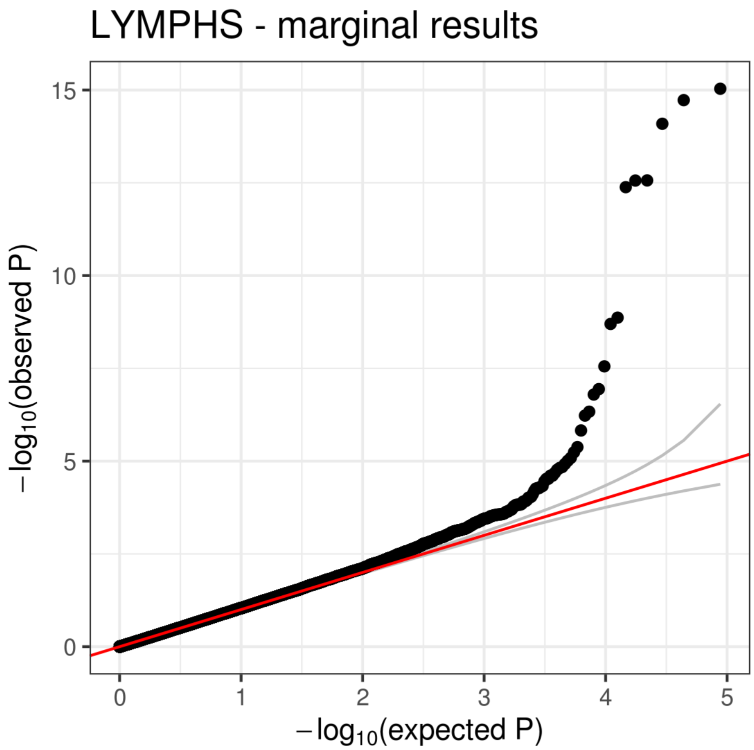

LYMPHS - marginal results
15
10
5
0
0
1
2
3
4
5
$-\log_{10}$(observed P)
$-\log_{10}$(expected P)

(K)

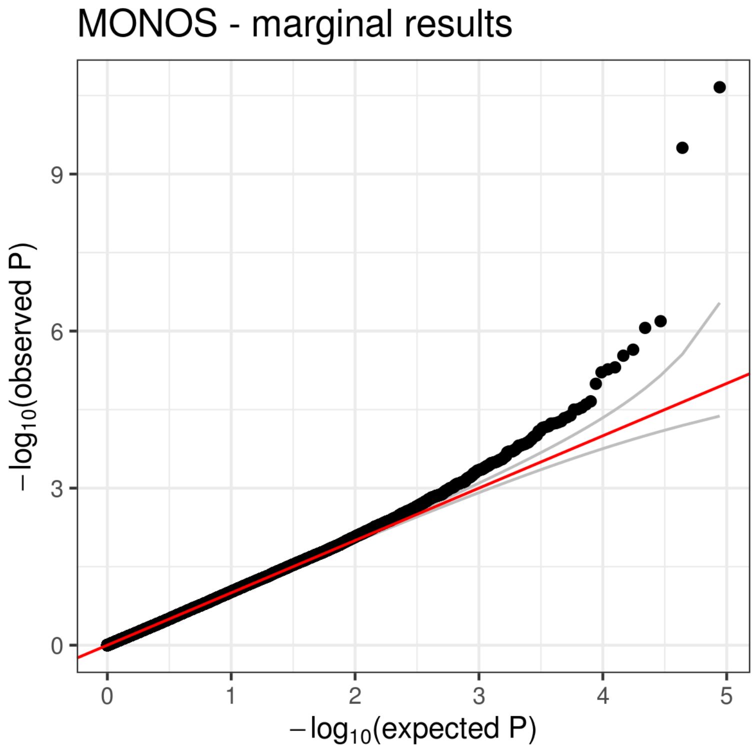

MONOS - marginal results
9
6
3
0
0
1
2
3
4
5
$-\log_{10}$(observed P)
$-\log_{10}$(expected P)

(L)

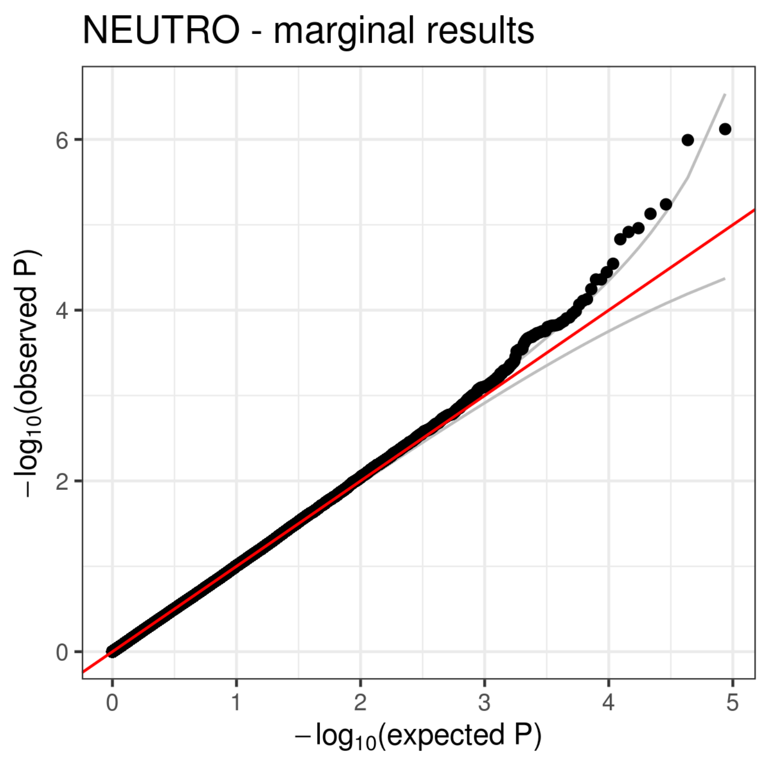
NEUTRO - marginal results
−log10(observed P)
−log10(expected P)
0
2
4
6
0
1
2
3
4
5

(M)

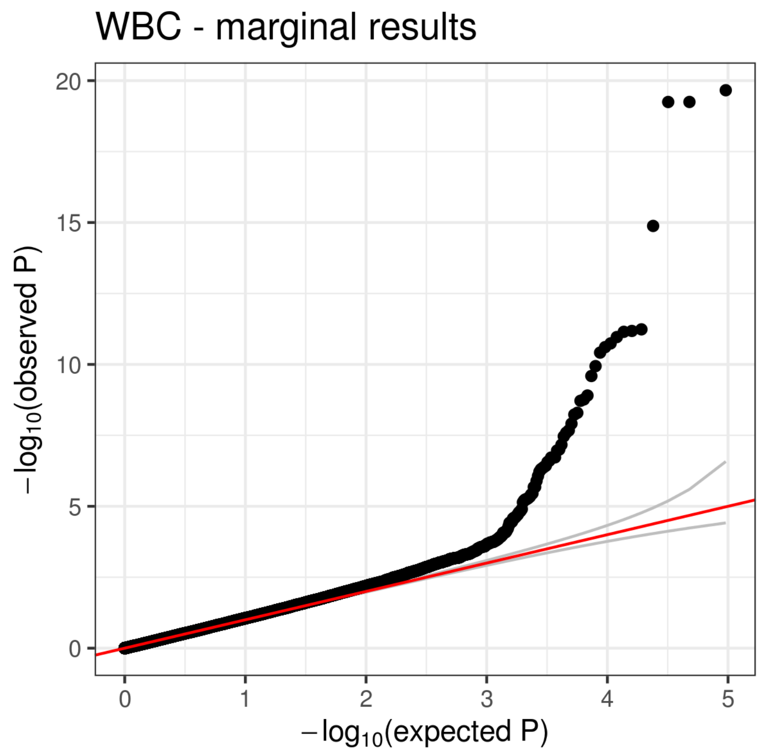
WBC - marginal results
−log10(observed P)
−log10(expected P)
0
5
10
15
20
0
1
2
3
4
5

(N)

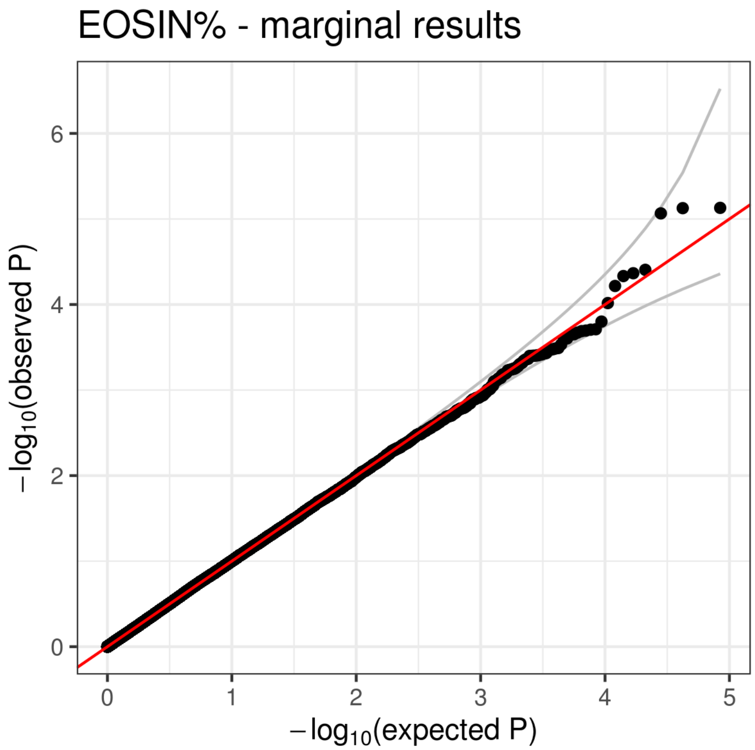
EOSIN% - marginal results
−log10(observed P)
−log10(expected P)
0
2
4
6
0
1
2
3
4
5

(O)

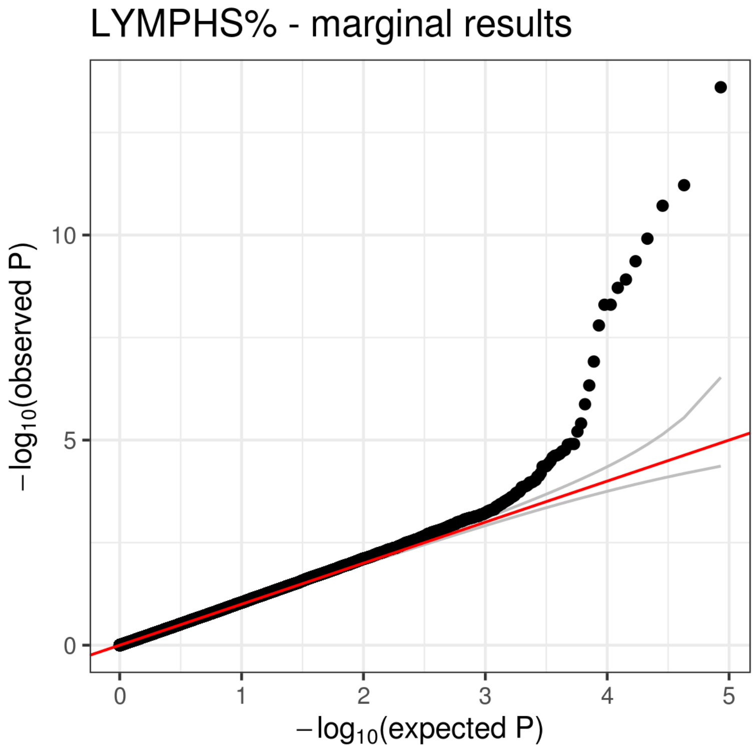
LYMPHS% - marginal results
−log10(observed P)
−log10(expected P)
0
5
10
0
1
2
3
4
5

(P)

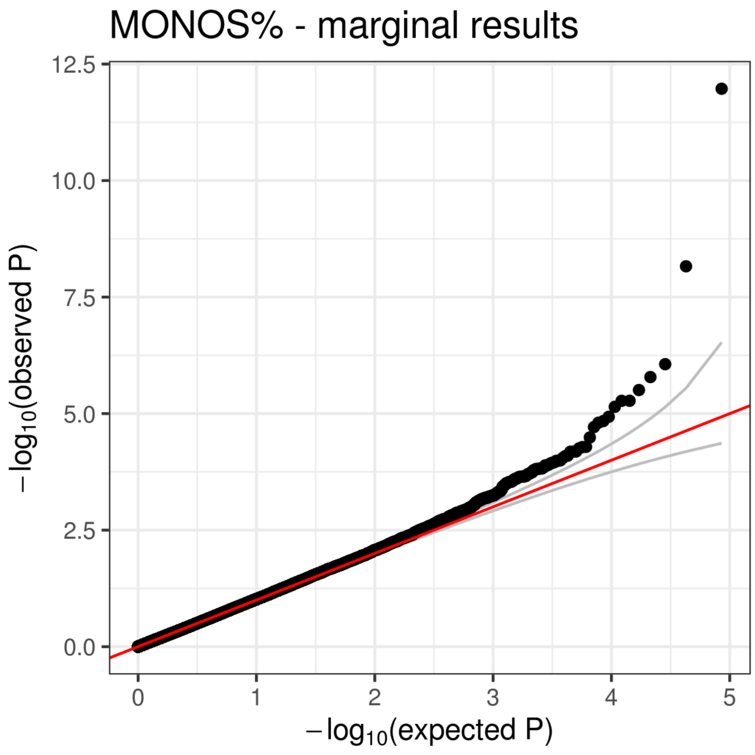
MONOS% - marginal results
12.5
10.0
7.5
5.0
2.5
0.0
0
1
2
3
4
5
$-\log_{10}$(observed P)
$-\log_{10}$(expected P)

(Q)

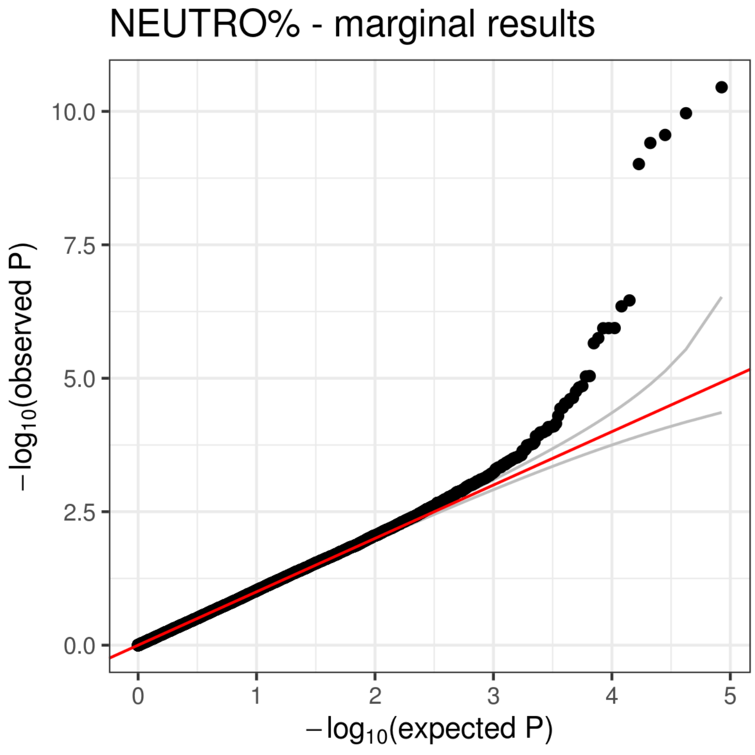
NEUTRO% - marginal results
10.0
7.5
5.0
2.5
0.0
0
1
2
3
4
5
$-\log_{10}$(observed P)
$-\log_{10}$(expected P)

(R)

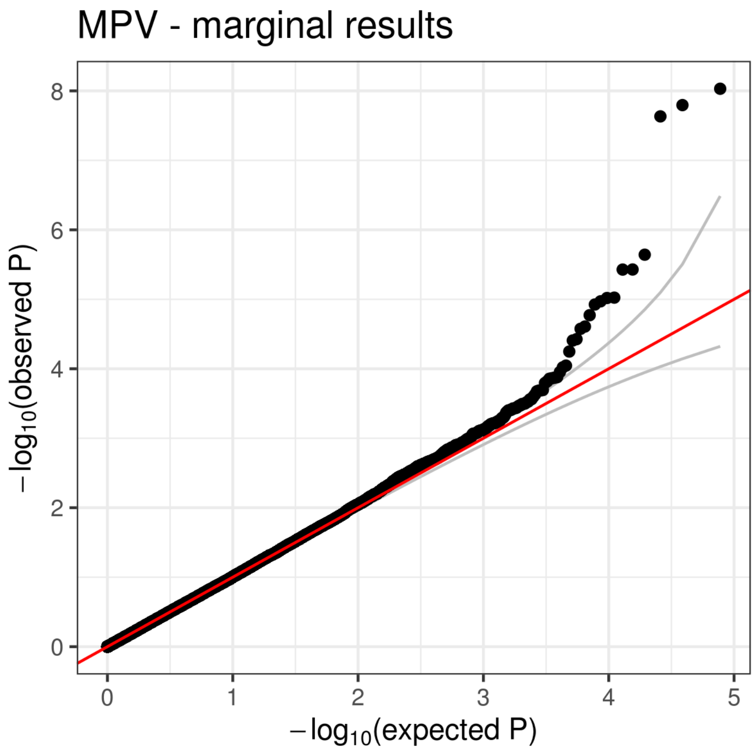
MPV - marginal results
8
6
4
2
0
0
1
2
3
4
5
$-\log_{10}$(observed P)
$-\log_{10}$(expected P)

(S)

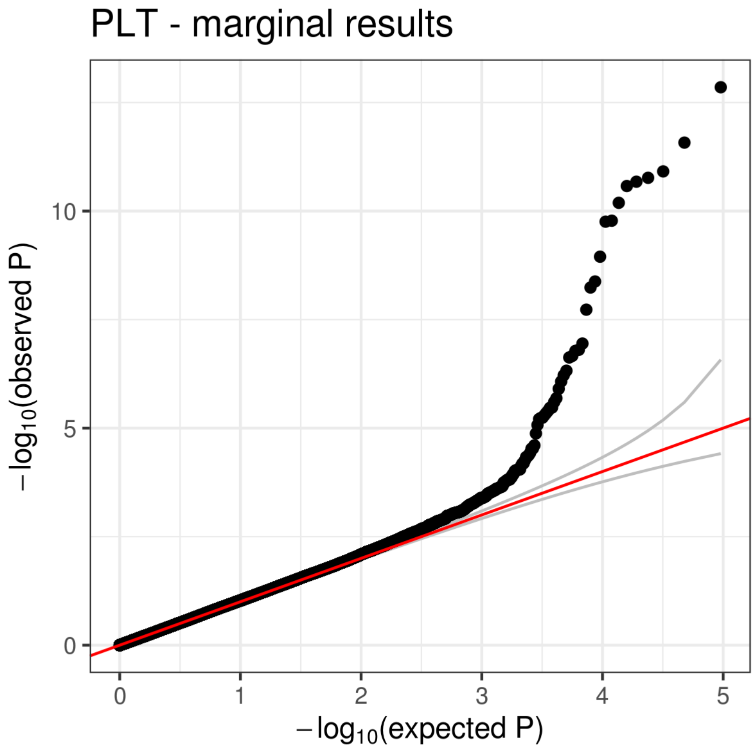
PLT - marginal results
10
5
0
0
1
2
3
4
5
$-\log_{10}$(observed P)
$-\log_{10}$(expected P)

(T)

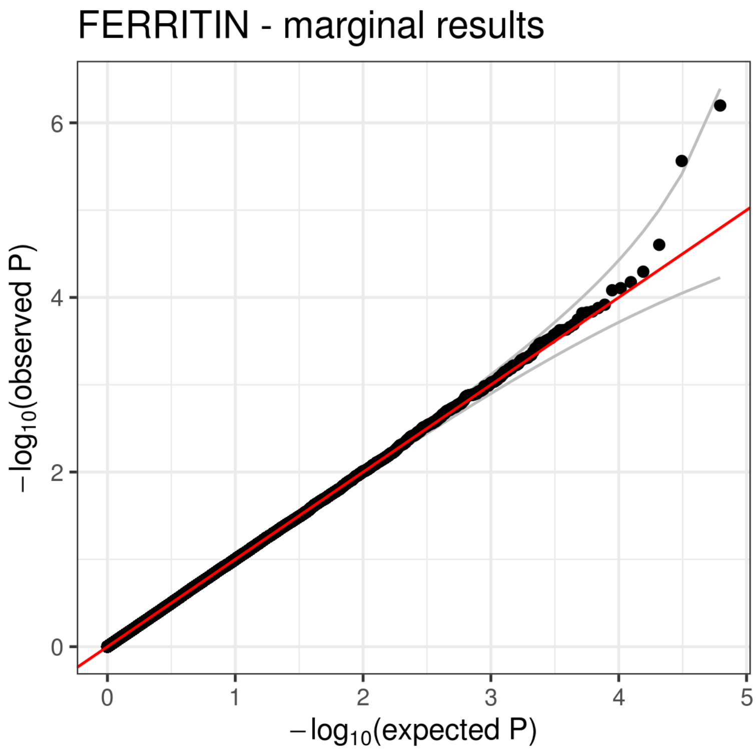
FERRITIN - marginal results
−log10(observed P)
0
2
4
6
0
1
2
3
4
5
−log10(expected P)

(U)

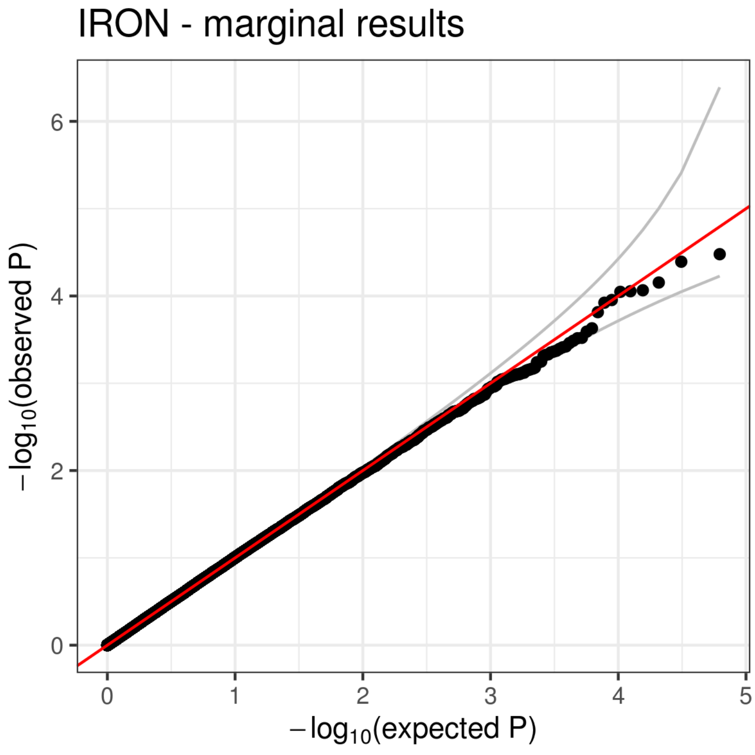
IRON - marginal results
−log10(observed P)
0
2
4
6
0
1
2
3
4
5
−log10(expected P)

(V)

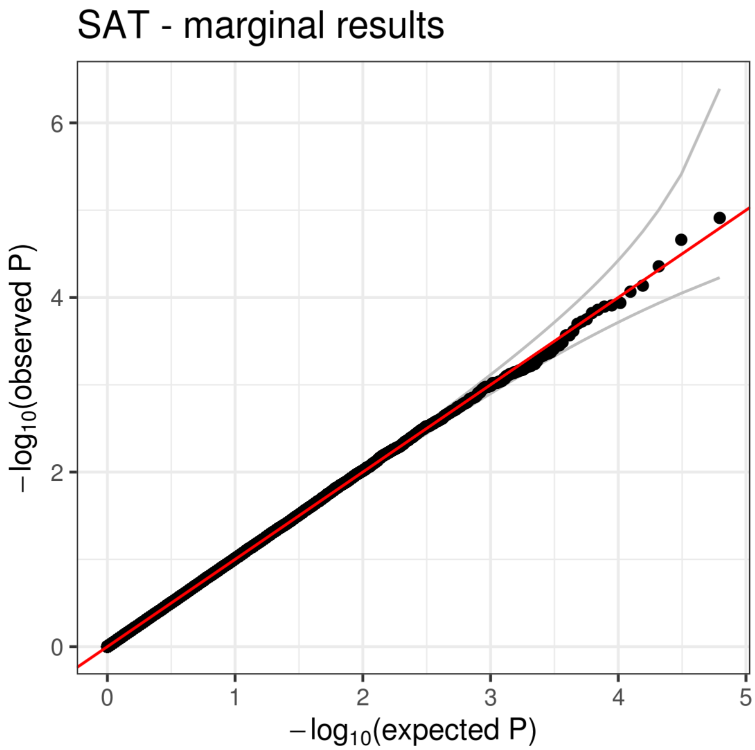
SAT - marginal results
$-\log_{10}$(observed P)
$-\log_{10}$(expected P)
0
2
4
6
0
1
2
3
4
5

(W)

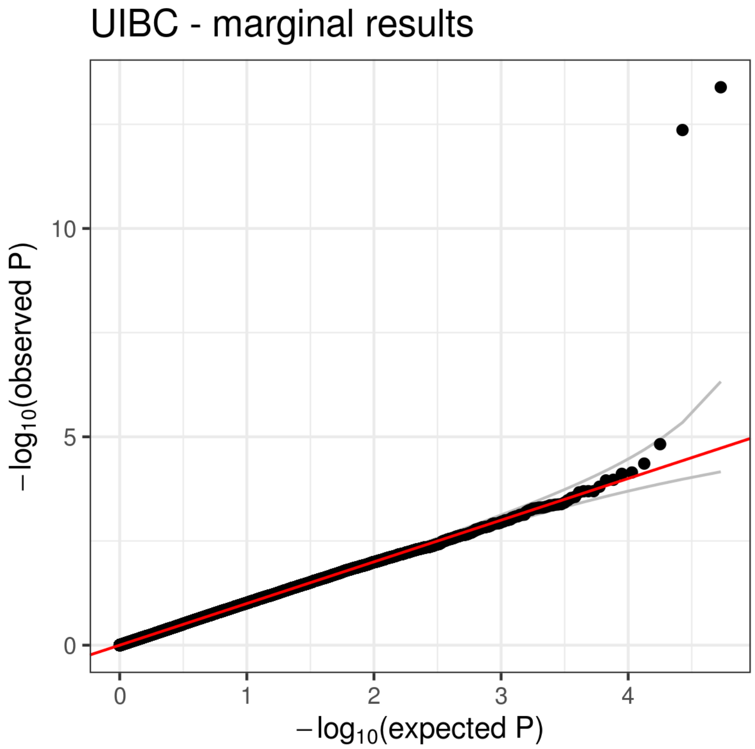
UIBC - marginal results
$-\log_{10}$(observed P)
$-\log_{10}$(expected P)
0
5
10
0
1
2
3
4

(X)

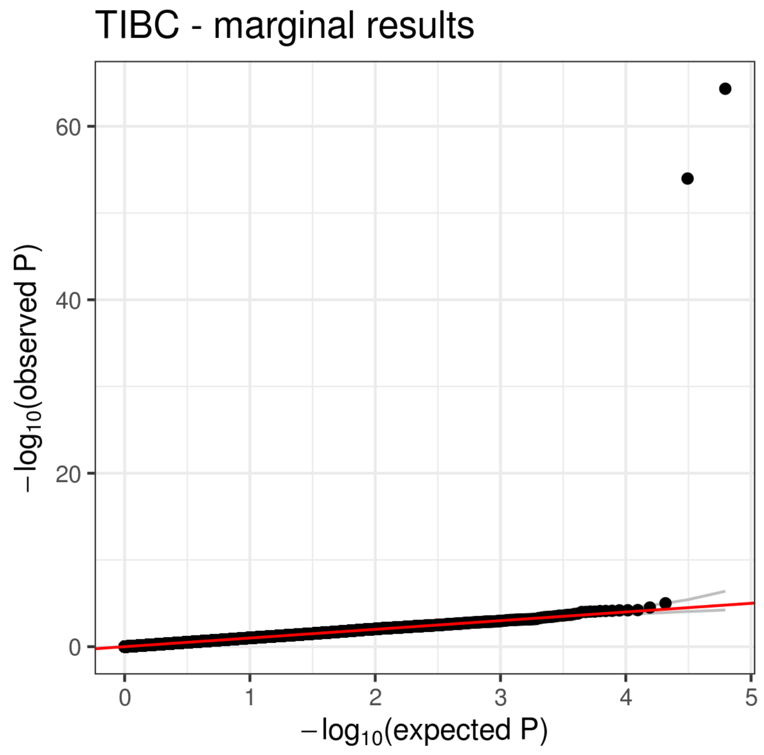
TIBC - marginal results
60
40
20
0
0
1
2
3
4
5
$-\log_{10}$(observed P)
$-\log_{10}$(expected P)

**Figure S2**

(A)

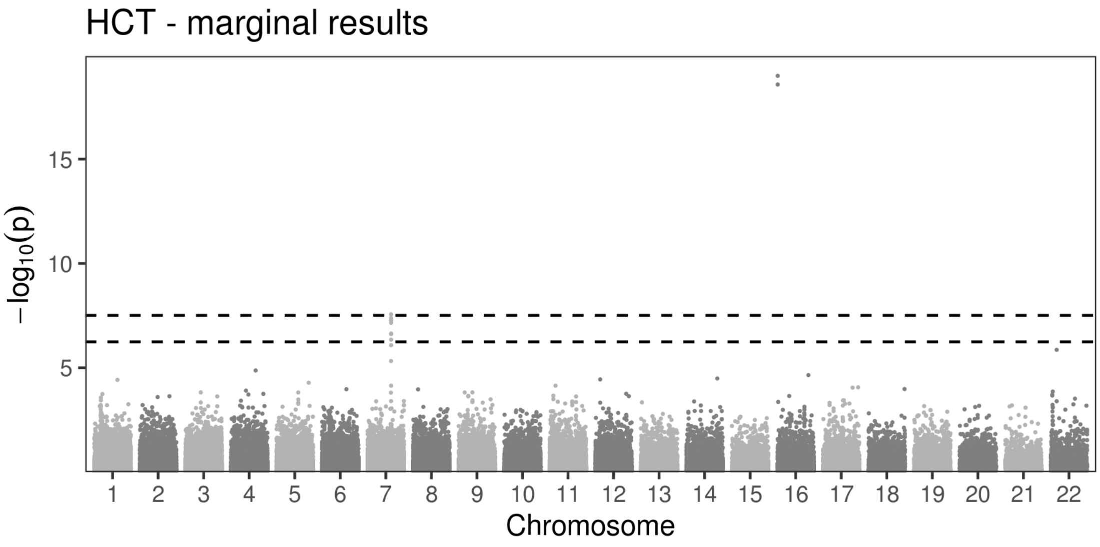

(B)

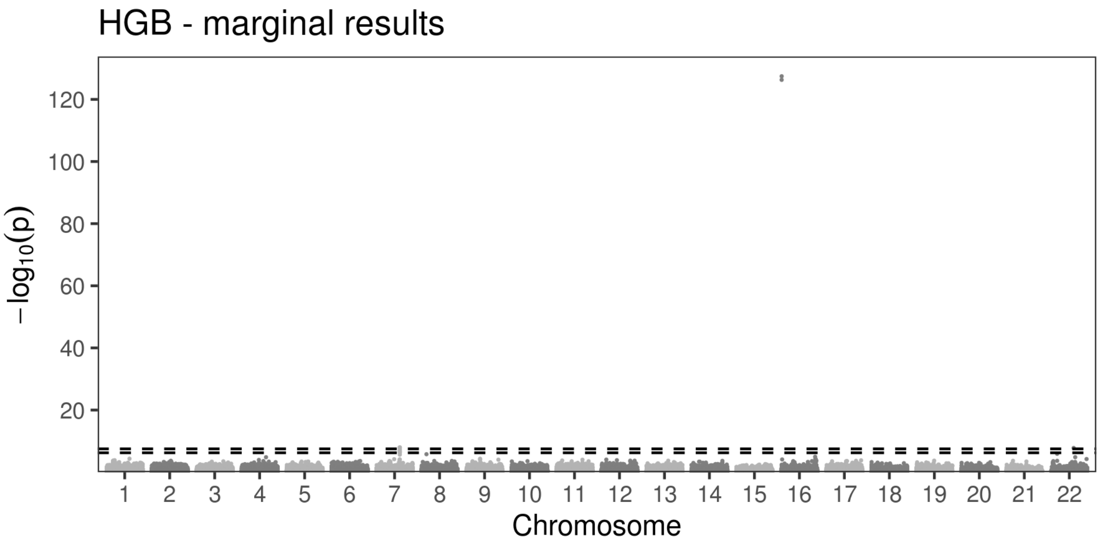

(C)

MCH - marginal results

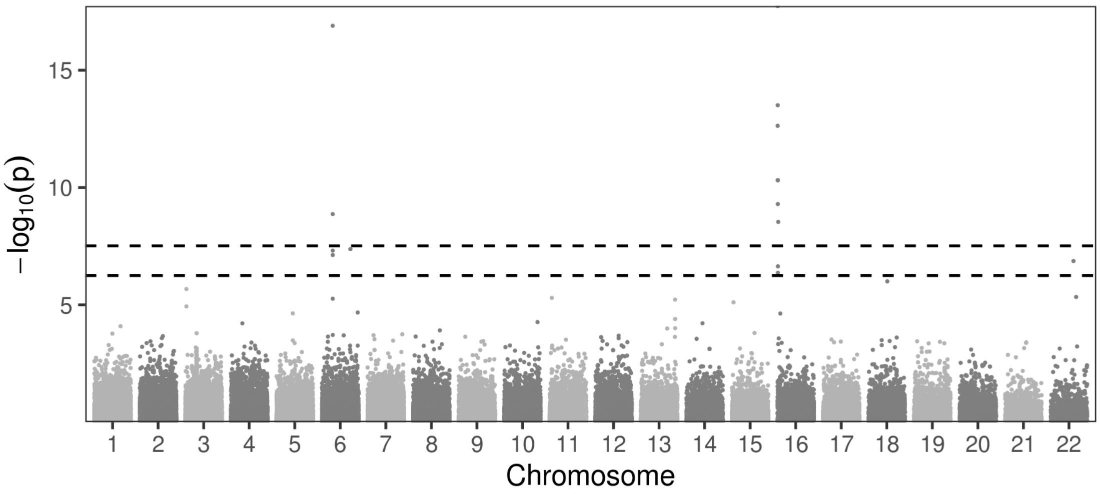

(D)

MCHC - marginal results

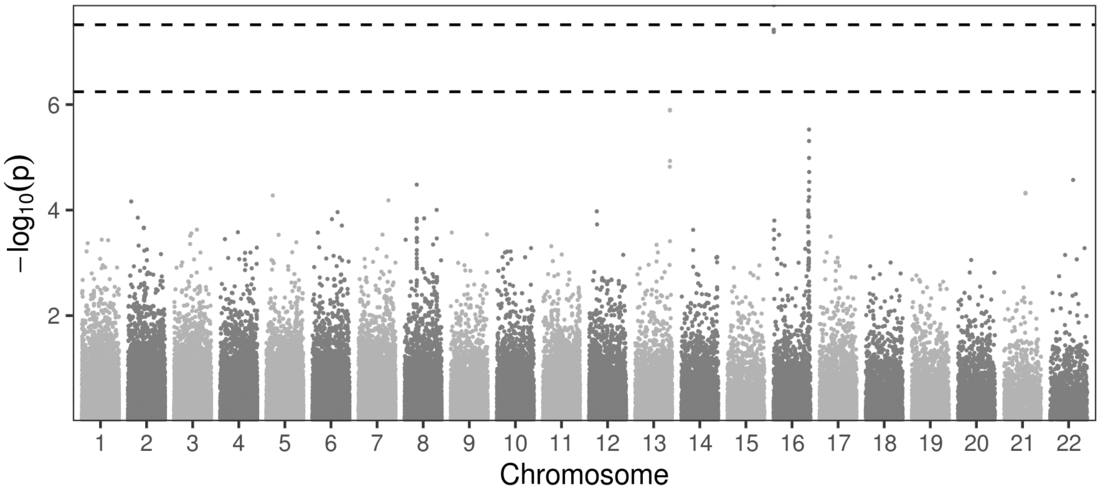

(E)

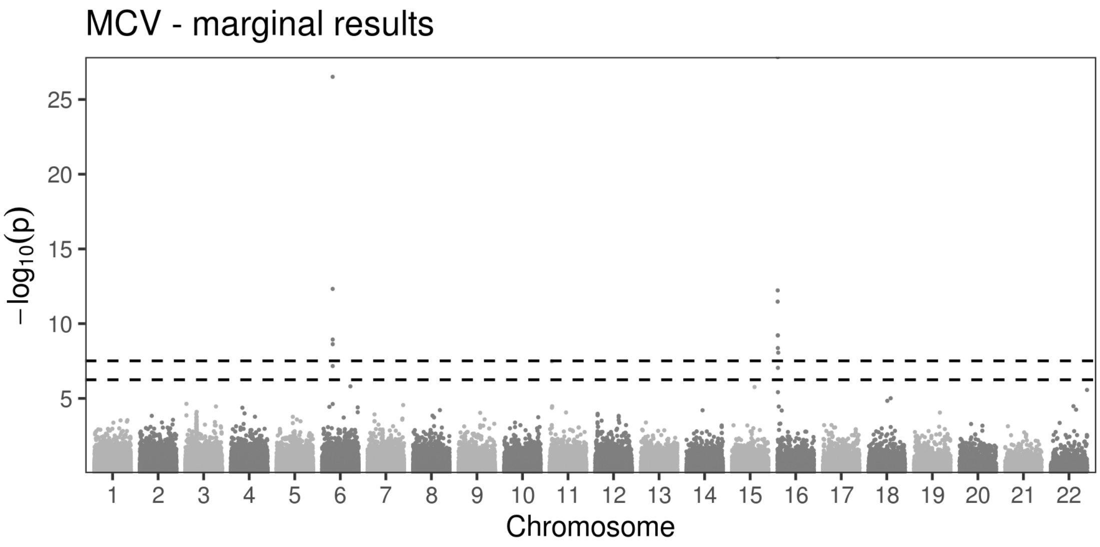
MCV - marginal results
25
20
15
10
5
−log10(p)
Chromosome

(F)

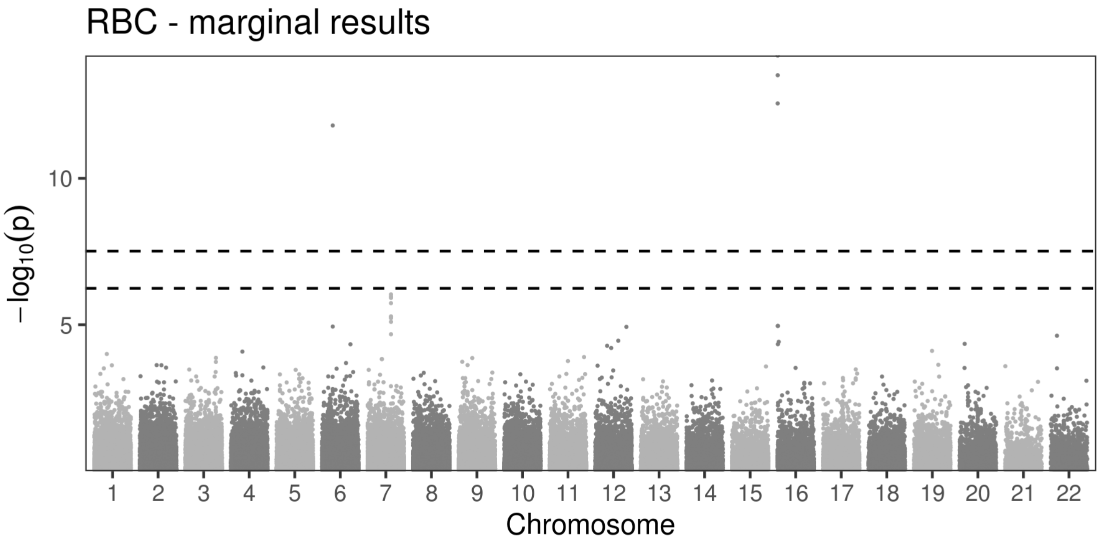
RBC - marginal results
10
5
−log10(p)
Chromosome

(G)

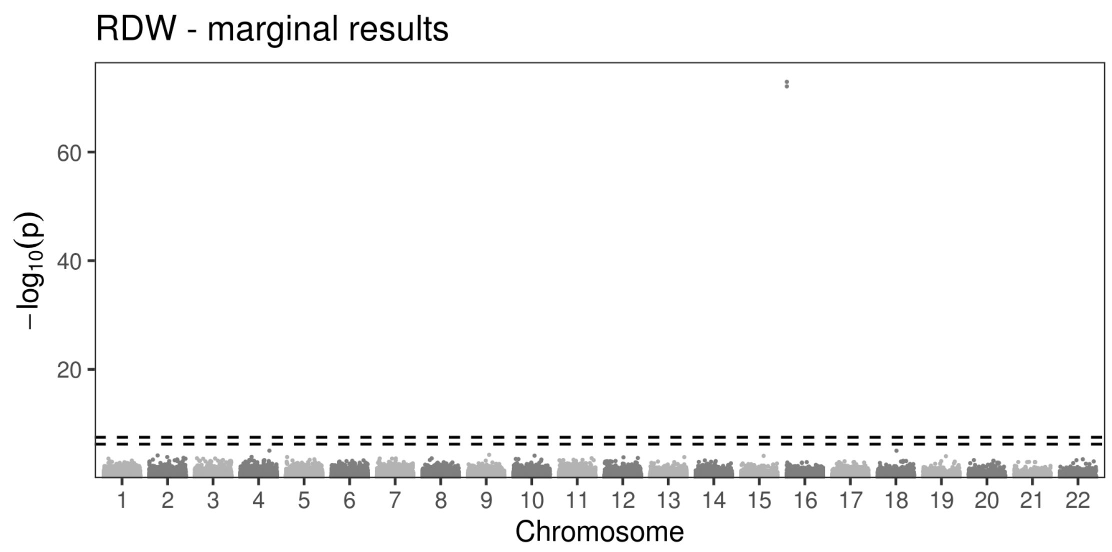
RDW - marginal results
60
40
20
−log10(p)
Chromosome

(H)

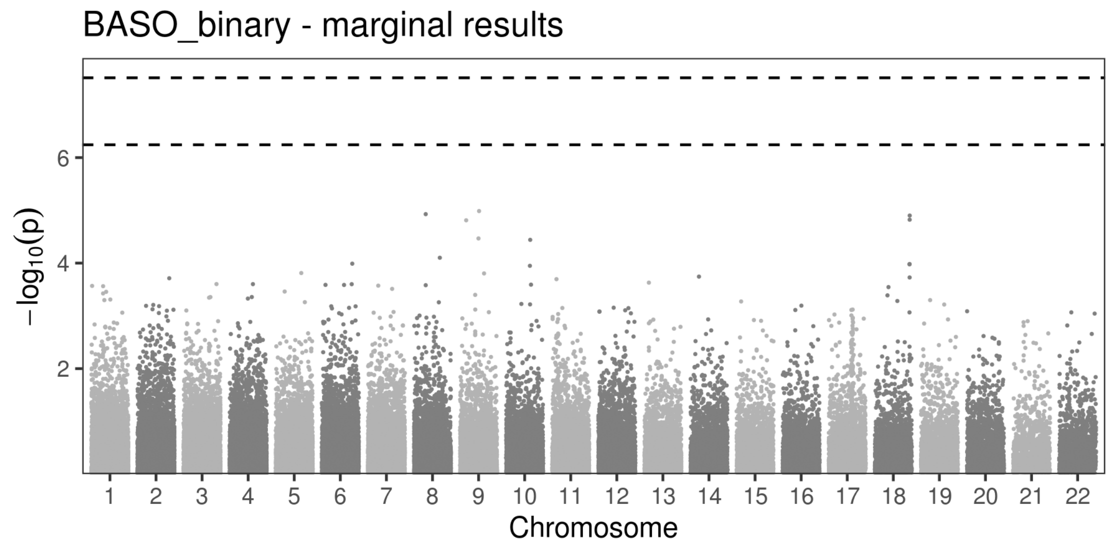
BASO_binary - marginal results
6
4
2
−log10(p)
Chromosome

(I)

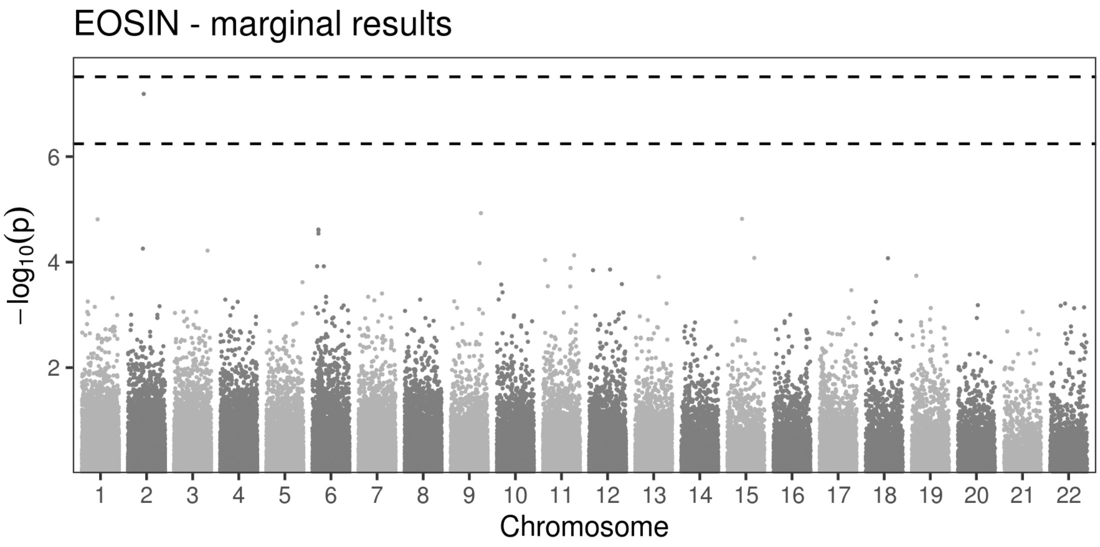
EOSIN - marginal results
−log10(p)
6
4
2
Chromosome

(J)

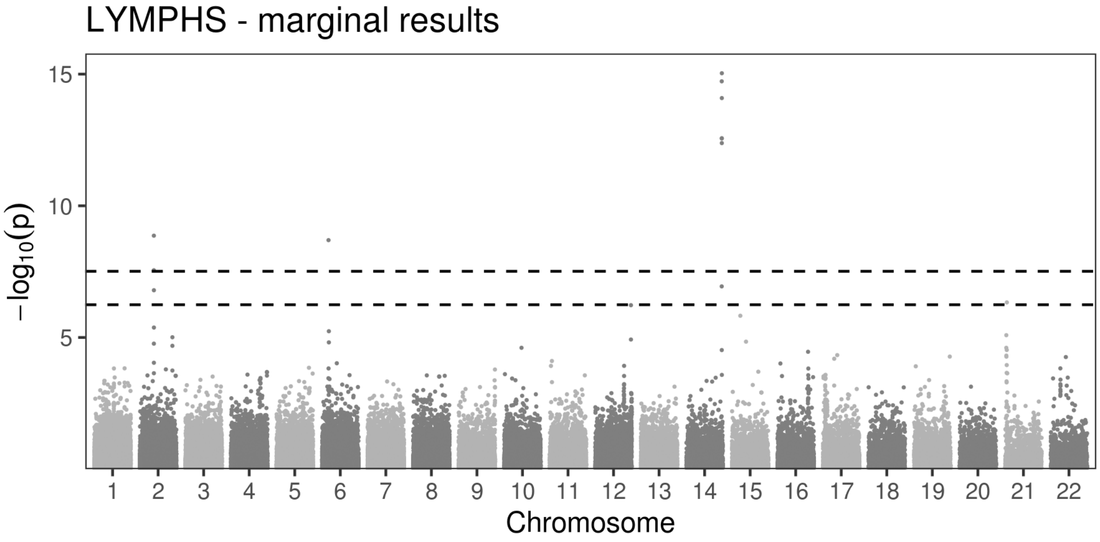
LYMPHS - marginal results
−log10(p)
15
10
5
Chromosome

(K)

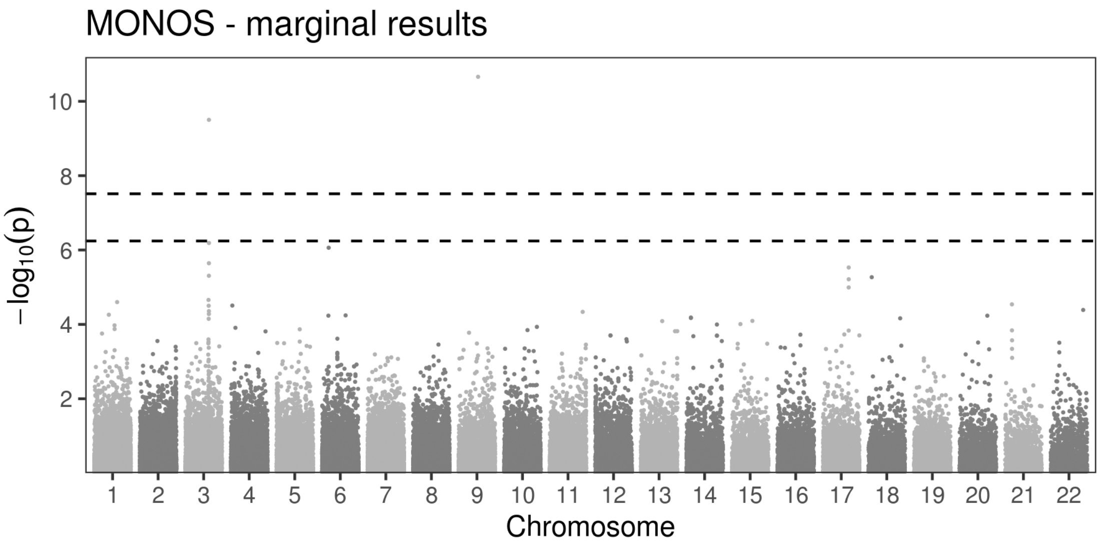
MONOS - marginal results
−log10(p)
10
8
6
4
2
1 2 3 4 5 6 7 8 9 10 11 12 13 14 15 16 17 18 19 20 21 22
Chromosome

(L)

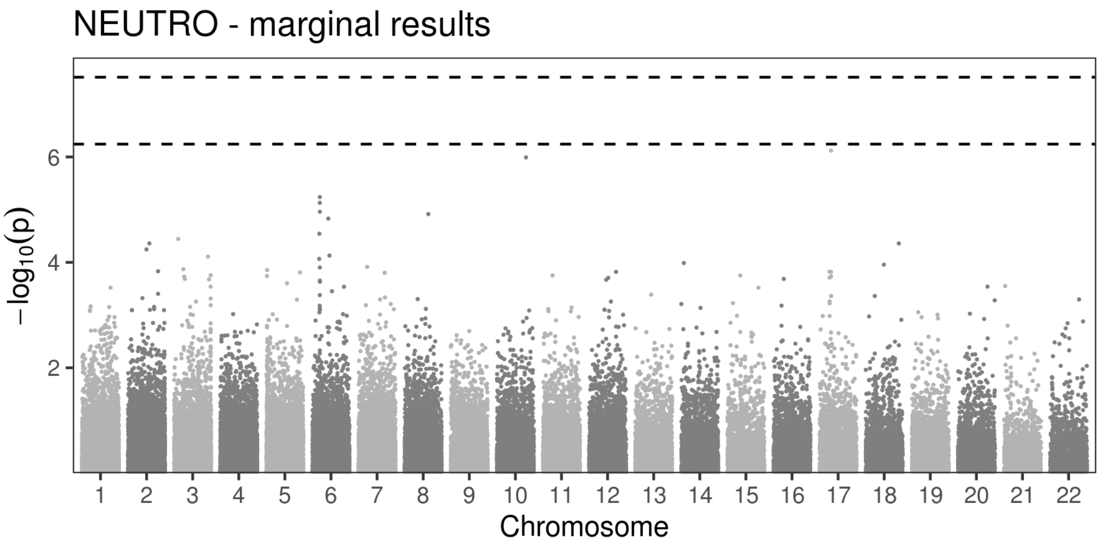
NEUTRO - marginal results
−log10(p)
6
4
2
1 2 3 4 5 6 7 8 9 10 11 12 13 14 15 16 17 18 19 20 21 22
Chromosome

(M)

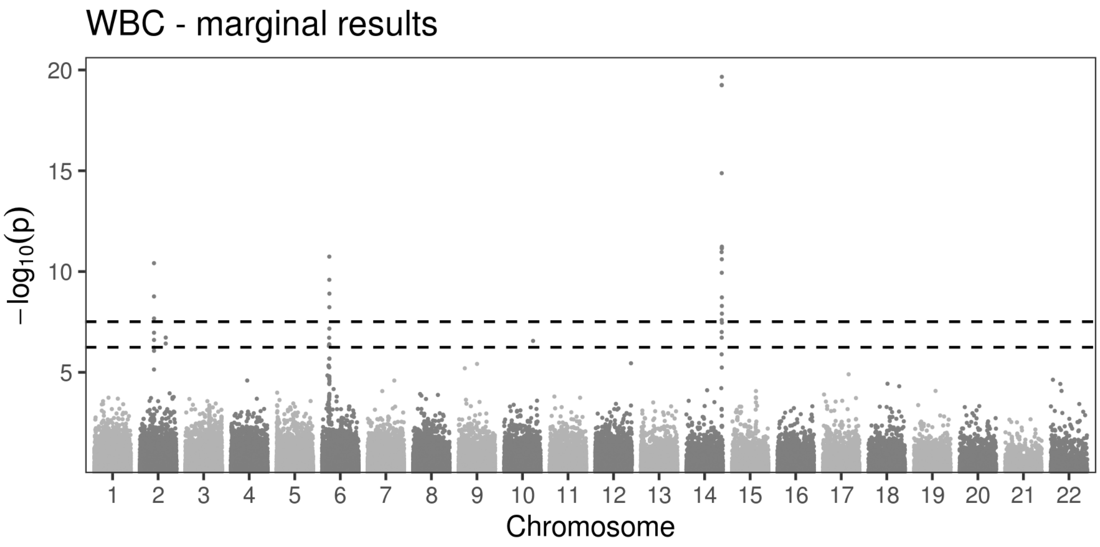
WBC - marginal results
−log10(p)
20
15
10
5
1 2 3 4 5 6 7 8 9 10 11 12 13 14 15 16 17 18 19 20 21 22
Chromosome

(N)

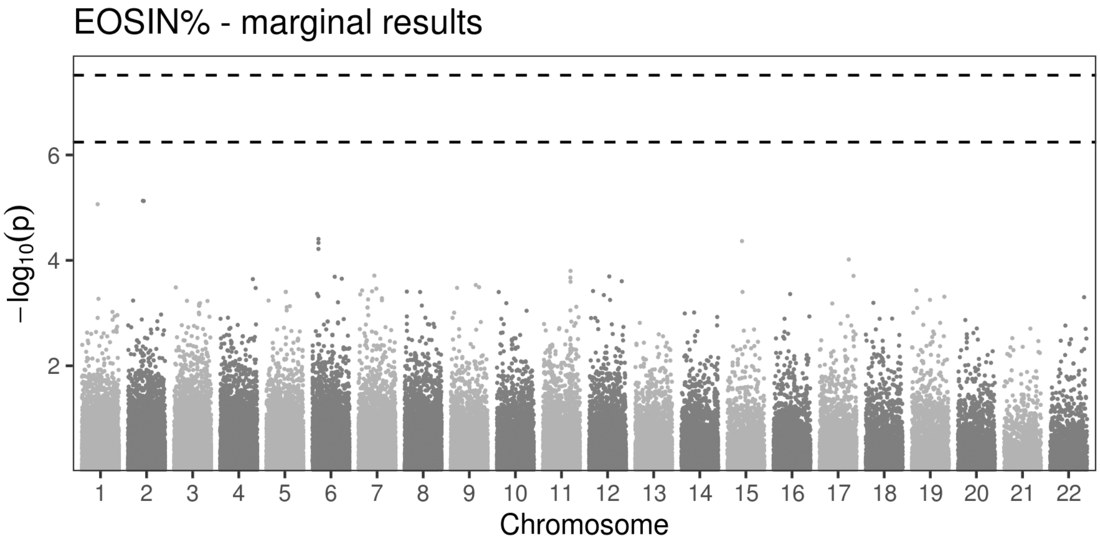
EOSIN% - marginal results
−log10(p)
6
4
2
1 2 3 4 5 6 7 8 9 10 11 12 13 14 15 16 17 18 19 20 21 22
Chromosome

(O)

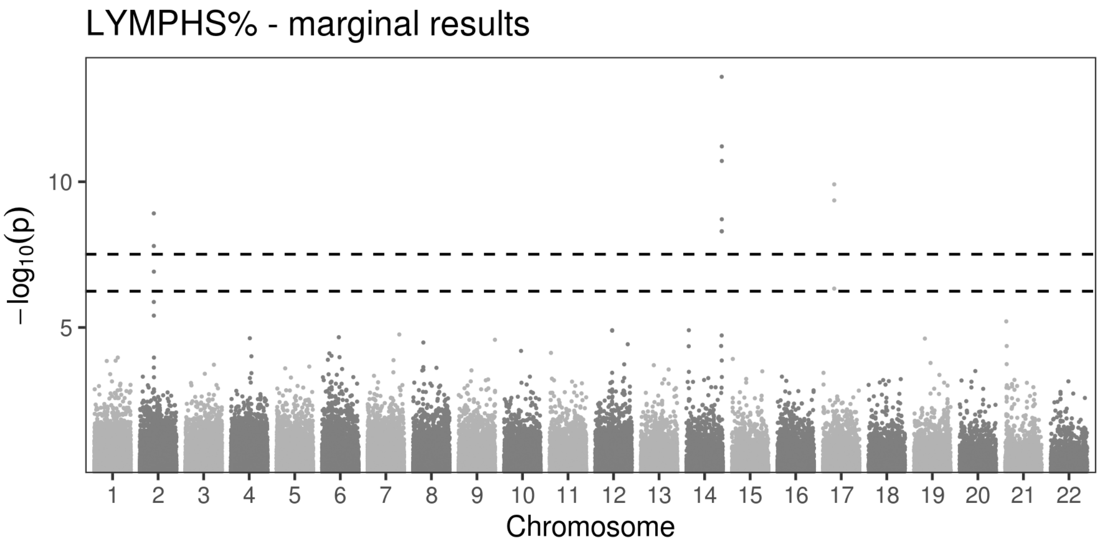
LYMPHS% - marginal results
−log10(p)
Chromosome

(P)

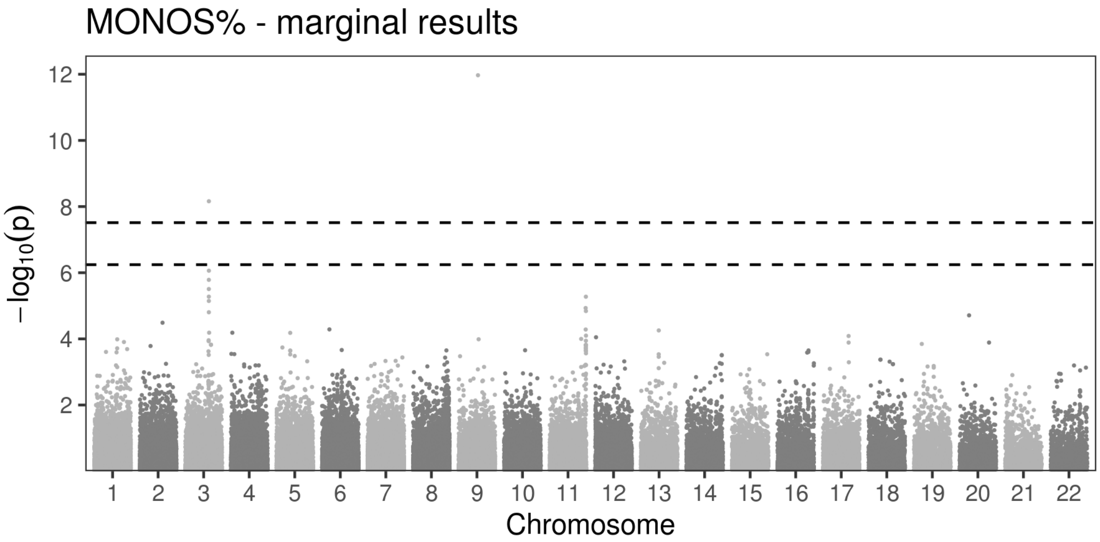
MONOS% - marginal results
−log10(p)
Chromosome

(Q)

NEUTRO% - marginal results

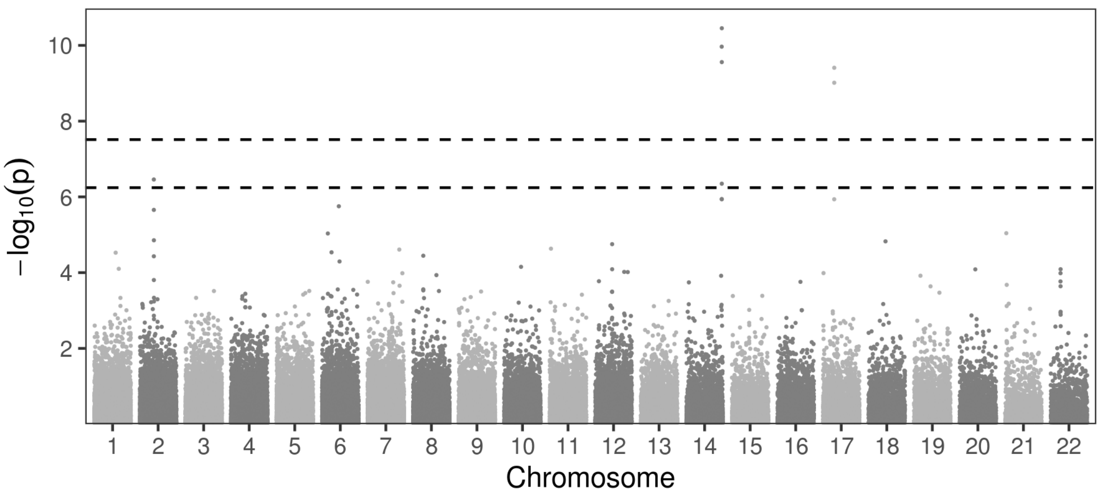

(R)

MPV - marginal results

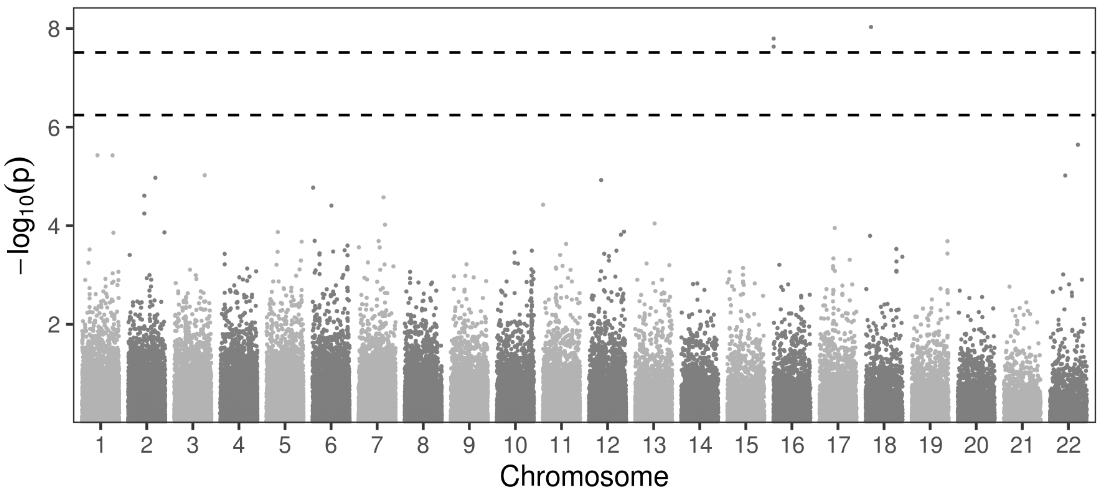

(S)

PLT - marginal results

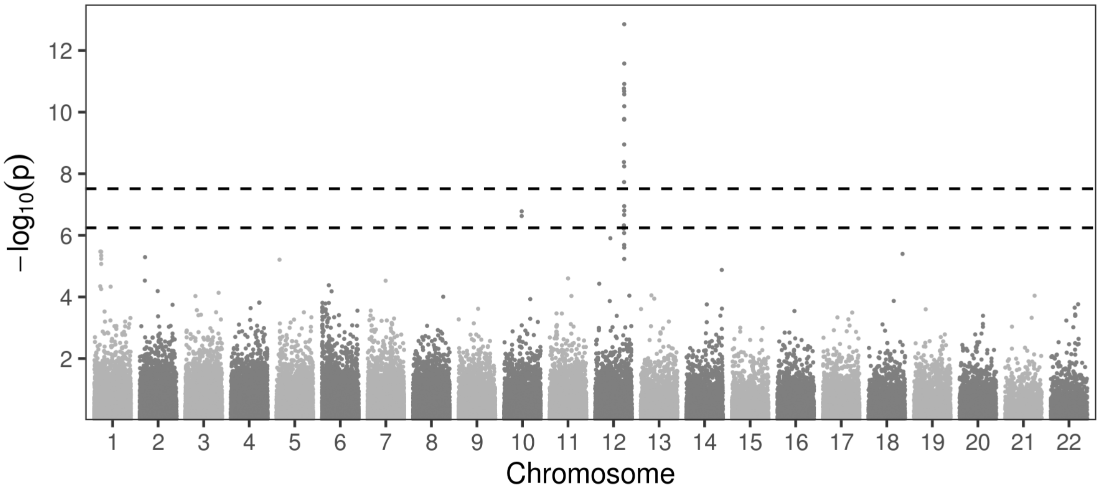

(T)

FERRITIN - marginal results

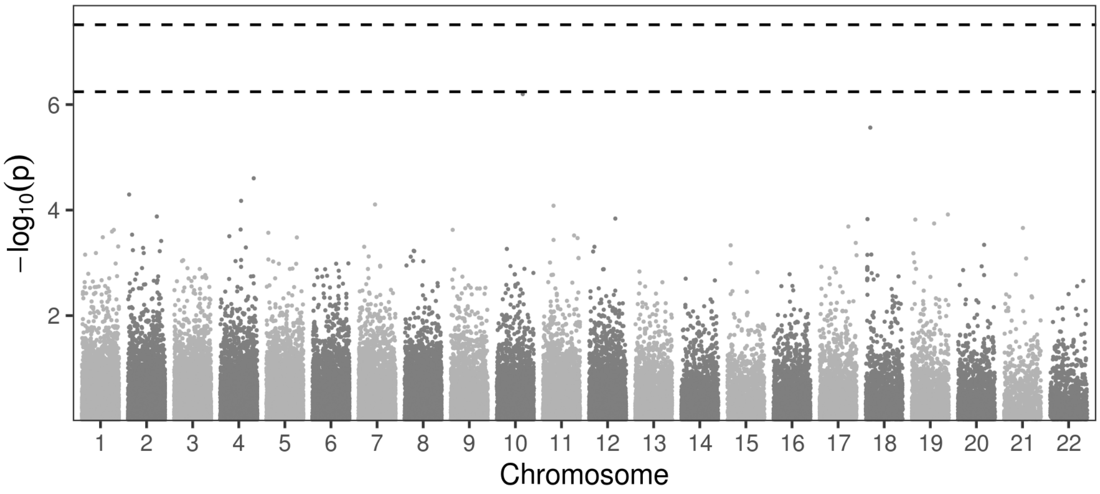

(U)

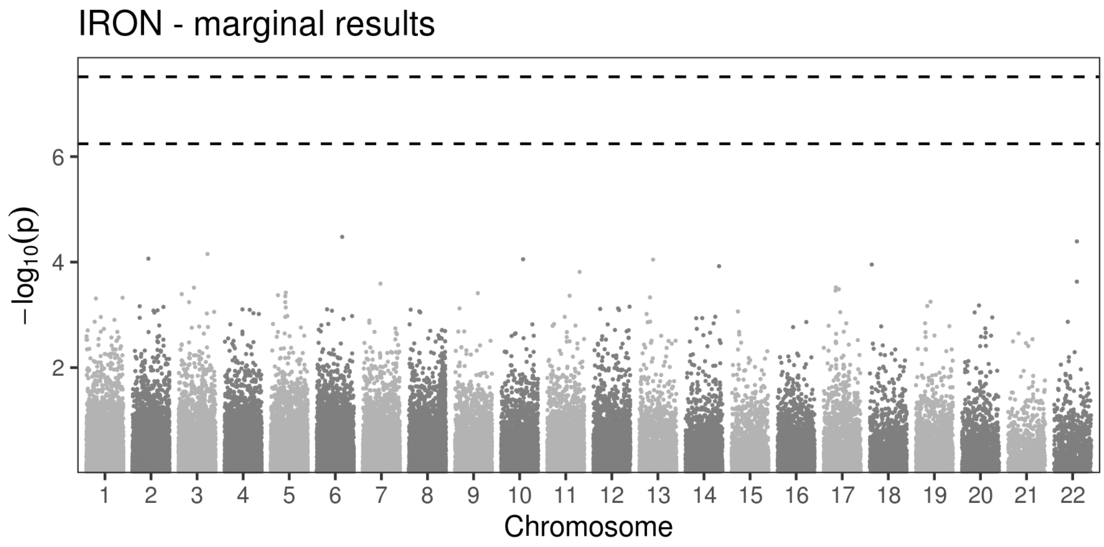
IRON - marginal results
−log10(p)
2
4
6
1 2 3 4 5 6 7 8 9 10 11 12 13 14 15 16 17 18 19 20 21 22
Chromosome

(V)

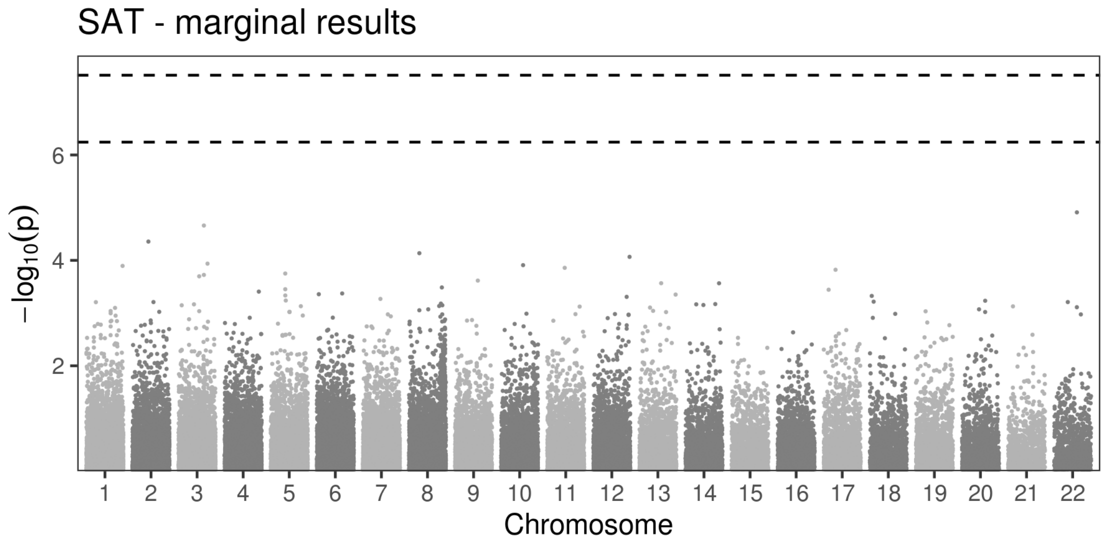
SAT - marginal results
−log10(p)
2
4
6
1 2 3 4 5 6 7 8 9 10 11 12 13 14 15 16 17 18 19 20 21 22
Chromosome

(W)

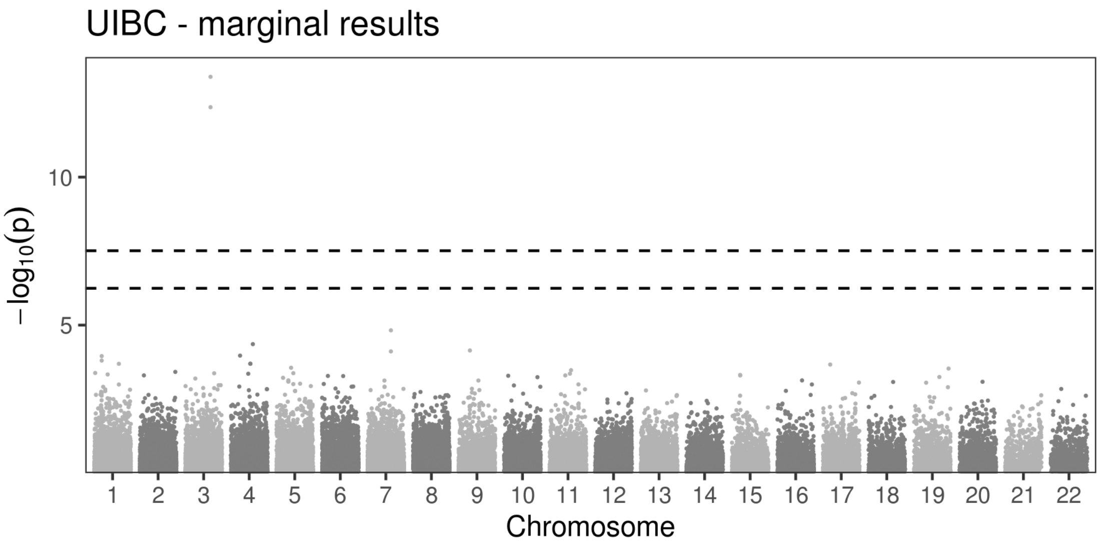
UIBC - marginal results
−log10(p)
10
5
Chromosome

(X)

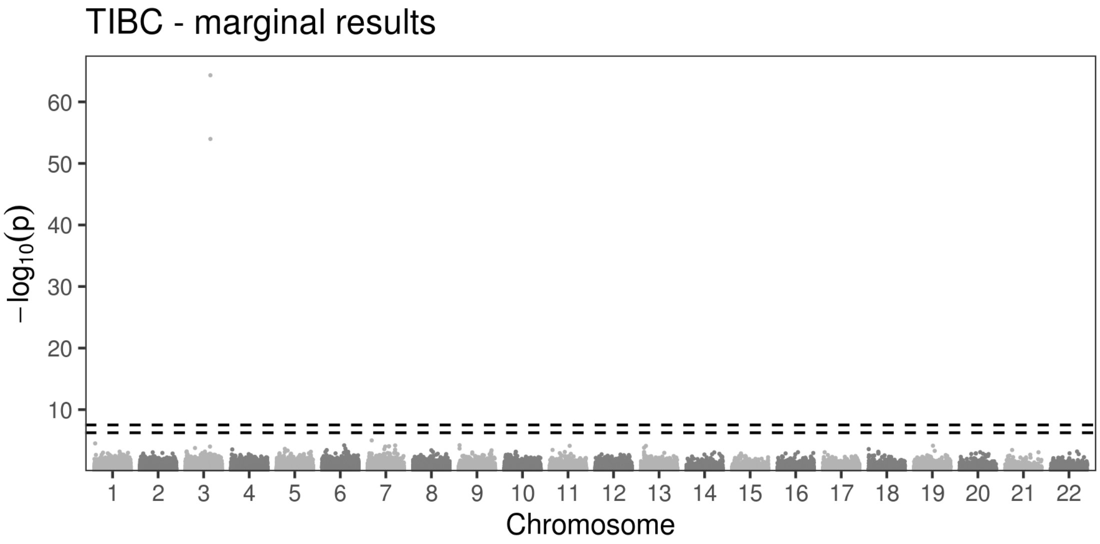
TIBC - marginal results
−log10(p)
60
50
40
30
20
10
Chromosome

**Figure S3**

**A)**

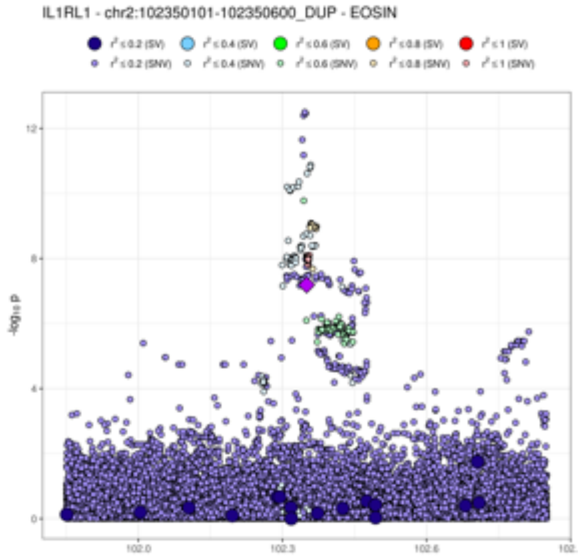
IL1RL1 - chr2:102350101-102350600_DUP - EOSIN
$r^2 \le 0.2$ (SV)
$r^2 \le 0.4$ (SV)
$r^2 \le 0.6$ (SV)
$r^2 \le 0.8$ (SV)
$r^2 \le 1$ (SV)
$r^2 \le 0.2$ (SNV)
$r^2 \le 0.4$ (SNV)
$r^2 \le 0.6$ (SNV)
$r^2 \le 0.8$ (SNV)
$r^2 \le 1$ (SNV)
$-\log_{10} p$
12
8
4
0
102.0
102.3
102.6

**B)**

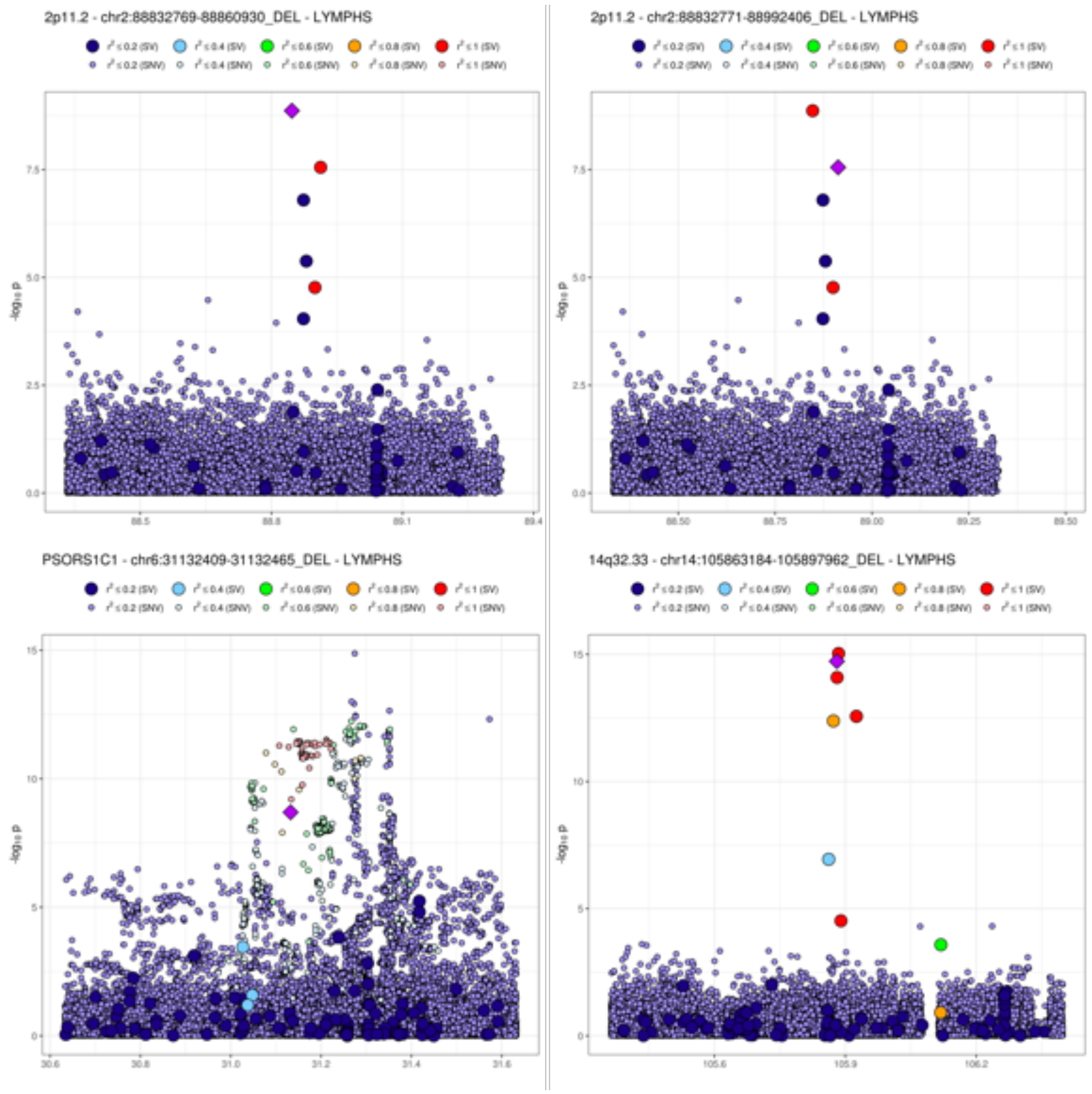
2p11.2 - chr2:88832769-88860930_DEL - LYMPHS
-log10 p
2p11.2 - chr2:88832771-88992406_DEL - LYMPHS
-log10 p
PSORS1C1 - chr6:31132409-31132465_DEL - LYMPHS
-log10 p
14q32.33 - chr14:105863184-105897962_DEL - LYMPHS
-log10 p

**C)**

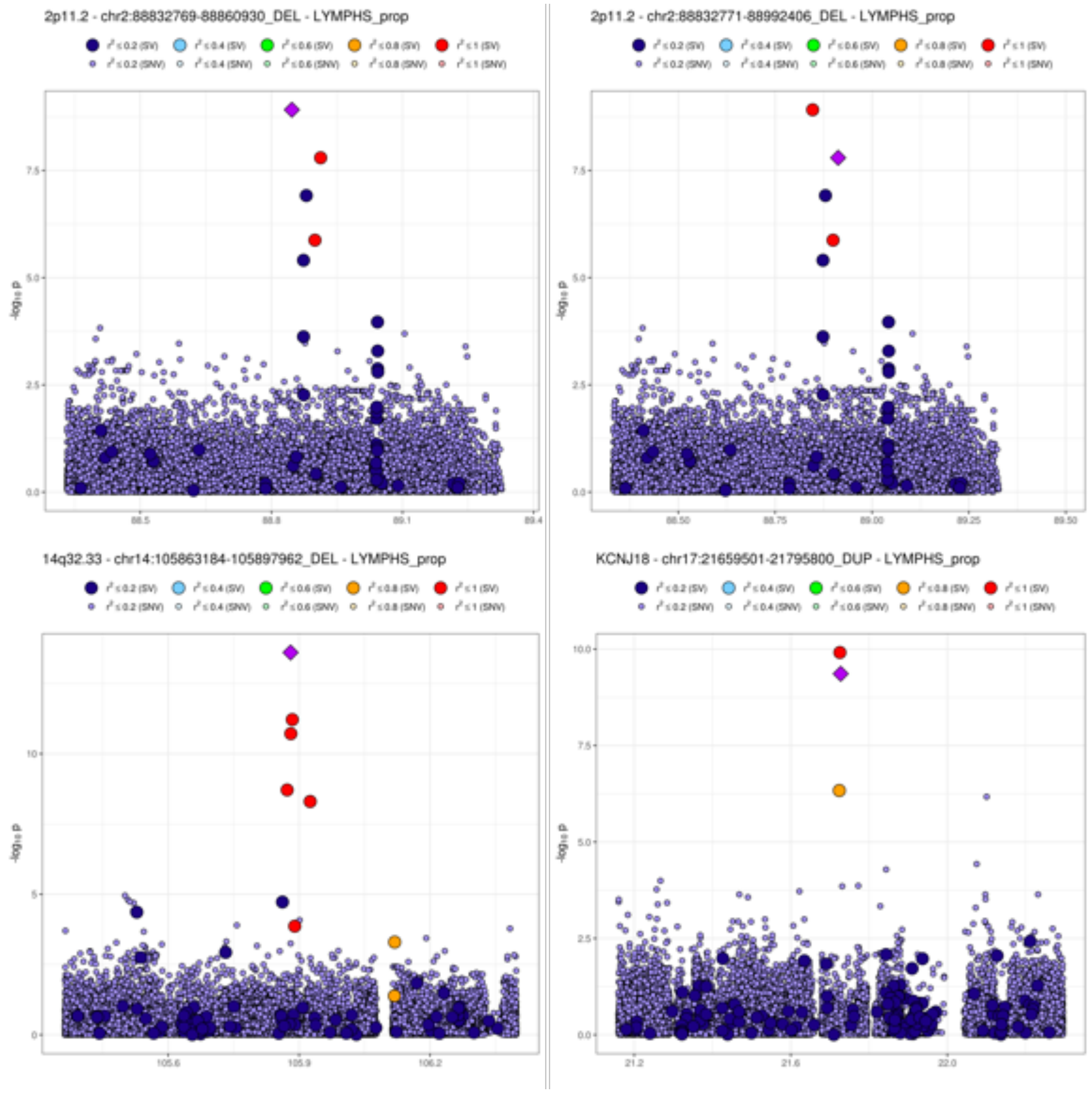
2p11.2 - chr2:88832769-88860930_DEL - LYMPHS_prop
2p11.2 - chr2:88832771-88992406_DEL - LYMPHS_prop
14q32.33 - chr14:105863184-105897962_DEL - LYMPHS_prop
KCNJ18 - chr17:21659501-21795800_DUP - LYMPHS_prop
-log10 p

**D)**

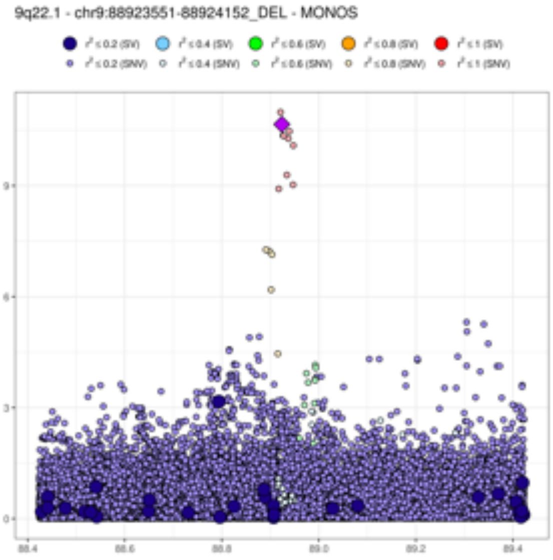
9q22.1 - chr9:88923551-88924152_DEL - MONOS
-log10 p

**E)**

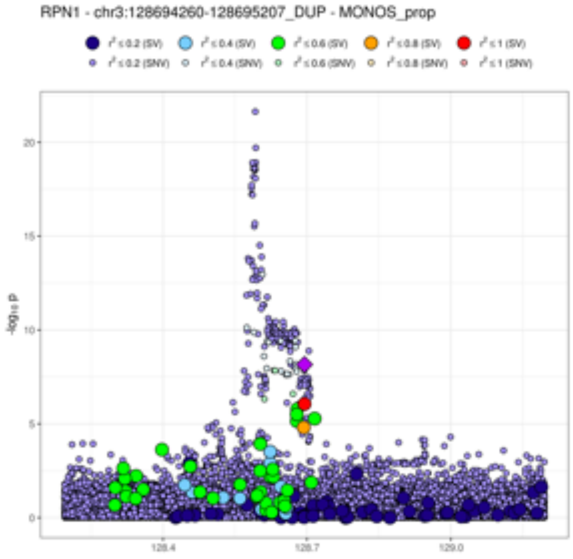
RPN1 - chr3:128694260-128695207_DUP - MONOS_prop
-log10 p

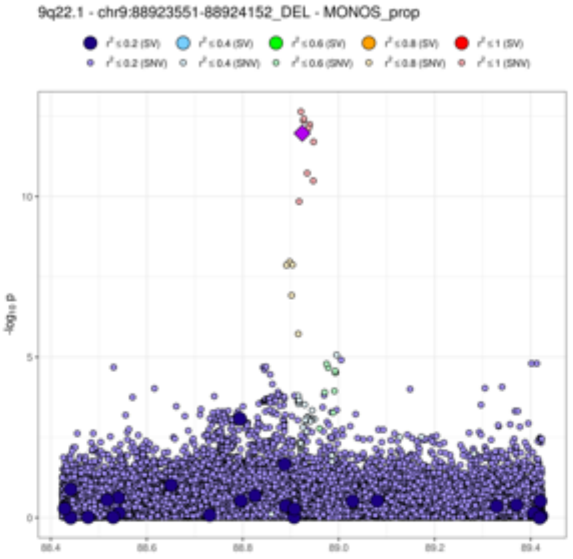
9q22.1 - chr9:88923551-88924152_DEL - MONOS_prop
-log10 p

**F)**

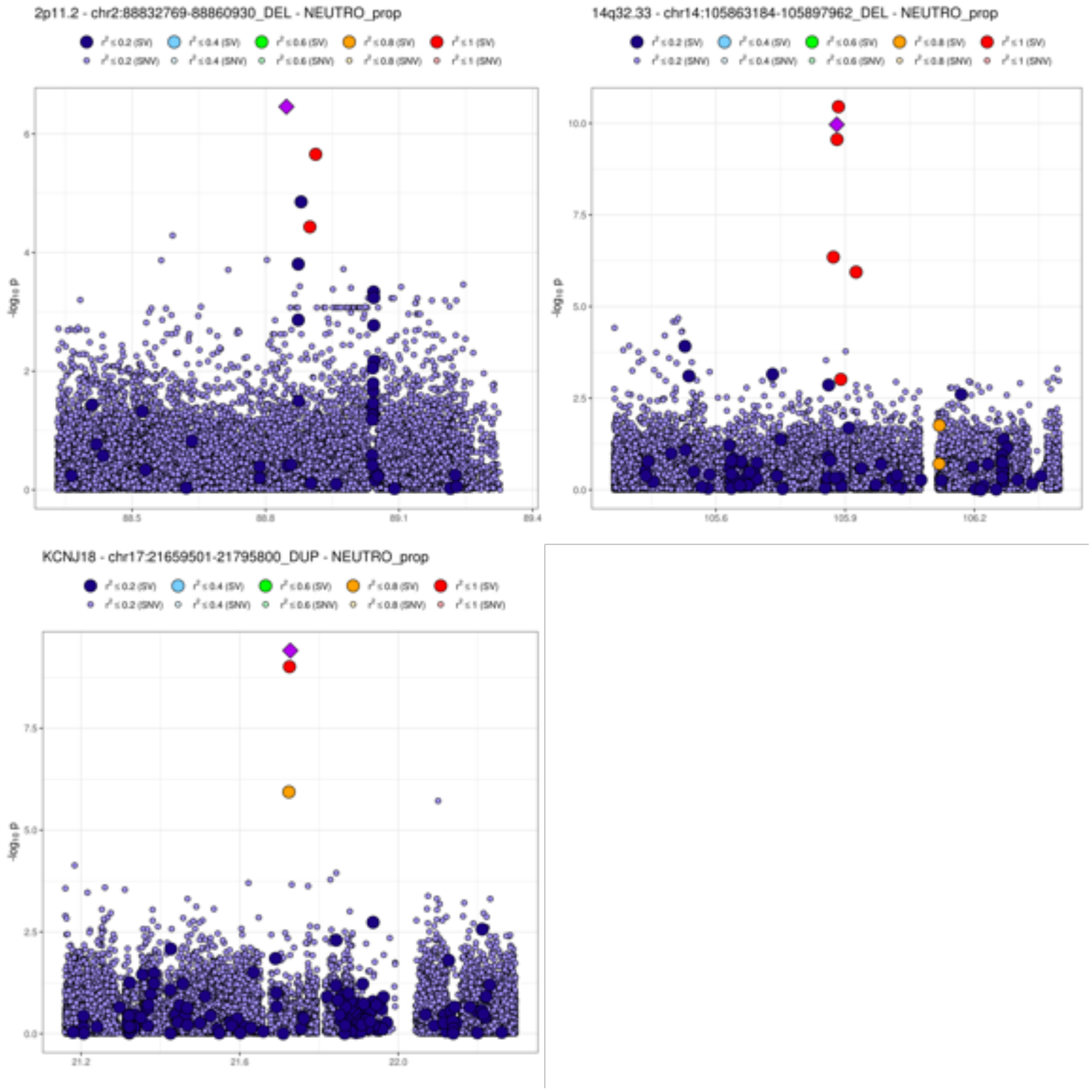
2p11.2 - chr2:88832769-88860930_DEL - NEUTRO_prop
14q32.33 - chr14:105863184-105897962_DEL - NEUTRO_prop
KCNJ18 - chr17:21659501-21795800_DUP - NEUTRO_prop
-log₁₀ p

**G)**

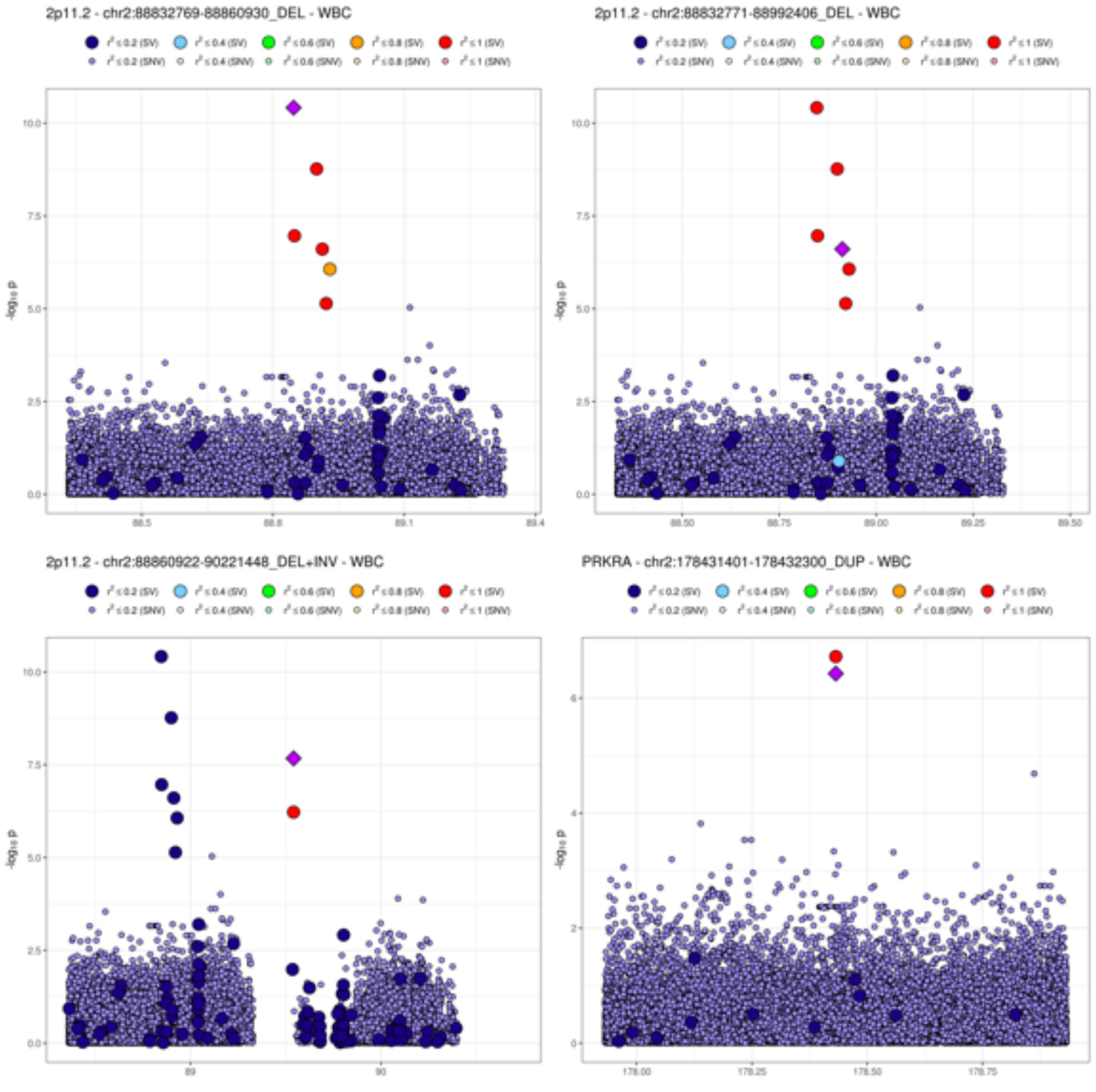
2p11.2 - chr2:88832769-88860930_DEL - WBC
2p11.2 - chr2:88832771-88992406_DEL - WBC
2p11.2 - chr2:88860922-90221448_DEL+INV - WBC
PRKRA - chr2:178431401-178432300_DUP - WBC
$r^2 \le 0.2$ (SV)
$r^2 \le 0.4$ (SV)
$r^2 \le 0.6$ (SV)
$r^2 \le 0.8$ (SV)
$r^2 \le 1$ (SV)
$r^2 \le 0.2$ (SNV)
$r^2 \le 0.4$ (SNV)
$r^2 \le 0.6$ (SNV)
$r^2 \le 0.8$ (SNV)
$r^2 \le 1$ (SNV)
$-\log_{10} p$

6p21.33 - chr6:31417422-31417499_DEL - WBC

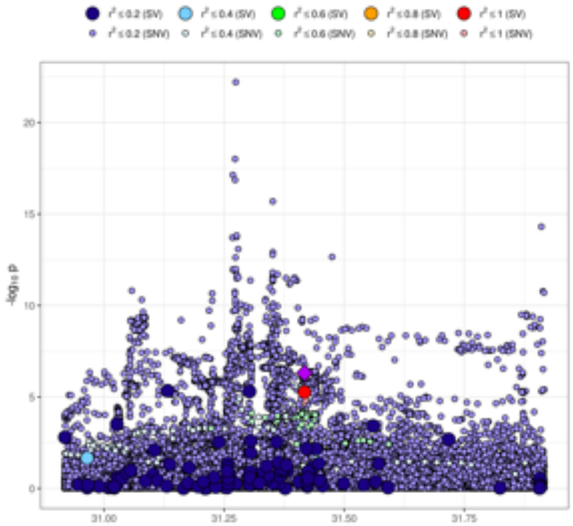

6p21.32 - chr6:32410098-32410877_DEL - WBC

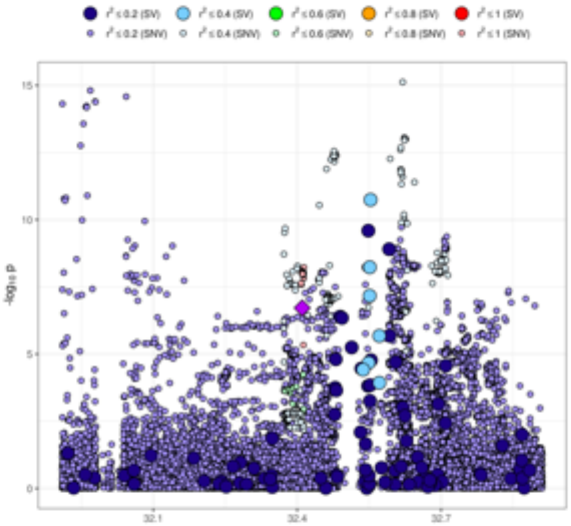

6p21.32 - chr6:32493182-32493439_DEL - WBC

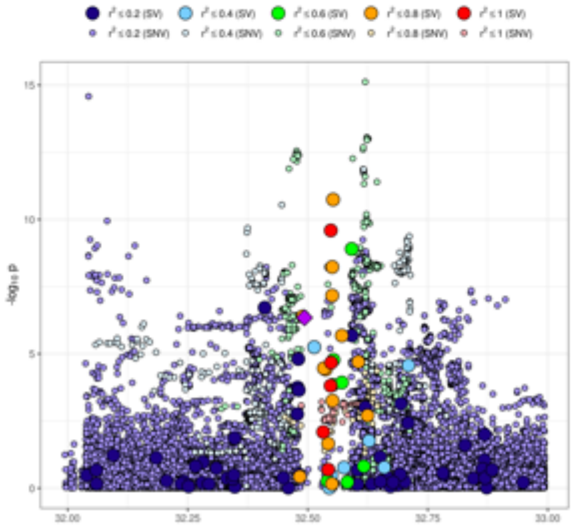

6p21.32 - chr6:32591559-32591660_DEL - WBC

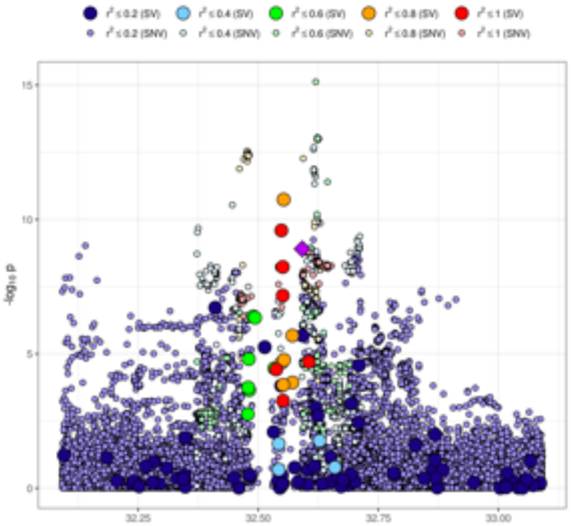

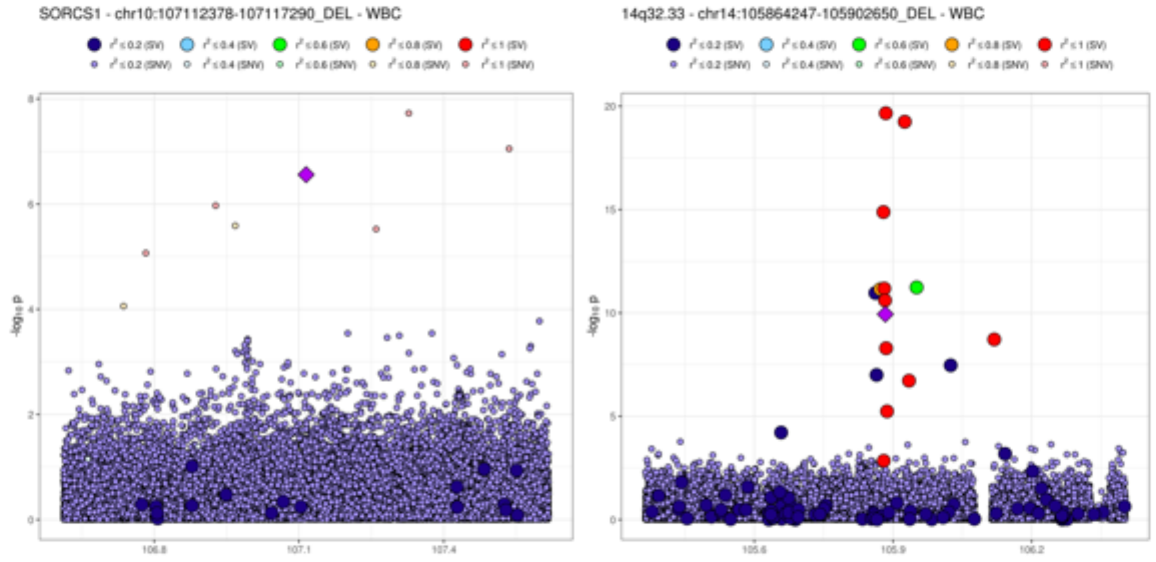
SORCS1 - chr10:107112378-107117290_DEL - WBC
14q32.33 - chr14:105864247-105902650_DEL - WBC
-log10 p
106.8
107.1
107.4
105.6
105.9
106.2

H)

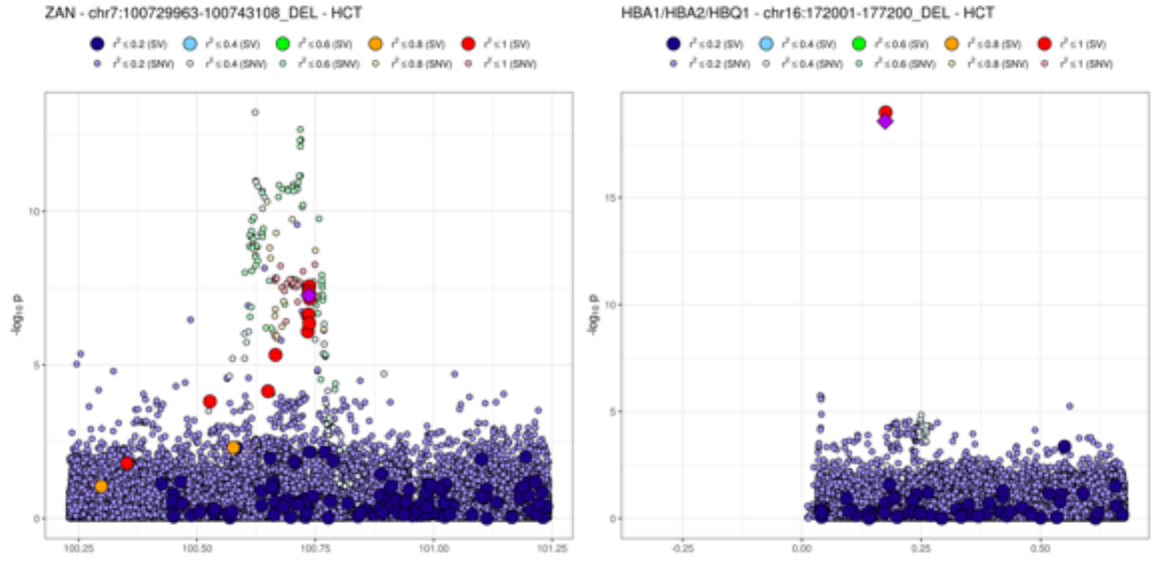
ZAN - chr7:100729963-100743108_DEL - HCT
HBA1/HBA2/HBQ1 - chr16:172001-177200_DEL - HCT
-log10 p
100.25
100.50
100.75
101.00
101.25
-0.25
0.00
0.25
0.50

**I)**

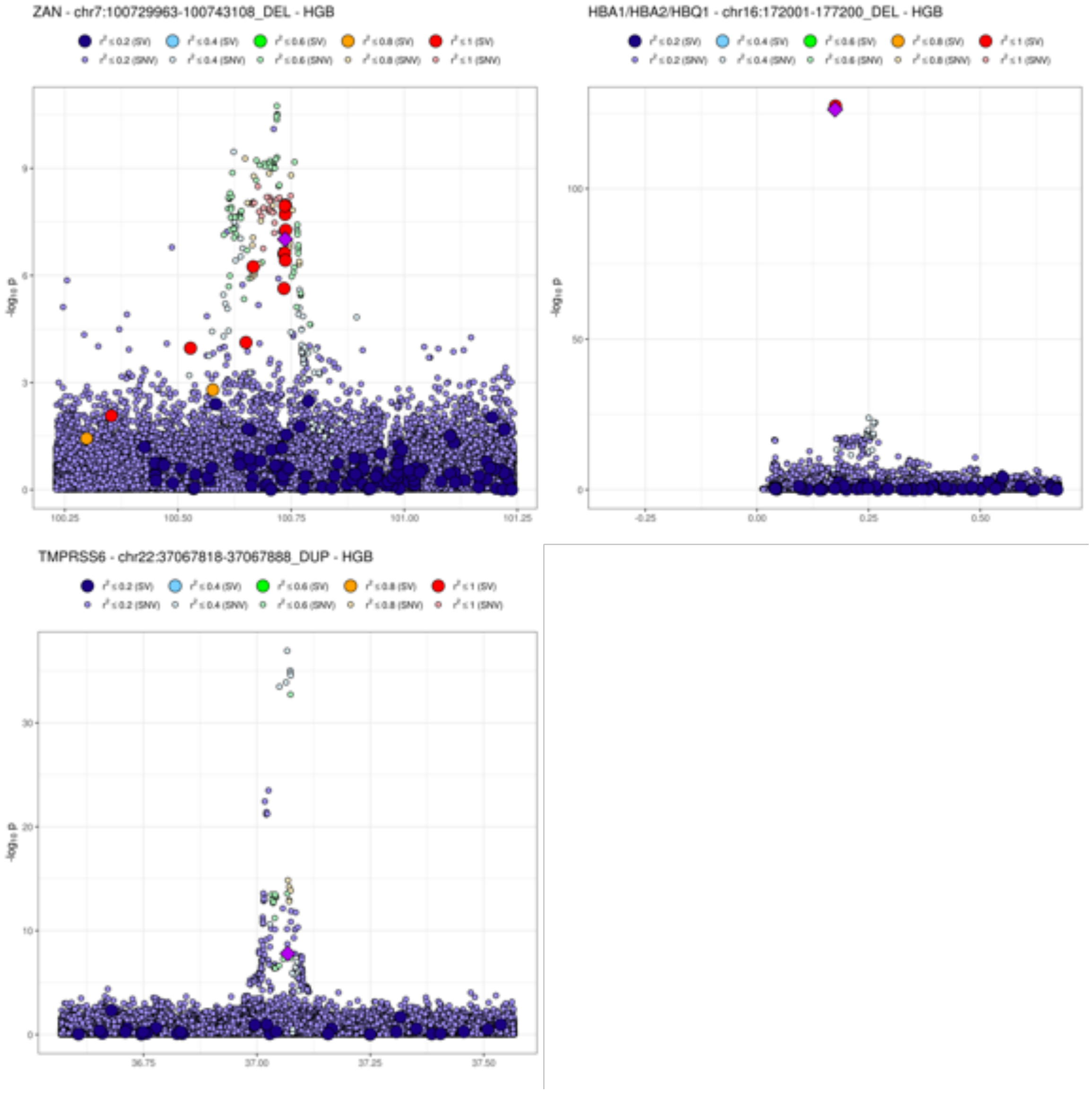
ZAN - chr7:100729963-100743108_DEL - HGB
r² ≤ 0.2 (SV)
r² ≤ 0.4 (SV)
r² ≤ 0.6 (SV)
r² ≤ 0.8 (SV)
r² ≤ 1 (SV)
r² ≤ 0.2 (SNV)
r² ≤ 0.4 (SNV)
r² ≤ 0.6 (SNV)
r² ≤ 0.8 (SNV)
r² ≤ 1 (SNV)
-log10 p
0
3
6
9
100.25
100.50
100.75
101.00
101.25
HBA1/HBA2/HBQ1 - chr16:172001-177200_DEL - HGB
0
50
100
-0.25
0.00
0.25
0.50
TMPRSS6 - chr22:37067818-37067888_DUP - HGB
0
10
20
30
36.75
37.00
37.25
37.50

J)

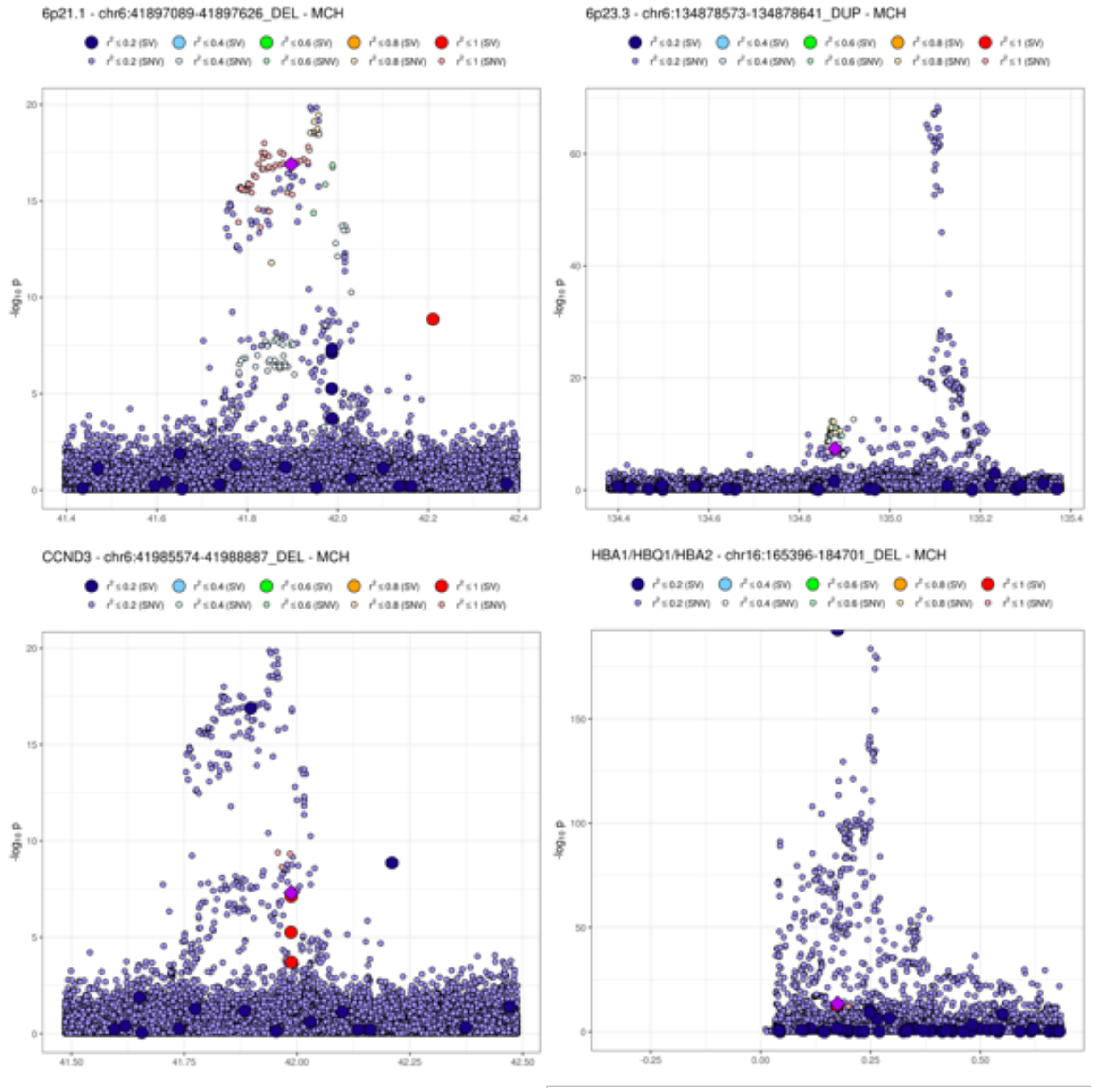
6p21.1 - chr6:41897089-41897626_DEL - MCH
6p23.3 - chr6:134878573-134878641_DUP - MCH
CCND3 - chr6:41985574-41988887_DEL - MCH
HBA1/HBQ1/HBA2 - chr16:165396-184701_DEL - MCH
r² ≤ 0.2 (SV)
r² ≤ 0.4 (SV)
r² ≤ 0.6 (SV)
r² ≤ 0.8 (SV)
r² ≤ 1 (SV)
r² ≤ 0.2 (SNV)
r² ≤ 0.4 (SNV)
r² ≤ 0.6 (SNV)
r² ≤ 0.8 (SNV)
r² ≤ 1 (SNV)
-log10 p

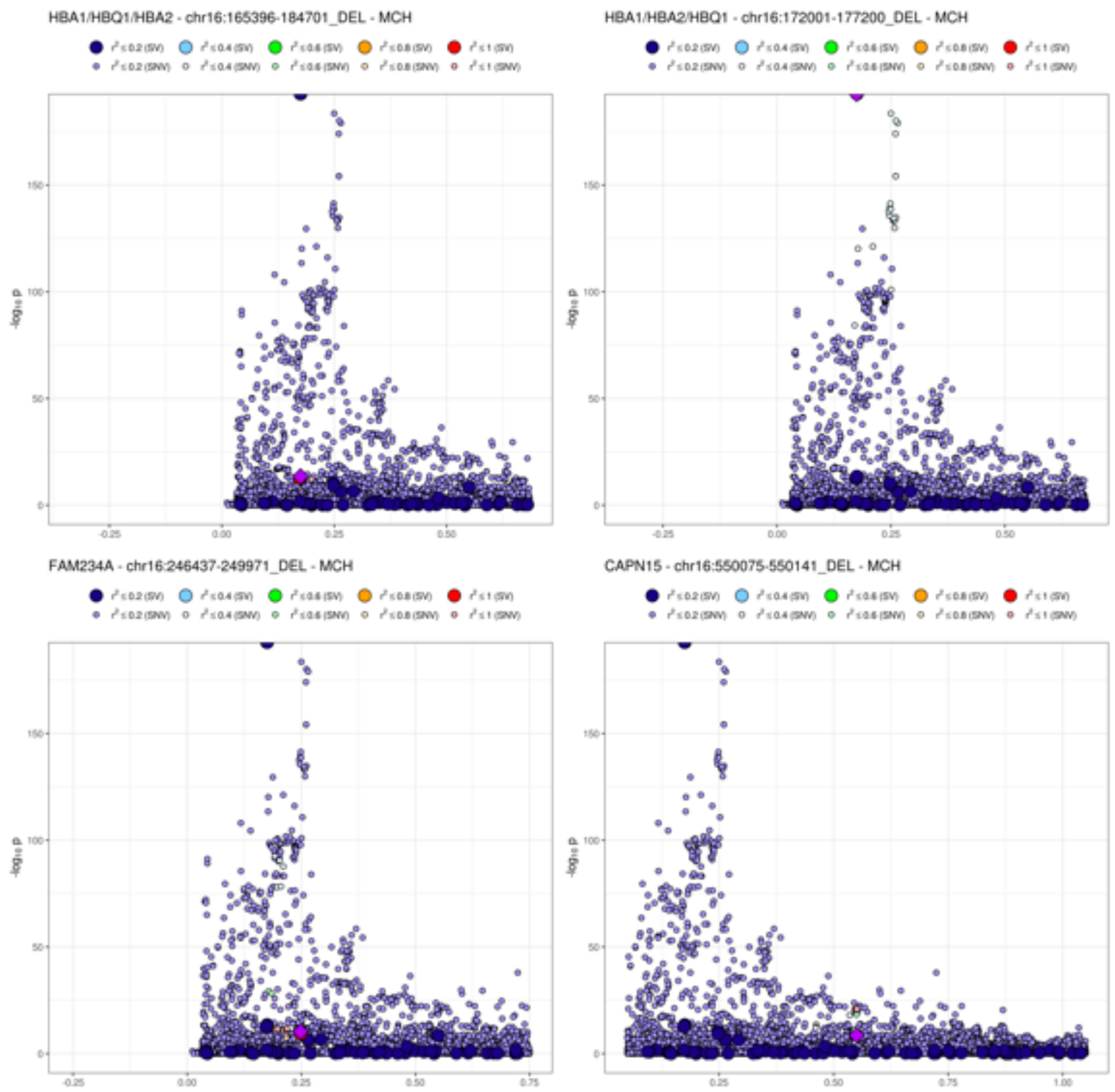
HBA1/HBQ1/HBA2 - chr16:165396-184701_DEL - MCH
HBA1/HBA2/HBQ1 - chr16:172001-177200_DEL - MCH
FAM234A - chr16:246437-249971_DEL - MCH
CAPN15 - chr16:550075-550141_DEL - MCH
r² ≤ 0.2 (SV)
r² ≤ 0.4 (SV)
r² ≤ 0.6 (SV)
r² ≤ 0.8 (SV)
r² ≤ 1 (SV)
r² ≤ 0.2 (SNV)
r² ≤ 0.4 (SNV)
r² ≤ 0.6 (SNV)
r² ≤ 0.8 (SNV)
r² ≤ 1 (SNV)
-log10 p
0
50
100
150
-0.25
0.00
0.25
0.50
0.75
1.00

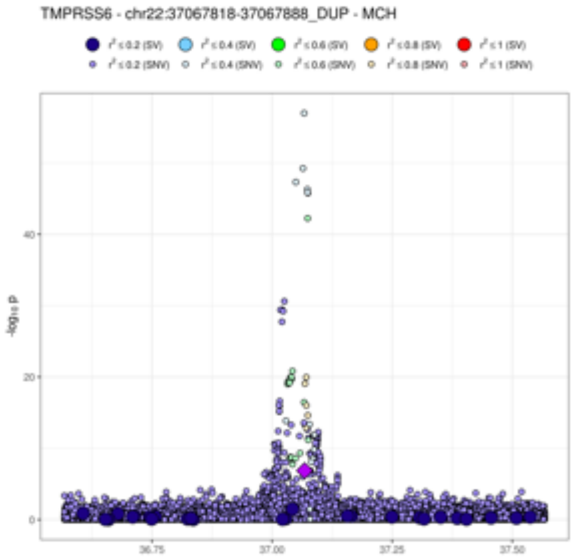
TMPRSS6 - chr22:37067818-37067888_DUP - MCH
r² ≤ 0.2 (SV) r² ≤ 0.4 (SV) r² ≤ 0.6 (SV) r² ≤ 0.8 (SV) r² ≤ 1 (SV)
r² ≤ 0.2 (SNV) r² ≤ 0.4 (SNV) r² ≤ 0.6 (SNV) r² ≤ 0.8 (SNV) r² ≤ 1 (SNV)
-log10 P
0
20
40
36.75
37.00
37.25
37.50

K)

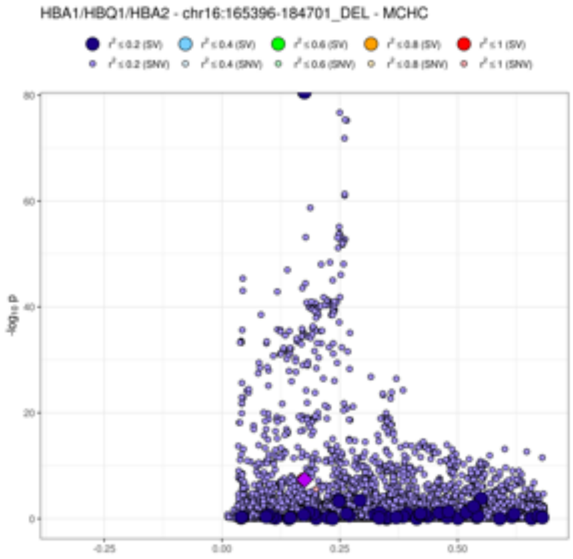
HBA1/HBQ1/HBA2 - chr16:165396-184701_DEL - MCHC
r² ≤ 0.2 (SV) r² ≤ 0.4 (SV) r² ≤ 0.6 (SV) r² ≤ 0.8 (SV) r² ≤ 1 (SV)
r² ≤ 0.2 (SNV) r² ≤ 0.4 (SNV) r² ≤ 0.6 (SNV) r² ≤ 0.8 (SNV) r² ≤ 1 (SNV)
-log10 P
0
20
40
60
80
-0.25
0.00
0.25
0.50

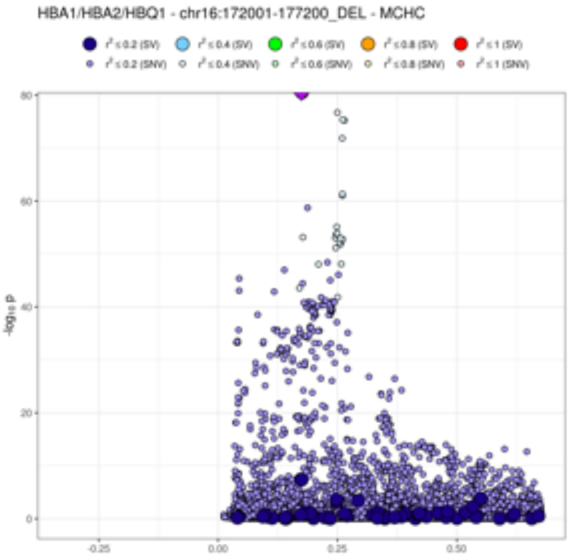
HBA1/HBA2/HBQ1 - chr16:172001-177200_DEL - MCHC
r² ≤ 0.2 (SV) r² ≤ 0.4 (SV) r² ≤ 0.6 (SV) r² ≤ 0.8 (SV) r² ≤ 1 (SV)
r² ≤ 0.2 (SNV) r² ≤ 0.4 (SNV) r² ≤ 0.6 (SNV) r² ≤ 0.8 (SNV) r² ≤ 1 (SNV)
-log10 P
0
20
40
60
80
-0.25
0.00
0.25
0.50

L)

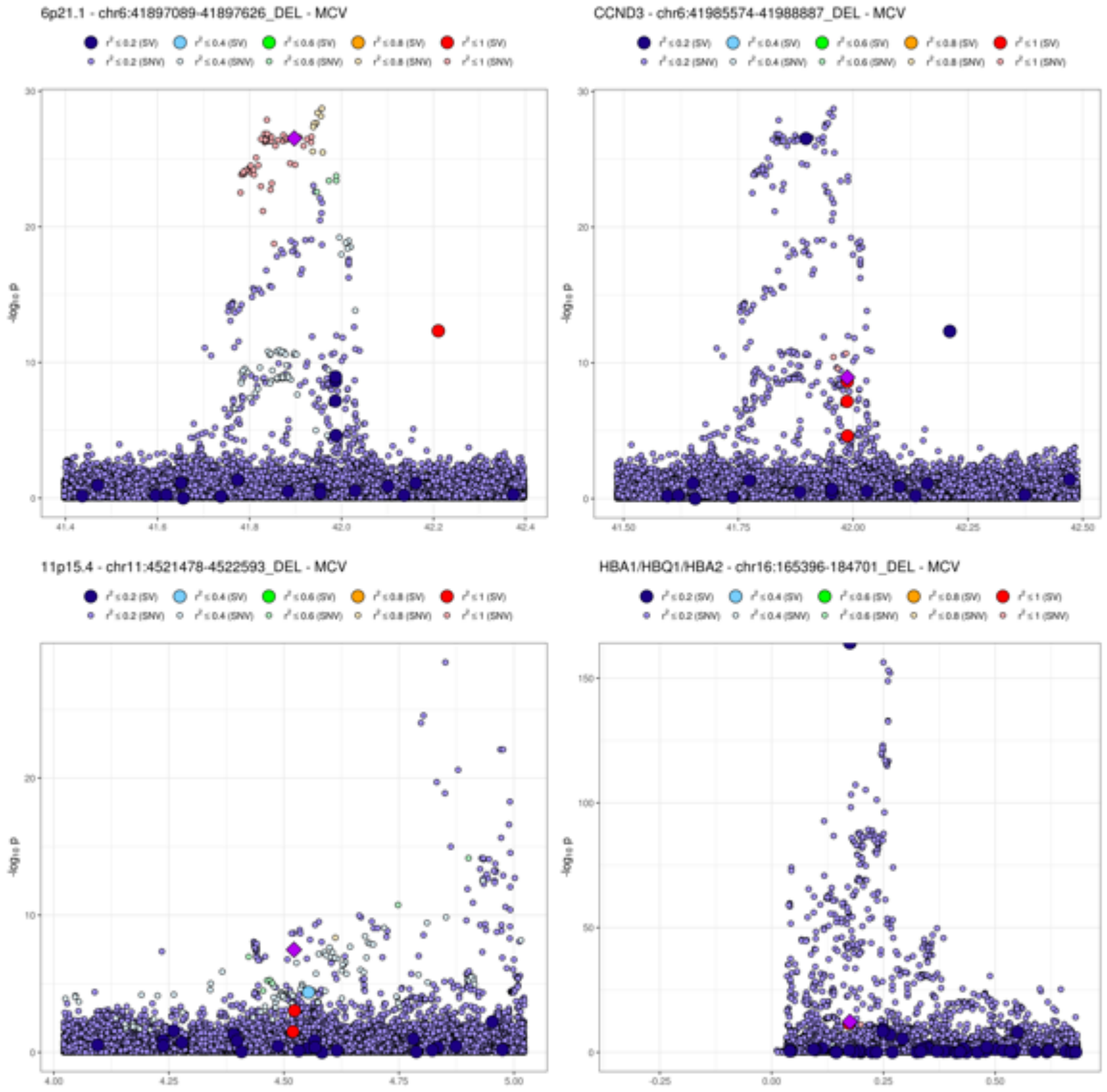
6p21.1 - chr6:41897089-41897626_DEL - MCV
CCND3 - chr6:41985574-41988887_DEL - MCV
11p15.4 - chr11:4521478-4522593_DEL - MCV
HBA1/HBQ1/HBA2 - chr16:165396-184701_DEL - MCV
r² ≤ 0.2 (SV)
r² ≤ 0.4 (SV)
r² ≤ 0.6 (SV)
r² ≤ 0.8 (SV)
r² ≤ 1 (SV)
r² ≤ 0.2 (SNV)
r² ≤ 0.4 (SNV)
r² ≤ 0.6 (SNV)
r² ≤ 0.8 (SNV)
r² ≤ 1 (SNV)
-log10 p

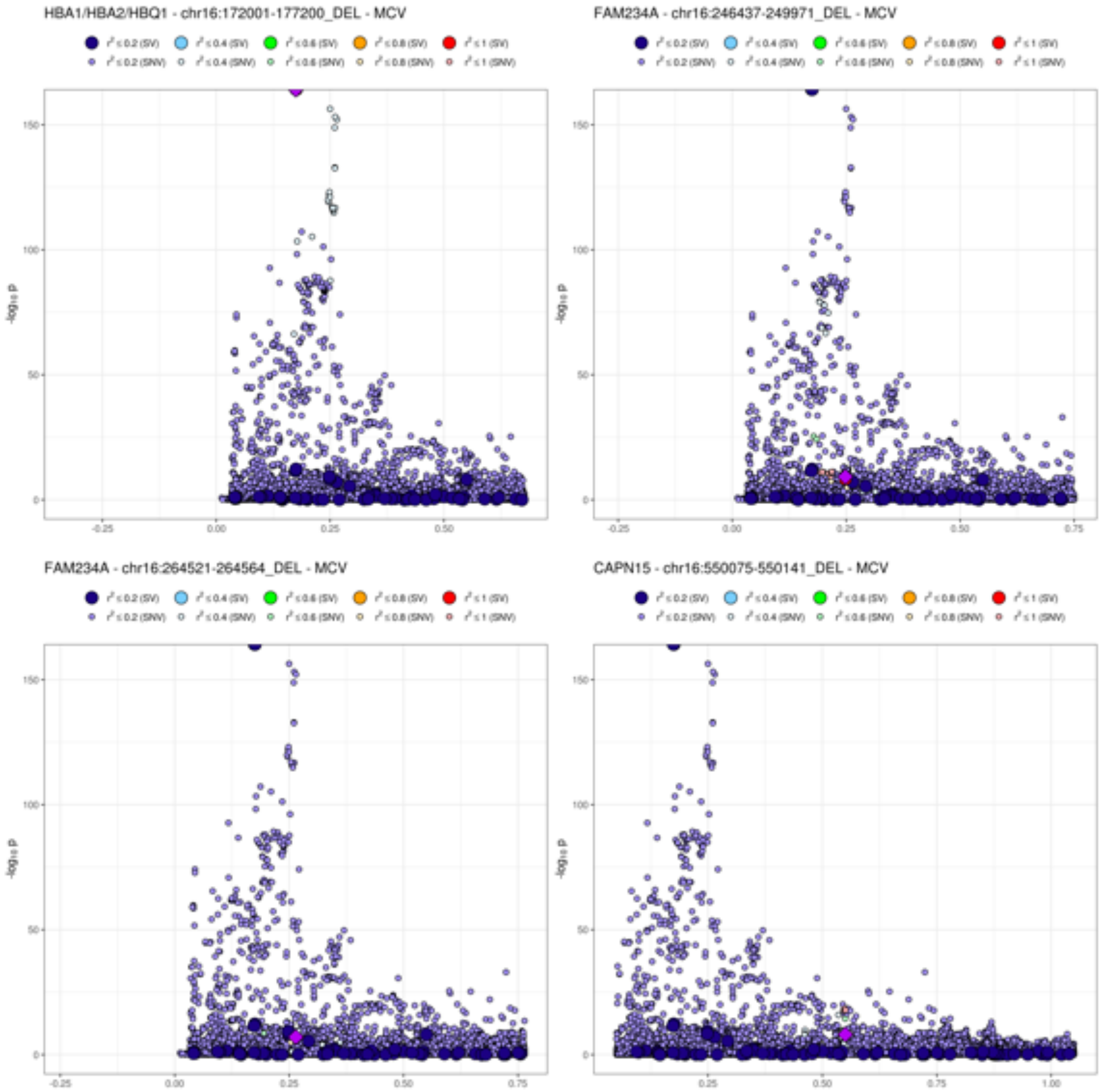
HBA1/HBA2/HBQ1 - chr16:172001-177200_DEL - MCV
FAM234A - chr16:246437-249971_DEL - MCV
FAM234A - chr16:264521-264564_DEL - MCV
CAPN15 - chr16:550075-550141_DEL - MCV
$r^2 \le 0.2$ (SV)
$r^2 \le 0.4$ (SV)
$r^2 \le 0.6$ (SV)
$r^2 \le 0.8$ (SV)
$r^2 \le 1$ (SV)
$r^2 \le 0.2$ (SNV)
$r^2 \le 0.4$ (SNV)
$r^2 \le 0.6$ (SNV)
$r^2 \le 0.8$ (SNV)
$r^2 \le 1$ (SNV)
$-\log_{10} p$

**M)**

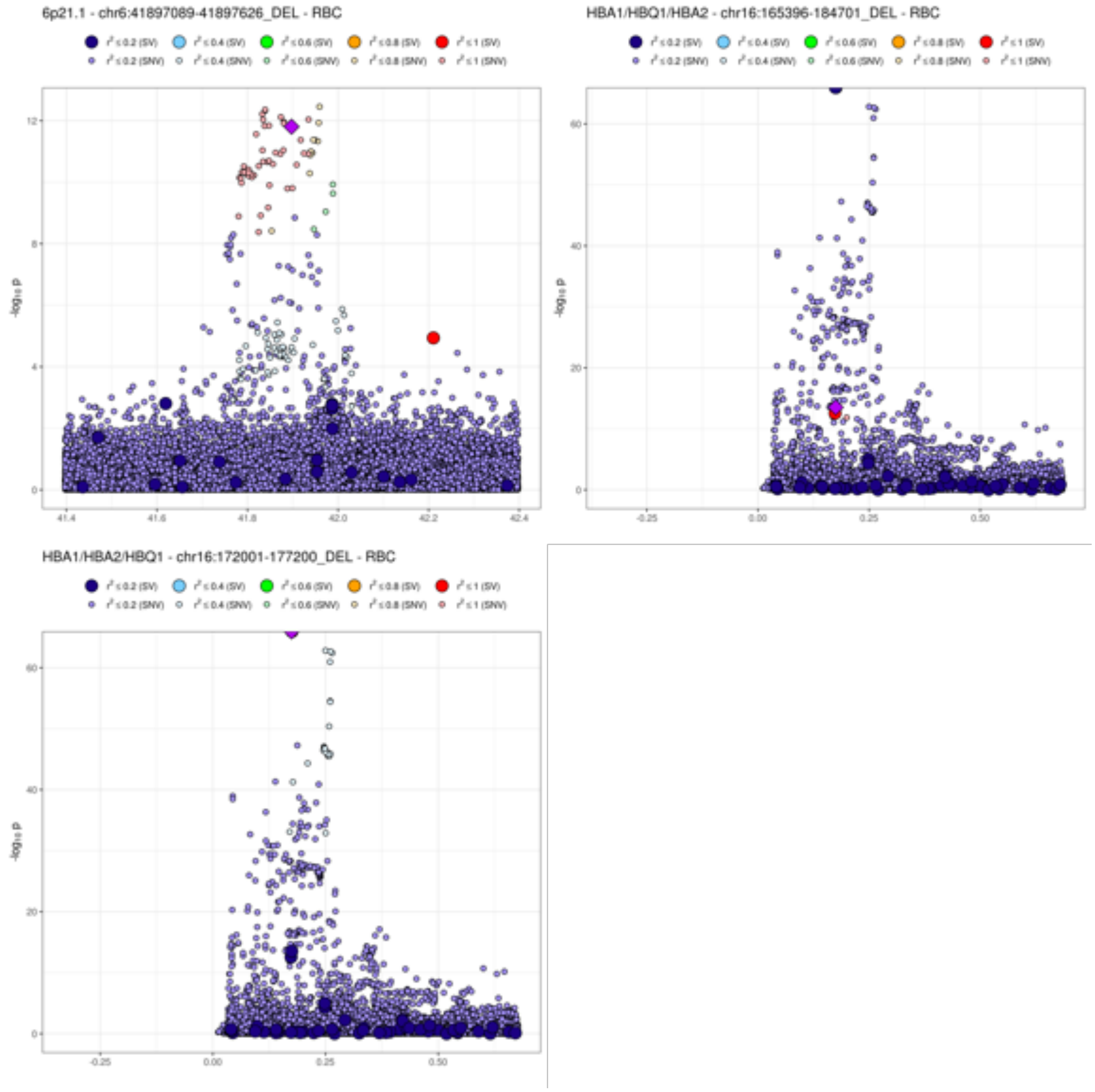
6p21.1 - chr6:41897089-41897626_DEL - RBC
HBA1/HBQ1/HBA2 - chr16:165396-184701_DEL - RBC
HBA1/HBA2/HBQ1 - chr16:172001-177200_DEL - RBC
r² ≤ 0.2 (SV)
r² ≤ 0.4 (SV)
r² ≤ 0.6 (SV)
r² ≤ 0.8 (SV)
r² ≤ 1 (SV)
r² ≤ 0.2 (SNV)
r² ≤ 0.4 (SNV)
r² ≤ 0.6 (SNV)
r² ≤ 0.8 (SNV)
r² ≤ 1 (SNV)
-log10 p

**N)**

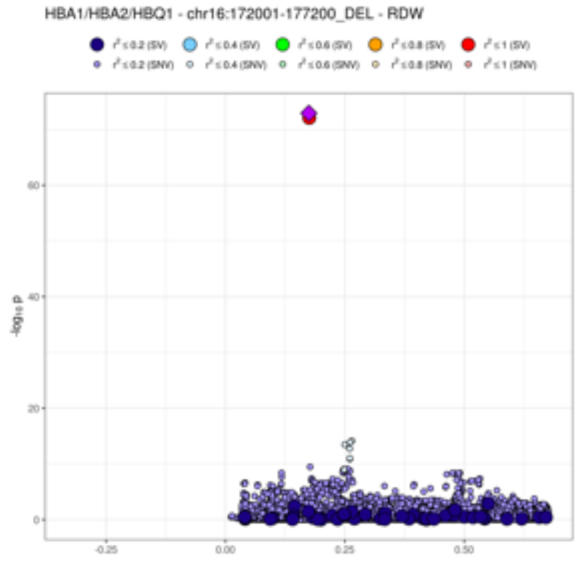
HBA1/HBA2/HBQ1 - chr16:172001-177200_DEL - RDW
-log10 p

O)

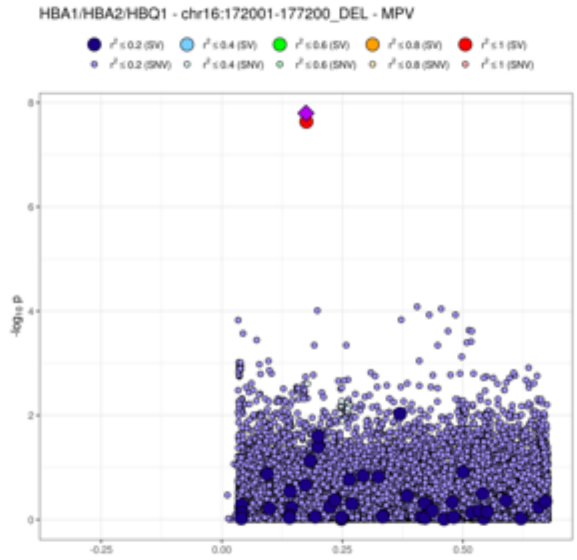
HBA1/HBA2/HBQ1 - chr16:172001-177200_DEL - MPV
-log10 p

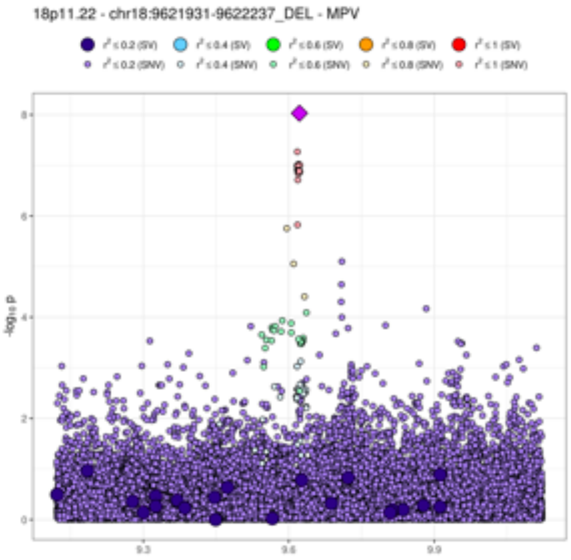
18p11.22 - chr18:9621931-9622237_DEL - MPV
-log10 p

P)

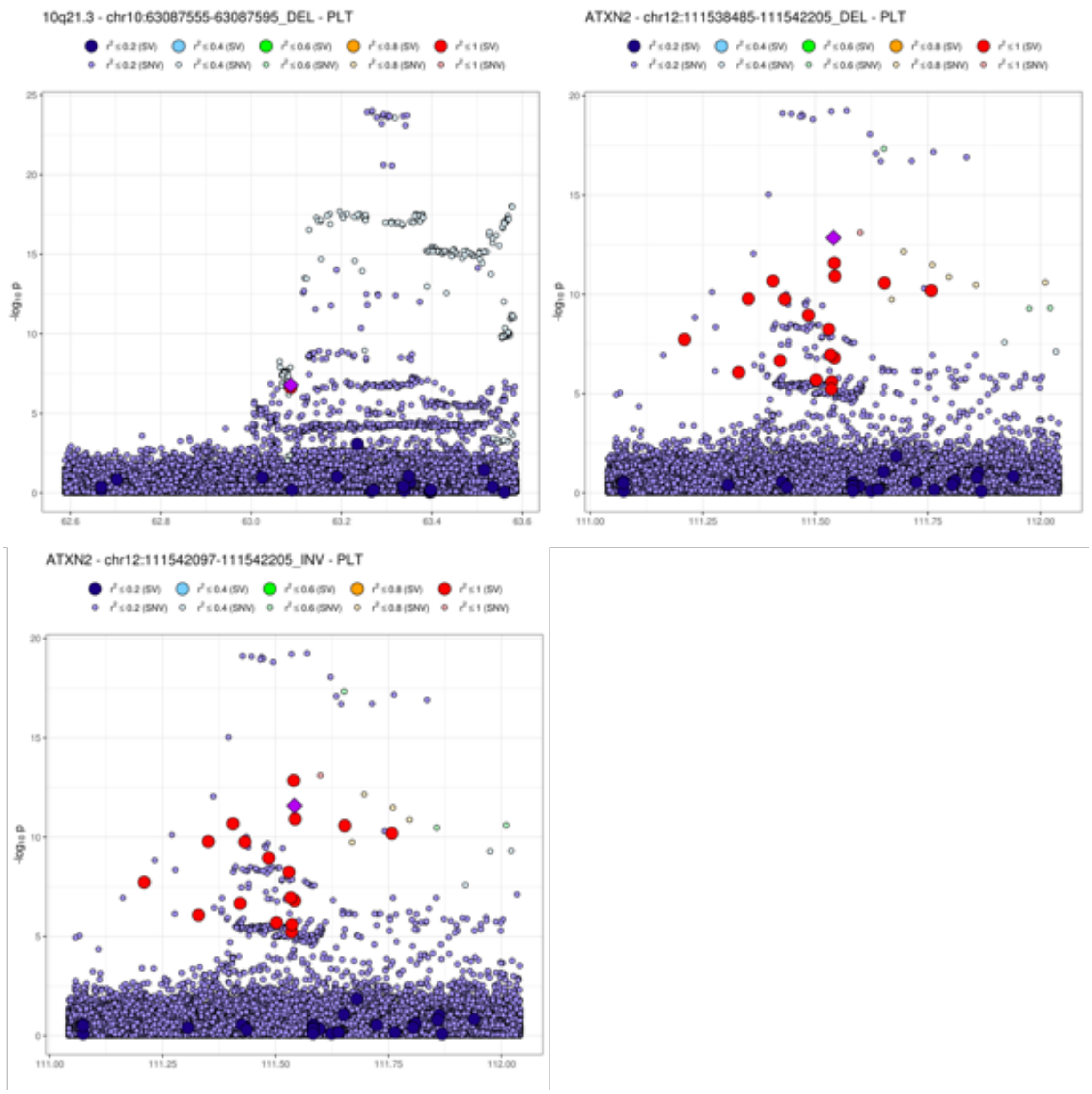
10q21.3 - chr10:63087555-63087595_DEL - PLT
ATXN2 - chr12:111538485-111542205_DEL - PLT
ATXN2 - chr12:111542097-111542205_INV - PLT
-log10 p

**Q)**

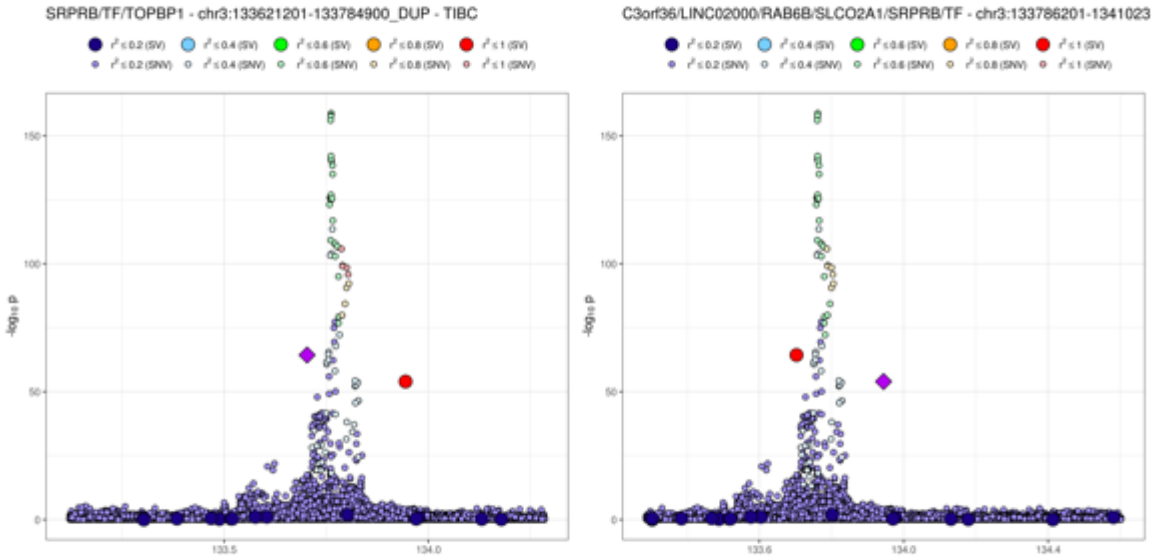
SRPRB/TF/TOPBP1 - chr3:133621201-133784900_DUP - TIBC
C3orf36/LINC02000/RAB6B/SLCO2A1/SRPRB/TF - chr3:133786201-1341023
-log10 p
150
100
50
0
133.5
134.0
133.6
134.4

R)

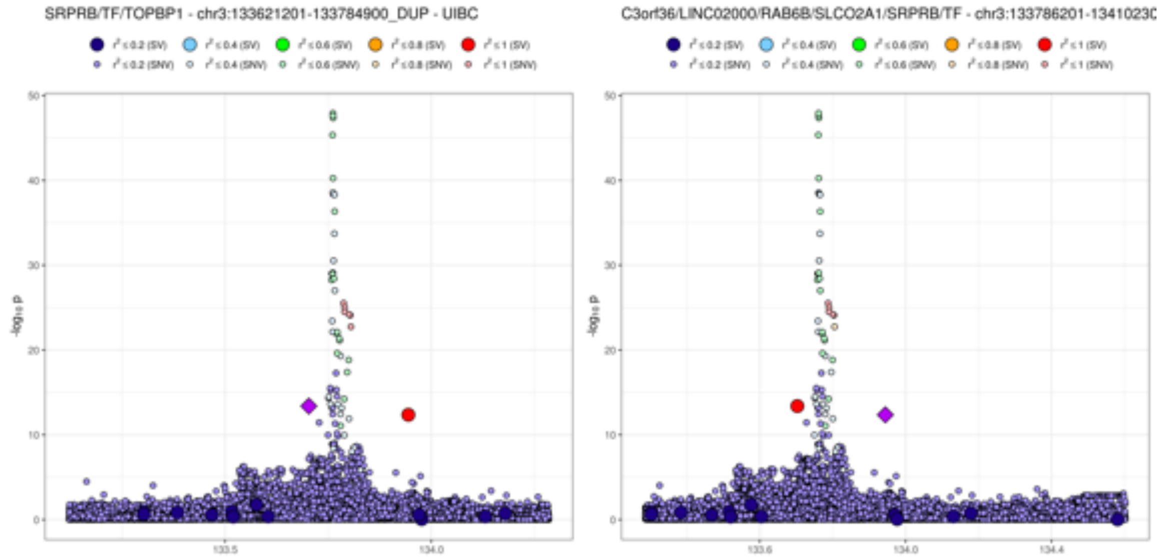
SRPRB/TF/TOPBP1 - chr3:133621201-133784900_DUP - UIBC
C3orf36/LINC02000/RAB6B/SLCO2A1/SRPRB/TF - chr3:133786201-13410230
-log10 p
50
40
30
20
10
0
133.5
134.0
133.6
134.4

**Figure S4.**

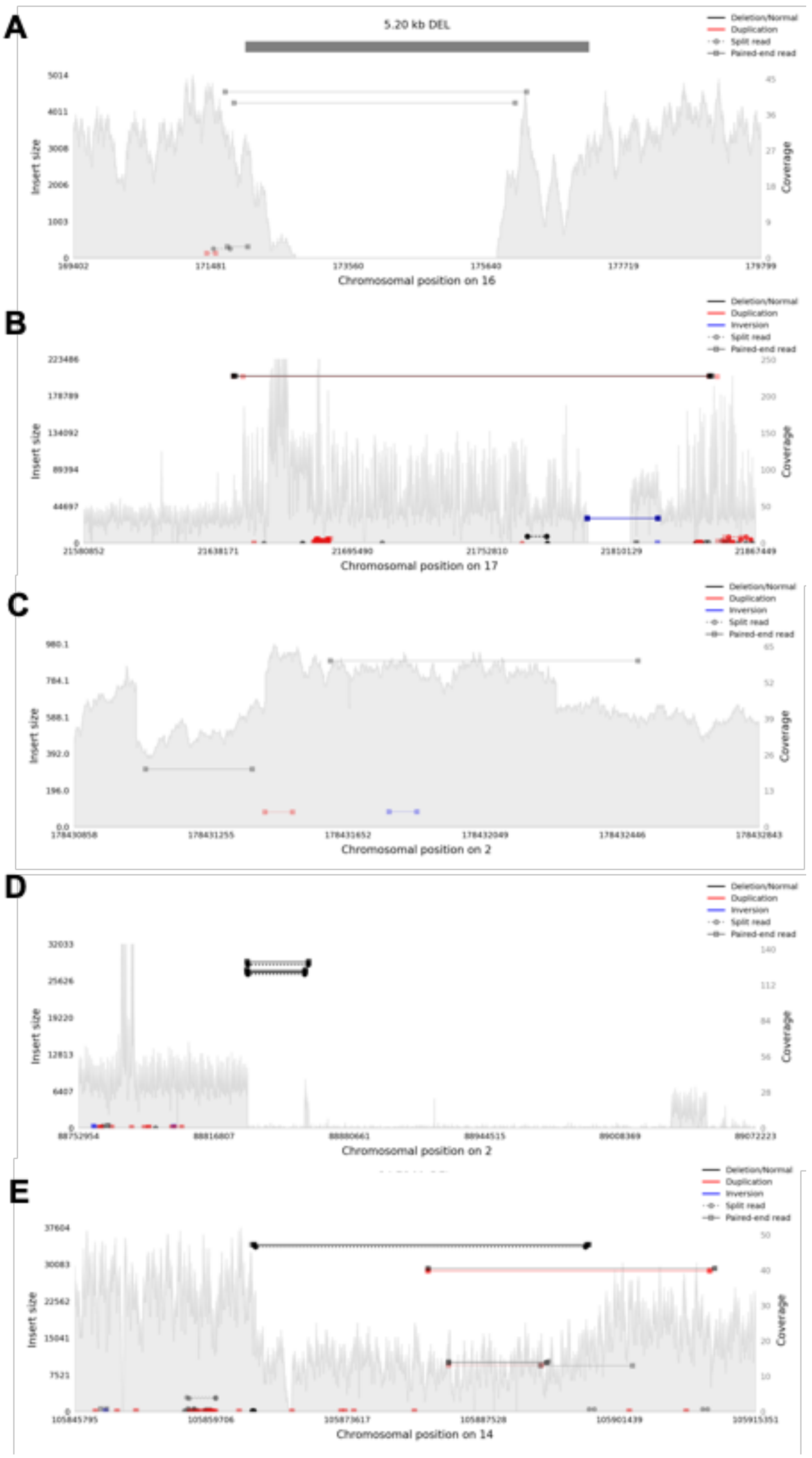
A
5.20 kb DEL
Deletion/Normal
Duplication
Split read
Paired-end read
Insert size
Coverage
Chromosomal position on 16
B
Deletion/Normal
Duplication
Inversion
Split read
Paired-end read
Insert size
Coverage
Chromosomal position on 17
C
Deletion/Normal
Duplication
Inversion
Split read
Paired-end read
Insert size
Coverage
Chromosomal position on 2
D
Deletion/Normal
Duplication
Inversion
Split read
Paired-end read
Insert size
Coverage
Chromosomal position on 2
E
Deletion/Normal
Duplication
Inversion
Split read
Paired-end read
Insert size
Coverage
Chromosomal position on 14

**Figure S5.**

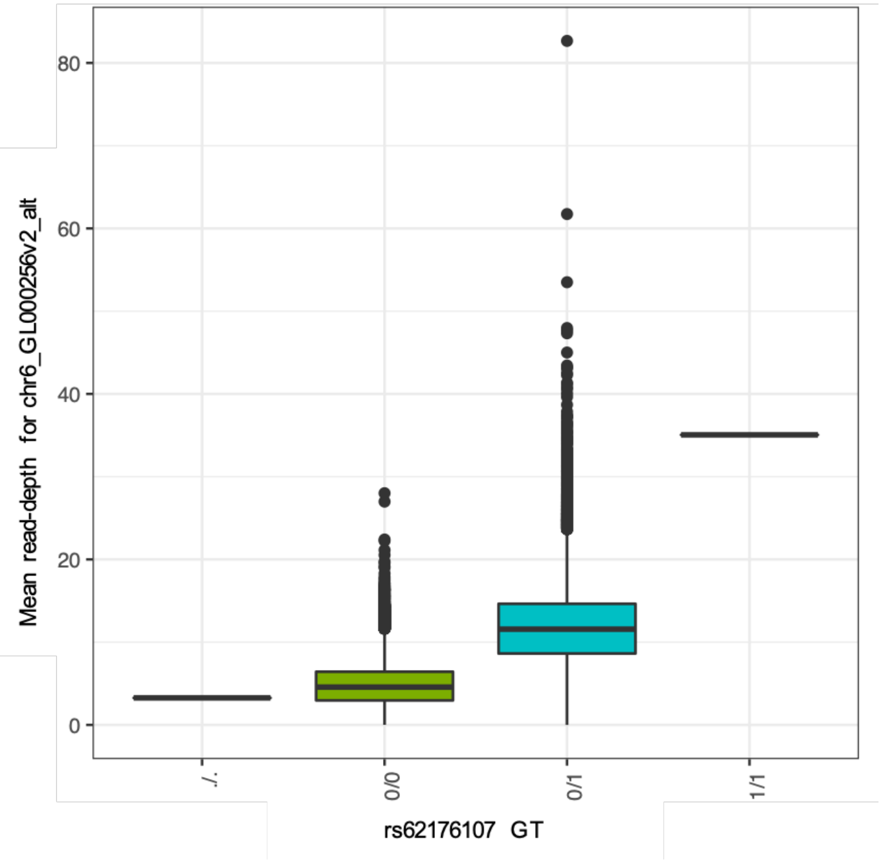

Mean read-depth for chr6_GL000256v2_alt
80
60
40
20
0
./.
0/0
0/1
1/1
rs62176107 GT

**Figure S6.**

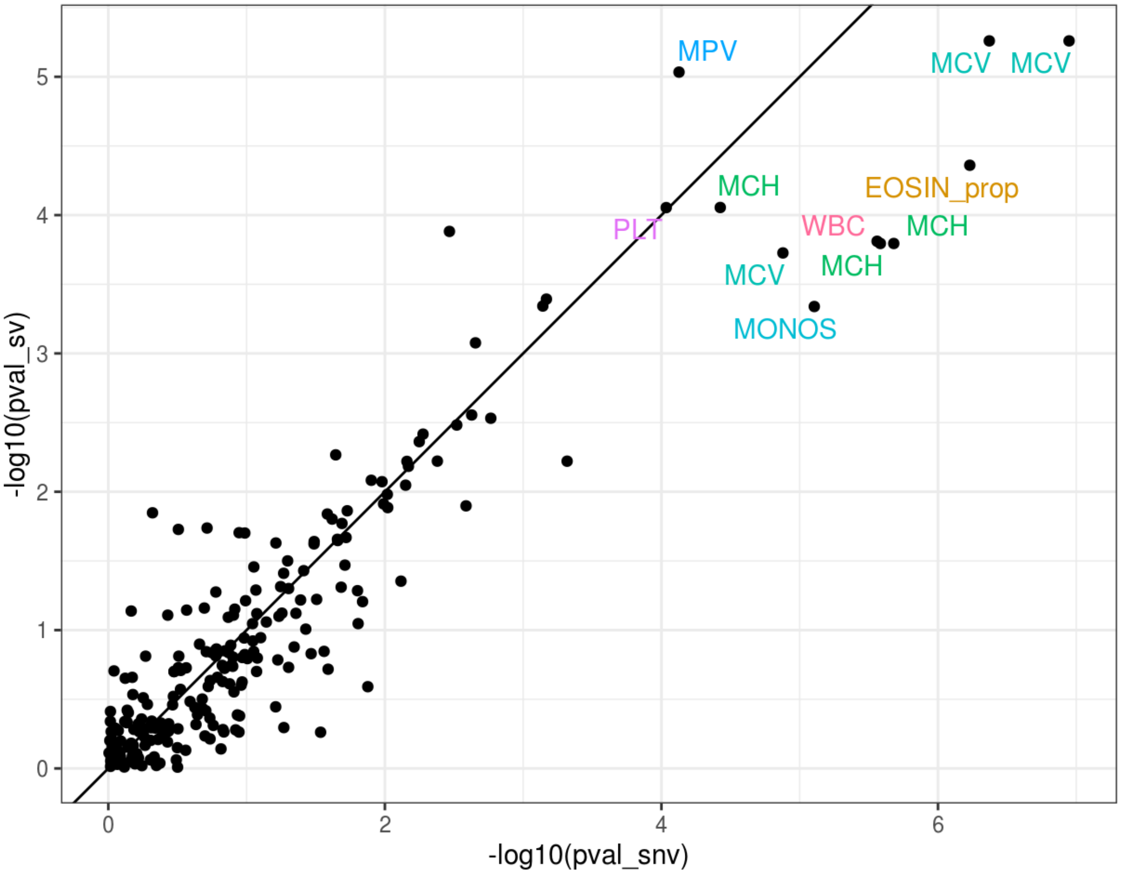
MPV
MCV
MCV
MCH
EOSIN_prop
PLT
WBC
MCH
MCV
MCH
MONOS
-log10(pval_sv)
-log10(pval_snv)
0
1
2
3
4
5
0
2
4
6
