## Supplementary_materials for "Whole genome sequencing identifies common and rare structural variants contributing to hematologic traits in the NHLBI TOPMed program": Supplemental_methods.docx

**Participating TOPMed studies**

*Amish*

The Amish Complex Disease Research Program includes a set of large community-based studies focused largely on cardiometabolic health carried out in the Old Order Amish (OOA) community of Lancaster, Pennsylvania (http://medschool.umaryland.edu/endocrinology/amish/research-program.asp). The OOA population of Lancaster County, PA immigrated to the Colonies from Western Europe in the early 1700’s. There are now over 30,000 OOA individuals in the Lancaster area, nearly all of whom can trace their ancestry back 12-14 generations to approximately 700 founders. Investigators at the University of Maryland School of Medicine have been studying the genetic determinants of cardiometabolic health in this population since 1993. To date, over 7,000 Amish adults have participated in one or more of our studies.

Due to their ancestral history, the OOA are enriched for rare exonic variants that arose in the population from a single founder (or small number of founders) and propagated through genetic drift. Many of these variants have large effect sizes and identifying them can lead to new biological insights about health and disease. The parent study for this WGS project provides one (of multiple) examples. In our parent study, we identified through a genome-wide association analysis a haplotype that was highly enriched in the OOA that is associated with very high LDL-cholesterol levels. At the present time, the identity of the causative SNP – and even the implicated gene – is not known because the associated haplotype contains numerous genes, none of which are obvious lipid candidate genes. A major goal of the WGS that will be obtained through the NHLBI TOPMed Consortium will be to identify functional variants that underlie some of the large effect associations observed in this unique population.

*ARIC*

The ARIC study is a population-based prospective cohort study of cardiovascular disease sponsored by the National Heart, Lung, and Blood Institute (NHLBI). ARIC included 15,792 individuals, predominantly European American and African American, aged 45-64 years at baseline (1987-89), chosen by probability sampling from four US communities. Cohort members completed three additional triennial follow-up examinations, a fifth exam in 2011-2013, a sixth exam in 2016-2017, and a seventh exam in 2018-2019. The ARIC study has been described in detail previously (The ARIC Investigators. The Atherosclerosis Risk in Communities (ARIC) study: Design and objectives. American Journal of Epidemiology 1989;129:687-702).

*BioMe*

The Charles Bronfman Institute for Personalized Medicine at Mount Sinai Medical Center (MSMC), BioMe Biobank, founded in September 2007, is an ongoing, broadly-consented electronic health record-linked clinical care biobank that enrolls participants non-selectively from the Mount Sinai Medical Center patient population. The MSMC serves diverse local communities of upper Manhattan, including Central Harlem (86% African American), East Harlem (88% Hispanic/Latino), and Upper East Side (88% Caucasian/White) with broad health disparities.

*CARDIA*

The Coronary Artery Risk Development in Young Adults (CARDIA) Study is a study examining the development and determinants of clinical and subclinical cardiovascular disease and their risk factors. It began in 1985-1986 with a group of 5,115 black and white men and women aged 18-30 years. The participants were selected so that there would be approximately the same number of people in subgroups of race, gender, education (high school or less and more than high school) and age (18-24 and 25-30) in each of 4 centers: Birmingham, AL; Chicago, IL; Minneapolis, MN; and Oakland, CA.

*CHS*

The Cardiovascular Health Study (CHS) is a population-based cohort study of risk factors for coronary heart disease and stroke in adults 65 years and older conducted across four field centers 2. The original predominantly European ancestry cohort of 5,201 persons was recruited in 1989-1990 from random samples of people on Medicare eligibility lists from four US communities. Subsequently, an additional predominantly African-American cohort of 687 persons was enrolled for a total sample of 5,888. Institutional review committees at each field center approved the CHS, and participants gave informed consent. Blood samples were drawn from all participants at their baseline examination, and DNA was subsequently extracted from available samples. These analyses were limited to participants with available DNA who also consented to genetic studies. Participants were examined annually from enrollment to 1999 and continued to be under surveillance for stroke following 1999.

*COPDGene*

COPDGene (also known as the Genetic Epidemiology of COPD Study) is an NIH-funded, multicenter study. A study population of more than 10,000 smokers (1/3 African American and 2/3 non-Hispanic White) has been characterized with a study protocol including pulmonary function tests, chest CT scans, six minute walk testing, and multiple questionnaires. Five years after this initial visit, all available study participants are being brought back for a follow-up visit with a similar study protocol. This study has been used for epidemiologic and genetic studies. Previous genetic analysis in this study has been based on genome-wide SNP genotyping data. Approximately 1,900 subjects underwent whole genome sequencing in this NHLBI WGS project, including severe COPD subjects and resistant smoking controls. The COPDGene Study web site is: http://www.copdgene.org/.

*FHS*

FHS is a three-generation, single-site, community-based, ongoing cohort study that was initiated in 1948 to investigate prospectively the risk factors for CVD including stroke. It now comprises 3 generations of participants: the Original cohort followed since 1948; their Offspring and spouses of the Offspring, followed since 1971 4; and children from the largest Offspring families enrolled in 2002 (Gen 3). The Original cohort enrolled 5,209 men and women who comprised two-thirds of the adult population then residing in Framingham, MA. Survivors continue to receive biennial examinations. The Offspring cohort comprises 5,124 persons (including 3,514 biological offspring) who have been examined approximately once every 4 years. The Gen 3 cohort contains 4,095 participants.

*GeneSTAR*

In 1982 The Johns Hopkins Sibling and Family Heart Study was created to study patterns of coronary heart disease and related risk factors in families with early-onset coronary disease, identified from10 Baltimore area hospitals. Renamed in 2003, the Genetic Study of Atherosclerosis Risk (GeneSTAR) continues to study mechanisms of coronary heart disease and stroke in families using novel models and exciting new methods. GeneSTAR is a family-based study including initially healthy brothers and sisters identified from probands with early-onset coronary disease, along with the healthy offspring of the siblings and the probands. The goal is to discover and amplify mechanisms of stroke and coronary heart disease. Our African American and European American family cohort has undergone extensive screening, genetic testing, and follow-up for new cardiovascular disease, stroke, and other clinical events for 5 to 38 years.

*HCHS/SOL*

The Hispanic Community Health Study/Study of Latinos (HCHS/SOL) is a multi-center study of Hispanic/Latino populations with the goal of determining the role of acculturation in the prevalence and development of diseases, and to identify other traits that impact Hispanic/Latino health. The study is sponsored by the National Heart, Lung, and Blood Institute (NHLBI) and other institutes, centers, and offices of the National Institutes of Health (NIH). Recruitment began in 2006 with a target population of 16,000 persons of Cuban, Puerto Rican, Dominican, Mexican or Central/South American origin. Participants were recruited through four sites affiliated with San Diego State University, Northwestern University in Chicago, Albert Einstein College of Medicine in Bronx, New York, and the University of Miami. Recruitment was implemented through a two-stage area household probability design. The study enrolled 16,415 participants who were self-identified Hispanic/Latino and aged 18-74 years, and the extensive psycho-social and clinical assessments were conducted during 2008-2011. Annual telephone follow-up interviews are ongoing since study inception. During the 2014-2017 second visit, the participants were re-examined again for various health outcomes of interest.

*JHS*

The Jackson Heart Study (JHS, https://www.jacksonheartstudy.org/jhsinfo/) is a large, community-based, observational study whose participants were recruited from urban and rural areas of the three counties (Hinds, Madison and Rankin) that make up the Jackson, MS metropolitan statistical area (MSA). Participants were enrolled from each of 4 recruitment pools: random, 17%; volunteer, 30%; currently enrolled in the Atherosclerosis Risk in Communities (ARIC) Study, 31% and secondary family members, 22%. Recruitment was limited to non-institutionalized adult African Americans 35-84 years old, except in a nested family cohort where those 21 to 34 years of age were also eligible. The final cohort of 5,306 participants included 6.59% of all African American Jackson MSA residents aged 35-84 during the baseline exam (N-76,426, US Census 2000). Among these, approximately 3,700 gave consent that allows genetic research and deposition of data into dbGaP, with 3406 participants with post-QC TOPMed whole genome sequencing data. Major components of three clinic examinations (Exam 1 – 2000-2004; Exam 2 – 2005-2008; Exam 3 – 2009-2013) include medical history, physical examination, blood/urine analytes and interview questions on areas such as: physical activity; stress, coping and spirituality; racism and discrimination; socioeconomic position; and access to health care. Extensive clinical phenotyping includes anthropometrics, electrocardiography, carotid ultrasound, ankle-brachial blood pressure index, echocardiography, CT chest and abdomen for coronary and aortic calcification, liver fat, and subcutaneous and visceral fat measurement, and cardiac MRI. At 12-month intervals after the baseline clinic visit (Exam 1), participants have been contacted by telephone to: update information; confirm vital statistics; document interim medical events, hospitalizations, and functional status; and obtain additional sociocultural information. Questions about medical events, symptoms of cardiovascular disease and functional status are repeated annually. Ongoing cohort surveillance includes abstraction of medical records and death certificates for relevant International Classification of Diseases (ICD) codes and adjudication of nonfatal events and deaths. CMS data are currently being incorporated into the dataset.

*MESA*

The MESA study is a study of the characteristics of subclinical cardiovascular disease (disease detected non-invasively before it has produced clinical signs and symptoms) and the risk factors that predict progression to clinically overt cardiovascular disease or progression of the subclinical disease 7. MESA researchers study a diverse, population-based sample of 6,814 asymptomatic men and women aged 45-84. Thirty-eight percent of the recruited participants are white, 28 percent African-American, 22 percent Hispanic, and 12 percent Asian, predominantly of Chinese descent. Participants were recruited from six field centers across the United States: Wake Forest University, Columbia University, Johns Hopkins University, University of Minnesota, Northwestern University and the University of California - Los Angeles.

*WHI*

The Women’s Health Initiative (WHI) is a long-term, prospective, multi-center cohort study that investigates post-menopausal women’s health. WHI was funded by the National Institutes of Health and the National Heart, Lung, and Blood Institute to study strategies to prevent heart disease, breast cancer, colon cancer, and osteoporotic fractures in women 50-79 years of age. WHI involves 161,808 women recruited between 1993 and 1998 at 40 centers across the US. The study consists of two parts: the WHI Clinical Trial which was a randomized clinical trial of hormone therapy, dietary modification, and calcium/Vitamin D supplementation, and the WHI Observational Study, which focused on many of the inequities in women’s health research and provided practical information about the incidence, risk factors, and interventions related to heart disease, cancer, and osteoporotic fractures.

**Genetic Ancestry and Relatedness**

Principal components (PCs) of genetic ancestry and pairwise relatedness measures were estimated for all 140,062 samples included in the TOPMed ‘Freeze 8’ data release. Autosomal genetic variants passing the quality filter with a MAF > 0.01 and missing call rate < 0.01 were LD-pruned with an r^2^ threshold of 0.1 to obtain a set of 638,486 effectively independent variants for genetic ancestry and relatedness estimation. PC-AiR was used to obtain ancestry informative PCs robust to familial relatedness; the first 11 PCs showed evidence of population structure. PC-Relate was then used to estimate pairwise kinship coefficients (KCs) for all pairs of samples, conditional on the genetic ancestry captured by PC-AiR PCs 1-11; these KC estimates reflect only recent genetic relatedness, e.g. due to pedigree structure. The PC-Relate KC estimates were used to construct a 4th degree sparse, block-diagonal, empirical kinship matrix (KM) for association testing, any pair of samples with estimated KC > 2^(-11/2) ~ 0.022 were clustered in the same block; all KC estimates within a block of samples were kept, regardless of value; and all KC estimates between blocks were set to 0. By using a sparse block-diagonal KM, the association tests are more computationally efficient yet recent genetic relatedness is still accounted for. We subset the freeze-wide PCs and sparse KM to the appropriate set of participants for each analysis.

**Classification of race/ethnicity groups using HARE**

Ancestry groups were based on a combination of participants reported race/ethnicity and genetic ancestry represented by PCs from PC-AiR. To infer race/population group membership for participants with missing values, we used the HARE method, a machine learning algorithm that uses a support vector machine (SVM) to determine stratum assignment, taking as input genetically estimated PC values and reported race/ethnicity for each participant. Strata are defined by the unique reported race/ethnicity values provided, then the HARE SVM uses the input (training) data to learn the probability of stratum membership across the entire PC space. The output of HARE consists of multinomial probability vectors of stratum membership for each participant. HARE was run on a subset of samples included in the TOPMed Freeze 8 data release; specifically, samples for participants from non-US-based studies and the Amish participants (because they were very distinct in PC space) were excluded from the HARE analysis. HARE was run using the first 9 PC-AiR PCs generated on this subset of samples to represent genetic ancestry with the following reported race/population groups: Asian, Black, Central American, Cuban, Dominican, Mexican, Puerto Rican, South American, and White. The genetic data from the 31,918 participants with either unreported or non-specific (e.g. ‘Multiple’ or ‘Other’) race and population membership was included in the HARE analysis, but they were not used to train the SVM. These participants were assigned to a population stratum based on their highest HARE output probability of membership. All other participants remained in the population stratum corresponding to their reported race/population group. Amish participants were assigned to their own stratum.
