## Supplementary figures and images for "Whole genome sequencing identifies common and rare structural variants contributing to hematologic traits in the NHLBI TOPMed program"

### Supplemental_figure_S7_sgRNAs_targeting_602bp_region.pdf

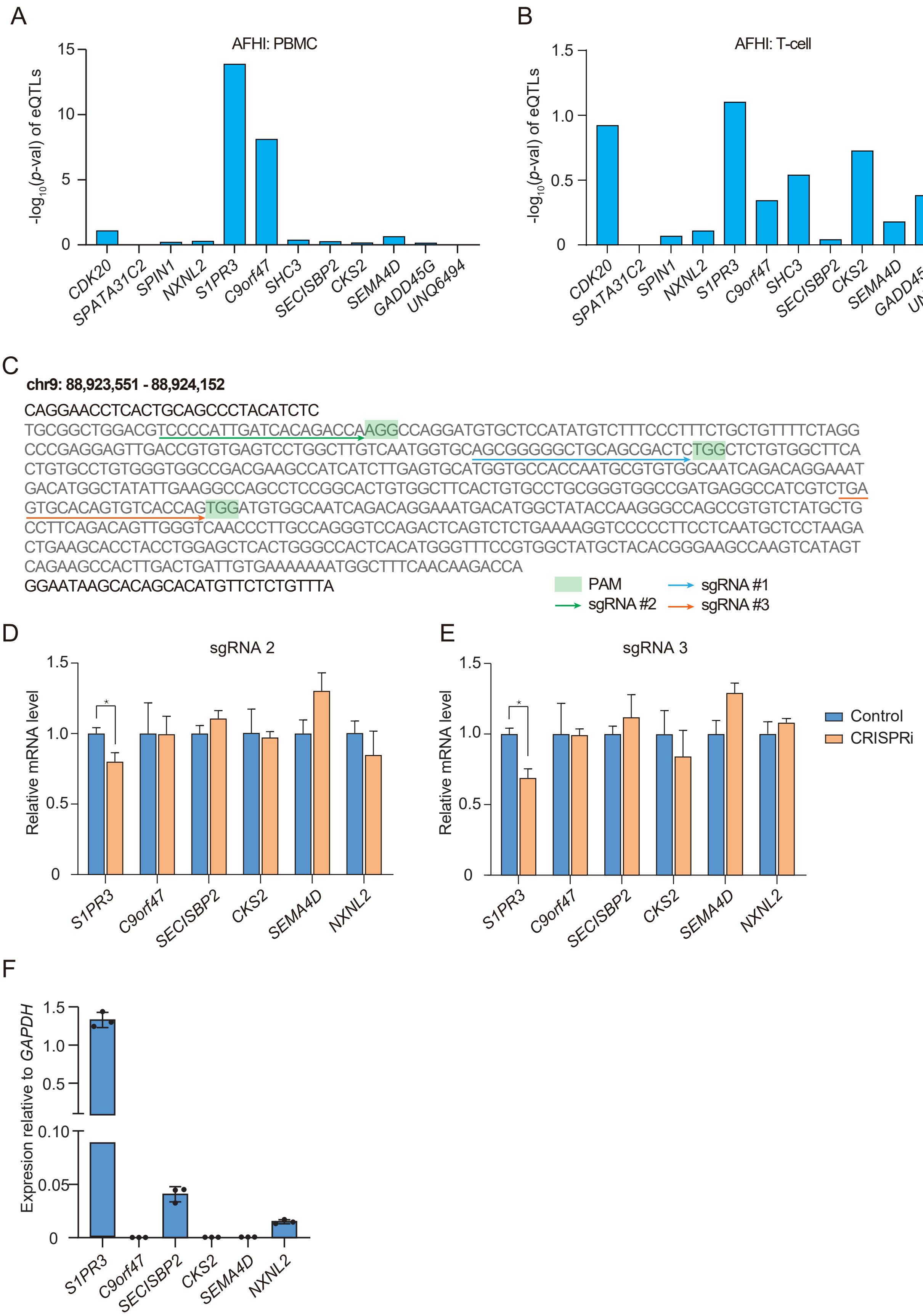

### Supplemental_figure_S8_eQTL_and_HiC_datasets.pdf

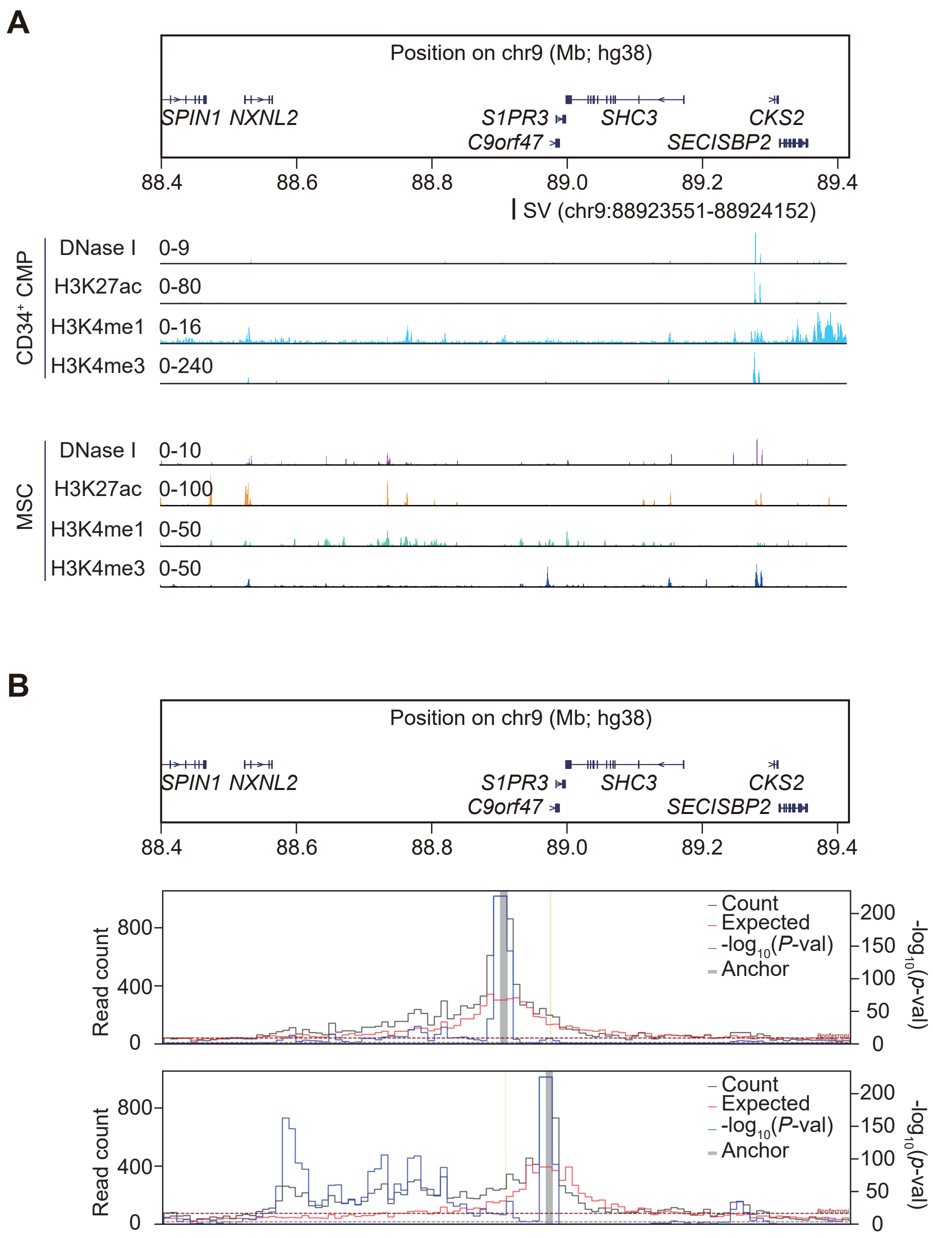

### Supplemental_figure_S9_Mice xenograft.pdf

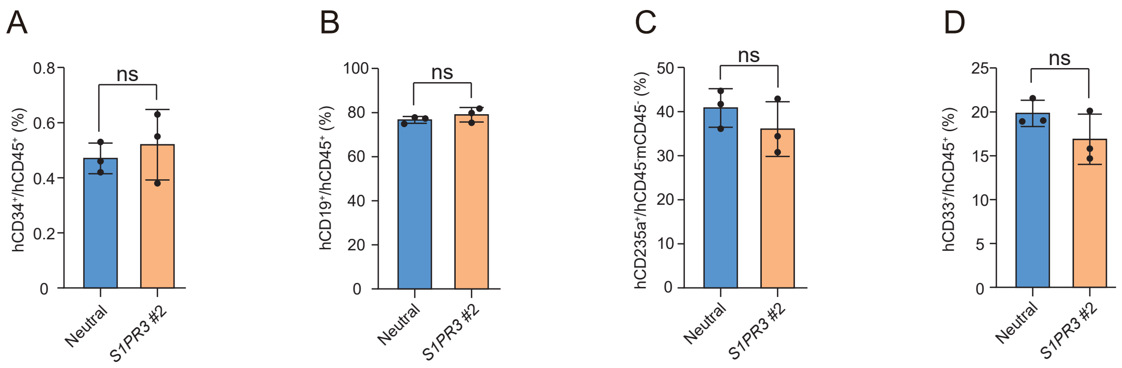
